# Low linoleic acid foods with added DHA given to Malawian children with severe acute malnutrition improves cognition: a randomized, triple blinded, controlled clinical trial

**DOI:** 10.1101/2021.09.07.21263231

**Authors:** Kevin Stephenson, Meghan Callaghan-Gillespie, Kenneth Maleta, Minyanga Nkhoma, Matthews George, Hui Gyu Park, Reginald Lee, Iona Humpheries-Cuff, R J Scott Lacombe, Donna R Wegner, Richard L Canfield, J Thomas Brenna, Mark J Manary

## Abstract

**Background:** There is concern that the PUFA composition of ready-to-use therapeutic food (RUTF) for treatment of severe acute malnutrition (SAM) is suboptimal for neurocognitive recovery.

**Objective:** We tested the hypothesis that RUTF made with reduced amounts of linoleic acid, achieved using high oleic (HO) peanuts, with or without added DHA, improves cognition when compared to standard RUTF (S-RUTF).

**Methods:** A triple-blind, randomized, controlled clinical feeding trial was conducted among children with uncomplicated SAM in Malawi with 3 types of RUTF; DHA-HO-RUTF, HO-RUTF and S-RUTF. The primary outcomes, measured in a subset of subjects, were the Malawi Developmental Assessment Tool (MDAT) global and 4 domain (gross motor, fine motor, language and social) z-scores and a modified Willatts problem solving assessment (PSA) intention score for 3 standardized problems, measured 6 months and immediately after completing RUTF therapy, respectively. Plasma fatty acid content, anthropometry and eye tracking were secondary outcomes. Comparisons were made between the novel PUFA RUTFs and S-RUTF.

**Results:** Among the 2565 SAM children enrolled, global MDAT z-score was -0.69 ± 1.19 and - 0.88 ± 1.27 for children receiving DHA-HO-RUTF and S-RUTF, respectively (difference 0.19, 95% CI 0.01 to 0.38). The gross motor and social domains had higher z-scores among children receiving either DHA-HO-RUTF than S-RUTF. The PSA problem 3 scores did not differ by dietary group (Odds ratio 0.92, 95% CI 0.67 to 1.26 for DHA-HO-RUTF). After 4 weeks of treatment, plasma phospholipid EPA and α-linolenic acid were greater in children consuming DHA-HO-RUTF or HO-RUTF when compared to S-RUTF (for all 4 comparisons *P* values < 0.001), but only plasma DHA was greater in DHA-HO-RUTF than S-RUTF (*P* <0.001).

**Conclusions:** Treatment of uncomplicated SAM with DHA-HO-RUTF resulted in an improved MDAT score, conferring a cognitive benefit six months after completing diet therapy. This treatment should be explored in operational settings.

## Introduction

Severe acute malnutrition (SAM) is a global insult to the young child’s developing mind and body. Therapeutic foods for SAM were largely designed to provide the nutrients known to affect recovery of anthropometry and muscle mass (1). Vegetable oil rich, peanut-based ready-to-use therapeutic food (RUTF) is the standard of care for most SAM because it can be used safely at home in the context of utmost poverty (2). It has been assumed that the brain and other viscera would receive what is needed for recovery from standard RUTF (S-RUTF).

Dietary omega-3 PUFAs are essential to normal brain development and function. Brain accretion of the key neural structural component, omega-3 DHA, accelerates in the third trimester of gestation and continues until age 18 yr (3,4). Tissue DHA is derived from either preformed dietary DHA or a precursor, typically plant-derived α-linolenic acid (ALA). ALA is less efficient in supplying brain DHA than dietary DHA (5). Omega-6 linoleic acid (LA) is metabolized by the same desaturases (*FADS2, FADS1*) and elongases (*ELOVL2, ELOVL5*) as omega-3 PUFA, and uses the same pathways to incorporate into membranes (6,7). LA intake required to eliminate overt clinical abnormalities and support growth in infants is about 2% of energy or about 1% of fatty acids from diets deriving half their energy from fat (8). RUTF and breastmilk are examples of such diets. Suppression of omega-3 PUFA tissue accretion by excess dietary LA is an impactful clinical phenomenon (9). High LA dietary vegetable oils create a metabolic demand for all omega-3 PUFA, thus requiring higher dietary DHA intake to produce similar tissue DHA levels (10).

The development of peaceful and just citizens requires a high-level mental functioning to support executive problem solving and the affect regulation needed for resilient responses. Perinatal brain development depends on a balanced supply of brain specific nutrients. Overwhelming amounts of LA in vegetable oils suppress trace amounts of ALA when the brain is rapidly accreting DHA. This inhibits neurobehavioral development at the levels of gene expression, hormonal balance, affect, and adaptive responses to environmental cues (11). Omega-3 deficient diets cause poor impulse control leading to potentiated stress response, increased depression and aggression, and poorer cognitive performance (12-14). Evidence-based guidelines include EPA and DHA supplements as a treatment for attention deficit hyperactivity disorder, autism spectrum disorder, and major depressive disorder (15,16).

The potential effects of excess LA and limited ALA in RUTF on cognition have not been measured. Our previous study showed that RUTF with excess LA caused a 25% reduction in circulating DHA in 4 weeks (17). This clinical trial tested the hypothesis that RUTF made with reduced amounts of LA, achieved by using high oleic (HO) peanuts and palm oil (HO-RUTF) with or without added DHA (DHA-HO-RUTF), confers lasting cognition improvement when compared to S-RUTF. Cognition was measured by the Malawi Developmental Assessment Tool (MDAT), a standardized, validated battery of exercises, and a modified Willatts problem solving assessment (PSA) intention score.

## Methods

### Study design

This triple-blind, randomized controlled clinical trial compared cognition in children treated for uncomplicated SAM receiving one of 3 RUTFs; HO-RUTF, DHA-HO-RUTF, or S-RUTF. The primary outcomes were the MDAT global z-score and PSA intention scores. Secondary outcomes included MDAT domain z-scores, recovery rates, anthropometric growth rates, Saccadic response time, visual paired comparison, novelty preference score, mean fixation during familiarization, and adverse events. Detailed descriptions of the study methods are provided in the study protocol available as supplementary material.

All sample size calculations were performed using G*Power (3.1.9.7) with two-tailed testing at a significance level of 0.05 and power of 0.80 (18). A total of 300 children per group were required to detect a difference in MDAT global z-score of 0.25, assuming a SD of 1.1 (19-21). The difference of 0.25 z-scores was chosen based on prior nutrition-based effect sizes on developmental scores as well as MDAT-specific changes seen in association with an illness (0.14 -0.19) (22,23). Assuming a 25% loss to follow-up, 400 participants per group were designated for testing. For the PSA, we assumed 20% of participants would not complete testing; thus, a total of 300 children per group were needed to detect a standardized effect size of 0.25. This effect size was chosen based on previous studies of healthy infants under controlled conditions, where standardized effects sizes ranged from 0.4-0.5 (24,25). Given the extended age range relative to prior trials, the proximity of participants to SAM, and testing environs, we chose a smaller effect size for sample size calculations.

The trial sample size was determined based on the key secondary outcome, recovery. A total of 900 participants per group was required to detect an improvement in recovery of 4% assuming an expected recovery rate of 89% and default rate of 5%, both of which were determined based on prior operational experience in Malawi (26,27).

Participants were randomized individually in a 1:1:1 ratio into the three intervention groups. Computer-generated block randomization lists were created in blocks of 54 using six colors, two of which corresponded to each food. Participants were assigned to their food group when they drew an opaque envelope containing one of the colored cards.

Study foods were dispensed in identical packets with a colored sticker. The three study foods underwent extensive testing to minimize differences in appearance and taste. Excepting one remote member of the study team, all research personnel, including nurses, investigators, and laboratory personnel, were blinded to the color code. This unblinded individual did not evaluate any of the study subjects, nor did she analyze the primary outcomes. The trial is registered at clinicaltrials.gov as NCT03094247.

### Subjects and setting

The trial was conducted at 28 clinics in rural Southern Malawi between October 2017 and December 2020. Most families in the study farm for subsistence, with corn as the staple crop. Breastfeeding is ubiquitous throughout the first year of life. Homes are built from mud with thatch or corrugated metal sheet roofing. Plumbing and electricity are exceedingly rare, and water is obtained from boreholes, wells, or rivers.

Children aged 6-59 mo were eligible for enrollment if they had uncomplicated SAM, which was defined as a mid-upper arm circumference (MUAC) < 11.5 cm, and/or weight-for-height z-score < -3, and/or bilateral pitting edema, with an adequate appetite as determined by a 30g test feeding. Exclusion criteria included participation in a feeding program in the prior 1 month, presence of developmental delay, chronic debilitating medical condition, peanut allergy, or a hearing or vision problem.

The trial was approved by the Human Studies Committee of Washington University and the College of Medicine Research and Ethics Committee of the University of Malawi. Nurses fluent in Chichewa explained the trial to each child’s caregiver and obtained verbal and written consent.

### Participation

Upon enrollment, baseline demographic, socioeconomic, and health data were collected. Weight, length, and MUAC were measured. Caregivers received counseling, a two-week supply of RUTF, and a seven-day course of amoxicillin. Healthy twins of subjects were given an allotment of RUTF as well. Follow-up visits were scheduled at 2-week intervals, at which time anthropometric measurements were repeated and caregivers were asked about adherence and symptoms. Children were fed until reaching a clinical outcome or for a maximum of six bi-weekly follow-up visits. The anthropometry methods and definitions of the clinical outcomes are described in the study protocol.

Beginning in February 2018, all participants under two years of age were assessed with a PSA within four weeks of their clinical outcome. Beginning in March 2018, all participants under 30 mo of age were asked to return to clinic 5 to 7 mo later for testing with the MDAT and Tobii X2-60 eye tracker eye-tracking testing.

Children that either missed fortnightly assessments during therapeutic feeding or neurocognitive testing were sought on two extra occasions by health surveillance assistants and asked to return for the scheduled treatment or testing.

### Study foods

All three RUTF formulations were produced by Project Peanut Butter in Lunzu, Malawi, meeting all international quality and nutrient specifications. DHA-HO-RUTF and HO-RUTF were formulated to reduce LA and increase the omega-3 PUFA content (**Table 1, Supplementary Tables 1-2**, protocol section 6.5). Reductions in LA were achieved by using high oleic peanuts. Increases in omega-3 PUFA were achieved by the addition of perilla oil. DHA was enhanced in one of the RUTFs by the addition of encapsulated fish oil. The formulation was demonstrated to be acceptable in a pilot trial (17).

**Table 1.** Composition of study ready-to-use therapeutic foods^1^

|  | DHA-HO-RUTF | HO-RUTF | S-RUTF |
| --- | --- | --- | --- |
| **Ingredient** |  |  |  |
| Milk powder, non-fat, dry, g | 22.0 | 22.0 | 27.3 |
| Milk, sweet whey powder, g | 6.1 | 6.1 | 0.0 |
| Palm oil, g | 14.0 | 14.0 | 16.0 |
| Canola oil, g | 0.0 | 0.0 | 5.1 |
| Sugar, g | 24.5 | 26.0 | 23.0 |
| High oleic peanuts, g | 25.0 | 25.0 | 0.0 |
| Standard, high omega6 peanuts, g | 0.0 | 0.0 | 23.7 |
| Perilla oil, g | 3.0 | 3.0 | 0.0 |
| Hydrogenated vegetable oil<sup>†</sup>, g | 1.0 | 1.0 | 2.0 |
| Micronutrient powder, g | 2.9 | 2.9 | 2.9 |
| DHA, g | 1.5 | 0.0 | 0.0 |
| **Macronutrient** |  |  |  |
| Energy, kcal | 532.0 | 531.0 | 541.0 |
| Protein, g | 15.2 | 15.2 | 15.7 |
| Protein, dairy, g | 8.9 | 8.9 | 9.8 |
| Lipids, g | 30.2 | 29.5 | 32.7 |
| Omega-6 polyunsaturated fatty acids, g | 2.2 | 2.1 | 5.8 |
| Omega-3 polyunsaturated fatty acids, g | 1.5 | 1.4 | 0.5 |
| DHA, mg | 72.0 | 0.0 | 0.0 |
| EPA, mg | 14.0 | 0.0 | 0.0 |

<sup>1</sup>Content expressed in per 100g. DHA, docosahexaenoic acid; DHA-HO-RUTF, docosahexaenoic acid added to RUTF made with high oleic acid peanuts; EPA, eicosapentaenoic acid; HO-RUTF, RUTF made with high oleic acid peanuts; RUTF, ready-to-use therapeutic food; S-RUTF, standard RUTF.
<sup>†</sup>This was a proprietary commercial product, Trancendim 180 (Caravan Foods)

### Neurocognitive testing

MDAT has been validated within the study context (28). We performed MDAT assessments between five and seven months after a SAM outcome was reached to assess the effect of the RUTFs on neurodevelopment. No intervention was performed between the SAM outcome and the MDAT testing. MDAT contains 136 items across four domains, three of which are assessed by direct observation of the child (gross motor, fine motor, and language), while the fourth (social) is assessed via caregiver interview. These domain scores are then combined into a global score and compared to normal population reference values to provide age-adjusted z-scores (28). In line with prior protocols, failure of any six consecutive items resulted in failure of that domain and skipping to the subsequent domain (19). Global z-score outliers were defined after evaluation of the score distribution, and values < -5 were excluded.

A modified version of the Willatts PSA was performed within four weeks of the outcome from treatment of SAM (24). This time point was chosen to detect the immediate effect of the intervention RUTFs. The assessment was modified to accommodate testing a wider age range than those used in prior studies and so contained three problems of increasing complexity (25). The goal of the assessment was to judge intentionality in means-end task completion. All assessments were video recorded for subsequent coding.

Each problem contained a barrier between the child and the goal, a toy of her or his choosing. In problem 1, the toy was placed on a cloth and distanced from the child such that the cloth had to be pulled by the child to reach the toy. In problem 2, the toy was covered with an opaque cloth such that the child had to remove the cover to obtain the toy. Problem 3 was an amalgam of problems 1 and 2, wherein the toy was placed on a cloth and under a cover, such that the child had to pull the apparatus toward them and uncover the toy to obtain it.

Videos were coded using Behavioral Observation Research Interactive Software (29). Coders were trained to achieve >.90 agreement by kappa statistic for intention scores on a random sample of 20 test videos before coding for the trial. High reliability was maintained throughout the coding period by double-coding >50% of videos followed by a joint resolution of discrepancies. In a modification of the procedure of Willatts et al problem solving behaviors, problems were coded for two primary components, each on a scale of 0-2 and each assessing an aspect of intentionality: attentional orienting to each problem subgoal and the execution of behaviors on each subgoal. Problems 1 and 2 were scored 0-4, while problem 3 had two subgoals (moving cloth and removing the cover) and thus was scored 0-8. “No score” was a possible outcome for all problems, and reasons for this result were recorded.

Two eye-tracking tests were performed as secondary cognitive outcomes: a visual paired comparison task, and the infant orienting with attention task. Identical procedures to those previously described were used (20). The visual paired task consisted of 4 trials of African faces. The infant orienting task measured saccadic reaction time in a standardized manner. The testing sequence for paired comparison task and orienting attention task was randomized.

All neurocognitive assessors underwent extensive training and were required to pass periodic evaluations before administering the tests and throughout the trial. Please see the study protocol for further details.

### Plasma sampling and analysis

The purpose of blood sampling was to characterize the concentrations of fatty acids in the study groups, thus only a subset of children were sampled. Blood collection and analysis followed best practices (30). After receiving 4 weeks of RUTF, blood was collected by venipuncture and placed in an ethylenediaminetetraacetic acid tube. Plasma was then separated from red cells and held at -20°C until transferred to the laboratories of JT Brenna at the Dell Pediatric Research Institute in Austin, TX. Plasma phospholipids were chosen as the preferred lipid pool because they respond in a matter of weeks to changes in the diet compared to months for red blood cells, but are stable to fatty acids from recent meals which are largely triacylglycerols. Red blood cells PUFA also degrade at -20°C by peroxidation catalyzed by iron release from ruptured cells (30).

For analyses, plasma phospholipids were isolated using an automated three-phase liquid-liquid extraction method (31). Samples were then transmethylated to generate fatty acid methyl esters according to our routine methods (32). These fatty acid derivatives were detected and quantified by gas chromatography (GC)-flame ionization detection and structure confirmed by a specialized GC tandem mass spectrometry method. Quantitative standards of 25 fatty acid methyl esters were then used to quantify the amounts of each detected. Results are reported as a weight percentage of total fatty acid identified. These methods are carefully detailed in the study protocol.

### Statistical analysis

Analysis were performed by a blinded investigator using a modified intention to treat methodology wherein children who were discovered not to meet enrollment criteria were excluded from analysis. Baseline characteristics were summarized as means ± SD, medians (IQR), or percentages. Anthropometric indices were calculated with WHO Anthro version 3.1 (WHO). Rates of weight, MUAC, and length gain were calculated over the entire period of feeding.

Outcomes from the DHA-HO-RUTF and HO-RUTF groups were compared to S-RUTF. Continuous outcomes were compared using Student’s *t*-test. Categorical outcomes were compared using the chi-square test. PSA scores were analyzed using ordinal logistic regression with age as a covariate. An interaction term between intervention group and age was offered into the model but was not significant for any problem and thus was not included in the final model. The proportional odds assumption was assessed graphically and deemed not violated by producing a series of binary logistic regressions at cut-points for all outcome values > 2. Significance testing was restricted to pairwise comparisons of the two primary outcomes and key secondary outcomes, namely the domain MDAT z-scores and recovery rates. In all other cases, differences with unadjusted 95% CIs were calculated and reported. Analysis was completed using R version 4.0.1 (R Foundation for Statistical Computing) and SPSS version 27 (SPSS Inc.) (33).

## Results

Of the 2758 children with SAM identified between October 2017 and December 2020, 2565 children were included in the study (**Figure 1**). In 2020, there were three stock-outs of RUTFs due to ingredient importation restrictions related to the COVID-19 pandemic. This precluded randomization as it was designed, and so subjects were randomized among the available RUTFs. This is described in the study protocol available as supplementary material. Baseline characteristics were similar among the three groups, both in the entire study sample and within the MDAT and problem solving assessment subgroups (**Table 2, Supplementary Tables 3-4**).

**Table 2.** Baseline characteristics of Malawian children receiving study foods^1^

|  | DHA-HO-RUTF<br>(N = 809) | HO-RUTF<br>(N = 860) | S-RUTF<br>(N = 896) |
| --- | --- | --- | --- |
| Female sex, no. (%) | 452 (55.9) | 469 (54.5) | 527 (58.8) |
| Age, mo., median (IQR) | 12.1 (7.9 - 20.4) | 12.2 (7.8 - 21.2) | 12.7 (8.1 - 21.5) |
| Edematous, no. (%) | 274 (33.9) | 302 (35.1) | 293 (32.7) |
| Mid-upper-arm circumference, cm | 11.6 ± 1.2 | 11.6 ± 1.2 | 11.6 ± 1.1 |
| MUAC < 11.5 cm, no. (%) | 534 (66) | 545 (63.4) | 600 (67) |
| Weight-for-height z-score | -1.9 ± 1.2 | -1.9 ± 1.2 | -2.0 ± 1.2 |
| WHZ ≤ -3, no. (%) | 167 (20.6) | 181 (21.0) | 186 (20.8) |
| Height-for-age z-score | -3.3 ± 1.5 | -3.3 ± 1.4 | -3.2 ± 1.4 |
| HAZ ≤ -2, no./total no. (%) | 667/808 (82.5) | 726/858 (84.6) | 746/896 (83.3) |
| Fever in past 2 weeks, no./total no. (%) | 461/805 (57.3) | 518/857 (60.4) | 554/893 (62.0) |
| Diarrhea in past 2 weeks, no./total no. (%) | 446/805 (55.4) | 474/858 (55.2) | 484/891 (54.3) |
| Child breastfed, no./total no. (%) | 520/808 (64.4) | 565/858 (65.9) | 575/893 (64.4) |
| HIV-seropositive, no./total no. tested (%) | 19/265 (7.2) | 25/301 (8.3) | 23/311 (7.4) |
| Mother alive, no./total no. (%) | 783/807 (97) | 827/860 (96.2) | 876/895 (97.9) |
| Number of siblings, median (IQR) | 2 (0-3) | 2 (1-3) | 2 (0-3) |
| Thatch roof, no./total no. (%) | 607/803 (75.6) | 659/859 (76.7) | 689/890 (77.4) |
| Radio in home, no./total no. (%) | 178/807 (22.1) | 196/854 (23.0) | 168/893 (18.8) |
| Clean water source, no./total no. (%) | 761/807 (94.3) | 795/857 (92.8) | 834/895 (93.2) |

<sup>1</sup>Plus-minus values are means ± SD. ART, antiretroviral therapy; DHA-HO-RUTF, docosahexaenoic acid added to ready-to-use therapeutic food made with high-oleic acid peanuts; HAZ, height-for-age z-score; HIV, human immunodeficiency virus; HO-RUTF, RUTF with high oleic acid peanuts; IQR, interquartile range; MUAC, mid-upper arm circumference; WHZ, weight-for-height z-score.

**Figure 1.**
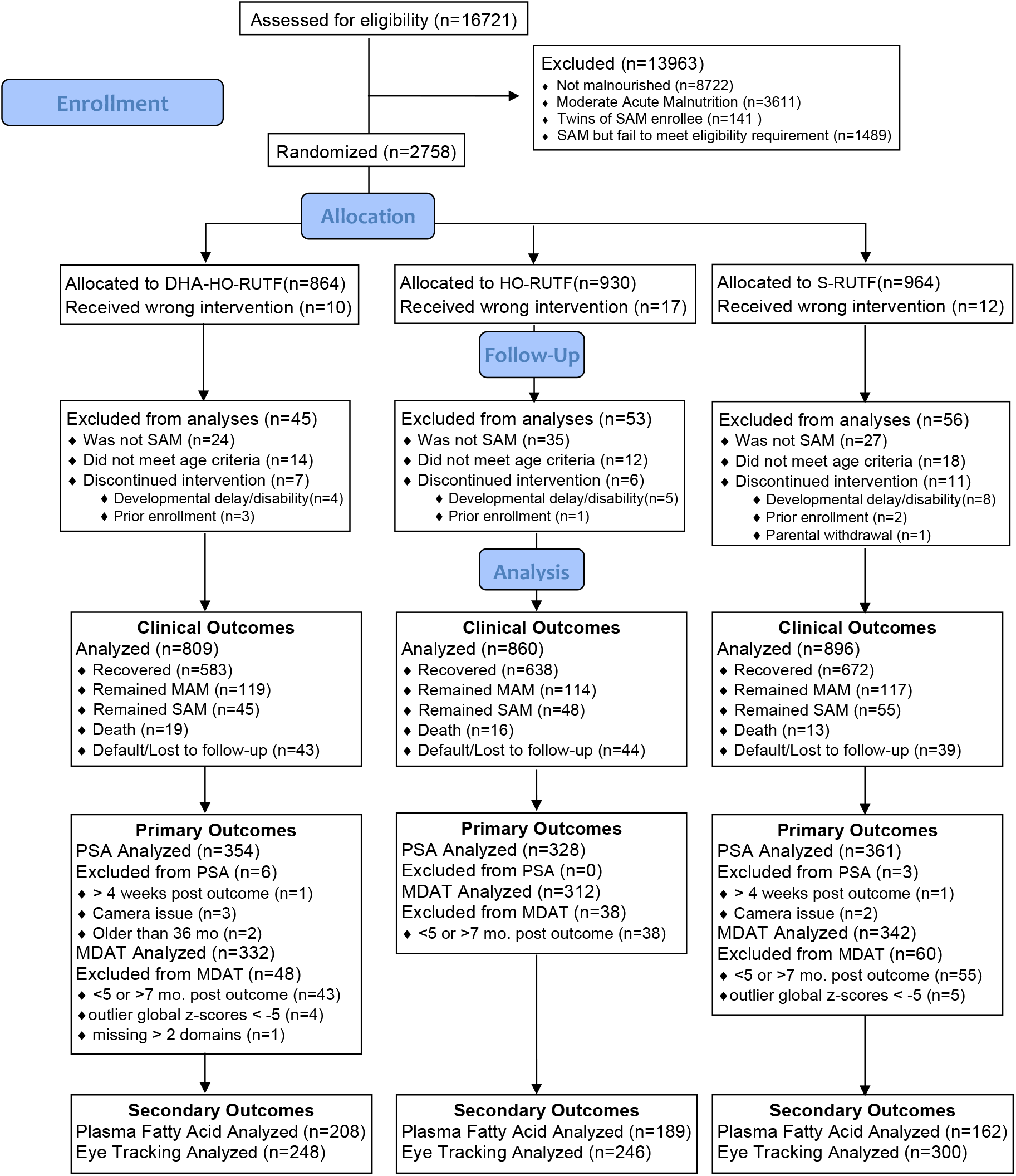
CONSORT diagram of study participation.

Children receiving DHA-HO-RUTF had higher MDAT global, gross motor, and social z-scores than those receiving S-RUTF, while children receiving HO-RUTF had higher social z-scores than those receiving S-RUTF at about 6 months post SAM outcome (**Table 3, Supplementary Table 3**). Probability density distributions for global and domain MDAT z-scores for each group are shown in **Figure 2**, while means are shown in **Supplementary Figure 1**. When stratified by age, the probability density distributions of global MDAT z-score show that children <12 mo at the time of SAM resemble healthy children, while children >18 mo at the time of SAM display scores approximately-2 z-scores worse. (**Supplementary Figure 2**).

**Table 3.** Outcomes and adverse events, according to intervention group^1^

|  | DHA-HO-RUTF |  | HO-RUTF |  | S-RUTF |  | DHA-HO-RUTF vs.<br>S-RUTF<br>Comparison† | HO-RUTF vs.<br>S-RUTF<br>Comparison† |
| --- | --- | --- | --- | --- | --- | --- | --- | --- |
|  | N |  | N |  | N |  |  |  |
| **Cognitive Outcomes** |  |  |  |  |  |  |  |  |
| Malawi Developmental Assessment Tool Z-scores |  |  |  |  |  |  |  |  |
| Global | 332 | -0.69 ± 1.19 | 312 | -0.80 ± 1.25 | 342 | -0.88 ± 1.27 | 0.19 (0.01 to 0.38)<sup>a</sup> | 0.08 (-0.11 to -.27) |
| Gross motor domain | 331 | -1.08 ± 1.66 | 312 | -1.34 ± 1.67 | 342 | -1.37 ± 1.78 | 0.29 (0.03 to 0.55)<sup>b</sup> | 0.02 (-0.24 to 0.29) |
| Fine motor domain | 323 | -1.81 ± 1.71 | 306 | -1.83 ± 1.80 | 337 | -1.91 ± 1.83 | 0.14 (-0.17 to 0.37) | 0.07 (-0.21 to 0.35) |
| Language domain | 331 | -0.73 ± 1.35 | 312 | -0.93 ± 1.51 | 342 | -0.90 ± 1.41 | 0.17 (-0.04 to 0.38) | -0.03 (-0.25 to 0.19) |
| Social domain | 332 | 0.40 ± 1.01 | 311 | 0.49 ± 1.00 | 342 | 0.24 ± 1.04 | 0.16 (0.00 to 0.31)<sup>c</sup> | 0.24 (0.09 to 0.40)<sup>d</sup> |
| Problem Solving Assessment Intention Scores |  |  |  |  |  |  |  |  |
| Problem 1, no. (%) with perfect score | 298 | 90 (30) | 281 | 88 (31) | 292 | 90(31) | 0.99 (0.66 to 1.19) ‡ | 0.89 (0.74 to 1.31) ‡ |
| Problem 2, no. (%) with perfect score | 293 | 169 (58) | 283 | 151 (53) | 294 | 168 (57) | 0.97 (0.71 to 1.33) ‡ | 0.88 (0.64 to 1.21) ‡ |
| Problem 3, no. (%) with perfect score | 236 | 50 (21) | 241 | 54 (22) | 238 | 51 (21) | 0.92 (0.67 to 1.26) ‡ | 1.01 (0.74 to 1.38) ‡ |
| Eye-tracking |  |  |  |  |  |  |  |  |
| Infant Oriented with Attention response time, ms | 217 | 430 ± 106 | 223 | 431 ± 106 | 254 | 418 ± 96 | 12 (-10 to 34) | 13 (-9 to 36) |
| Visual paired comparison novelty preference score | 239 | 0.60 ± 0.1 | 238 | 0.58 ± 0.1 | 290 | 0.59 ± 0.1 | 0.01 (-0.01 to 0.03) | -0.01 (-0.03 to 0.01) |
| Mean fixation duration, ms | 248 | 330 ± 152 | 244 | 358 ± 154 | 297 | 343 ± 162 | -13 (-45 to 19) | 15 (-17 to 47) |
| **Programmatic Outcomes** |  |  |  |  |  |  |  |  |
| Recovered, no. (%) | 809 | 583 (72.1) | 860 | 638 (74.2) | 896 | 672 (75.0) | -2.9 (-7.1 to 1.3) | -0.8 (-4.9 to 3.3) |
| Remained malnourished, no. (%) | 809 | 164 (20.3) | 860 | 162 (18.8) | 896 | 172 (19.2) | 1.1 (-2.7 to 4.9) | -0.4 (-4.1 to 3.3) |
| Improved to moderate acute malnutrition, no. (%) | 809 | 119 (14.7) | 860 | 114 (13.3) | 896 | 117 (13.1) | 1.6 (-1.7 to 4.9) | 0.2 (-3.0 to 3.4) |
| Remained severely malnourished, no. (%) | 809 | 45 (5.6) | 860 | 48 (5.6) | 896 | 55 (6.1) | 1.1 (-2.7 to 1.7) | -0.5 (-2.7 to 1.7) |
| Died, no. (%) | 809 | 19 (2.3) | 860 | 16 (1.9) | 896 | 13 (1.5) | 0.8 (-0.5 to 2.1) | 0.4 (-0.8 to 1.6) |
| Defaulted, no. (%) | 809 | 43 (5.3) | 860 | 44 (5.1) | 896 | 39 (4.4) | 0.9 (-1.1 to 2.9) | 0.7 (-0.13 to 2.7) |
| **Anthropometric Outcomes** |  |  |  |  |  |  |  |  |
| Rate of weight gain, g/kg/day | 786 | 3.7 ± 4.0 | 836 | 3.8 ± 3.8 | 875 | 4.1 ± 3.6 | -0.3 (-0.7 to 0.0) | -0.3 (-0.6 to 0.1) |
| Rate of mid-upper arm circumference gain, mm/day | 786 | 0.27 ± 0.27 | 836 | 0.26 ± 0.26 | 875 | 0.29 ± 0.27 | -0.02 (-0.04 to 0.01) | -0.03 (-0.05 to 0.00) |
| Rate of length gain, mm/day | 786 | 0.37 ± 0.36 | 836 | 0.36 ± 0.39 | 872 | 0.35 ± 0.33 | 0.02 (-0.01 to 0.06) | 0.01 (-0.02 to 0.05) |
| **Adverse events, first 2 weeks of intervention** |  |  |  |  |  |  |  |  |
| Fever, no. (%) | 785 | 209 (27) | 836 | 217 (26) | 875 | 269 (31) | -4.0 (-8.4 to 0.4) | -5.0 (-9.3 to -0.7) |
| Diarrhea, no. (%) | 784 | 242 (31) | 834 | 247 (30) | 874 | 292 (33) | -2.0 (-6.5 to 2.5) | -3.0 (-7.4 to 1.4) |
| Eating, no. (%) | 784 | 773 (99) | 836 | 819 (98) | 873 | 858 (98) | 1.0 (-0.2 to 2.2) | 0.0 (-1.3 to 1.3) |

<sup>1</sup> Plus-minus values are means ± SD. Significance testing was performed for primary outcomes only. Malawi Developmental Assessment Tool and modified Willatts Problem Solving Assessment. Pairwise comparisons for continuous variables were performed with Student's t-test. Comparisons below P = 0.05 threshold are indicated with superscript letters with values shown in this legend. DHA-HO-RUTF, docosahexaenoic acid added to ready-to-use therapeutic food made with high-oleic acid peanuts; HO-RUTF, high-oleic acid ready-to-use therapeutic food; S-RUTF, standard ready-to-use therapeutic food.
† All values are differences with 95% confidence intervals unless otherwise noted.
‡ This value is the odds ratio with 95% confidence interval.
<sup>a</sup> P = 0.044
<sup>b</sup> P = 0.031
<sup>c</sup> P = 0.047
<sup>d</sup> P = 0.002

**Figure 2.**
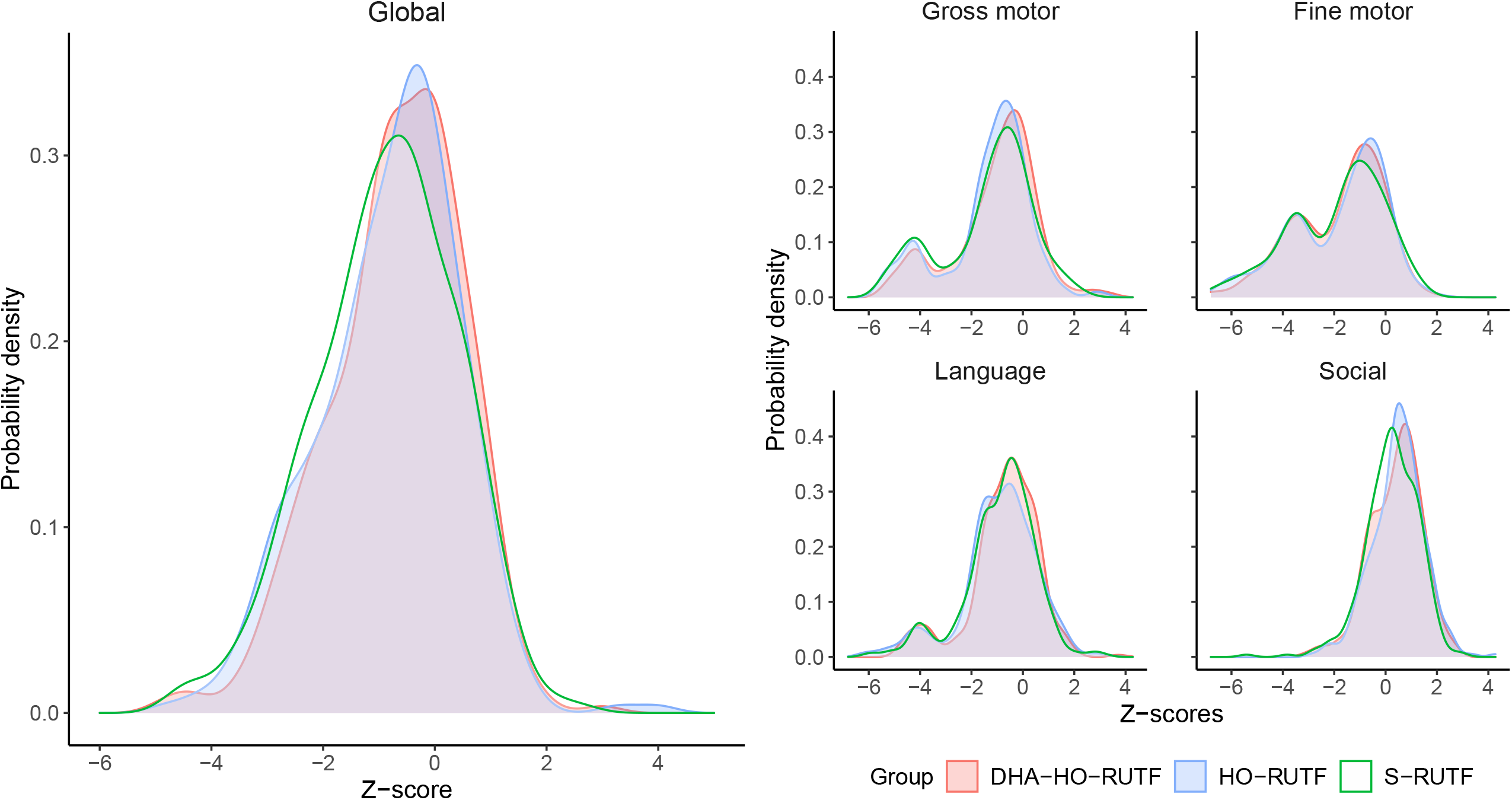
Probability density plots of Malawi Developmental Assessment Tool global and domain z-scores of children receiving RUTF made with high oleic acid peanuts with added docosahexaenoic acid (DHA-HO-RUTF), RUTF made with oleic acid peanuts but without added DHA (HO-RUTF), or standard RUTF (S-RUTF). Probability densities were constructed using kernel density estimation. Global n_*DHA-HO-RUTF*_ = 332, n_*HO-RUTF*_ = 312, n_*S-RUTF*_ = 342. Gross motor domain n_*DHA-HO-RUTF*_ = 331, n_*HO-RUTF*_ = 312, n_*S-RUTF*_ = 342. Fine motor n_*DHA-HO-RUTF*_ = 323, n_*HO- RUTF*_ = 306, n_*S-RUTF*_ = 337. Language n_*DHA-HO-RUTF*_ = 331, n_*HO-RUTF*_ = 312, n_*S-RUTF*_ = 342, n_*DHA-HO-RUTF*_ = 332, n_*HO-RUTF*_ = 311, n_*S-RUTF*_ = 342.

HO-RUTF or DHA-HO-RUTF did not lead to superior PSA scores compared to S-RUTF (Table 3, **Figure 3**). Inability to participate in problem 3 adequately to obtain a score occurred more often than anticipated, and resulted in an underpowered analysis for PSA. Predicted probabilities from the ordinal logistic regression model showed that intention scores were strongly influenced by age and varied most in problem 3 (**Supplementary Figure 3**).

**Figure 3.**
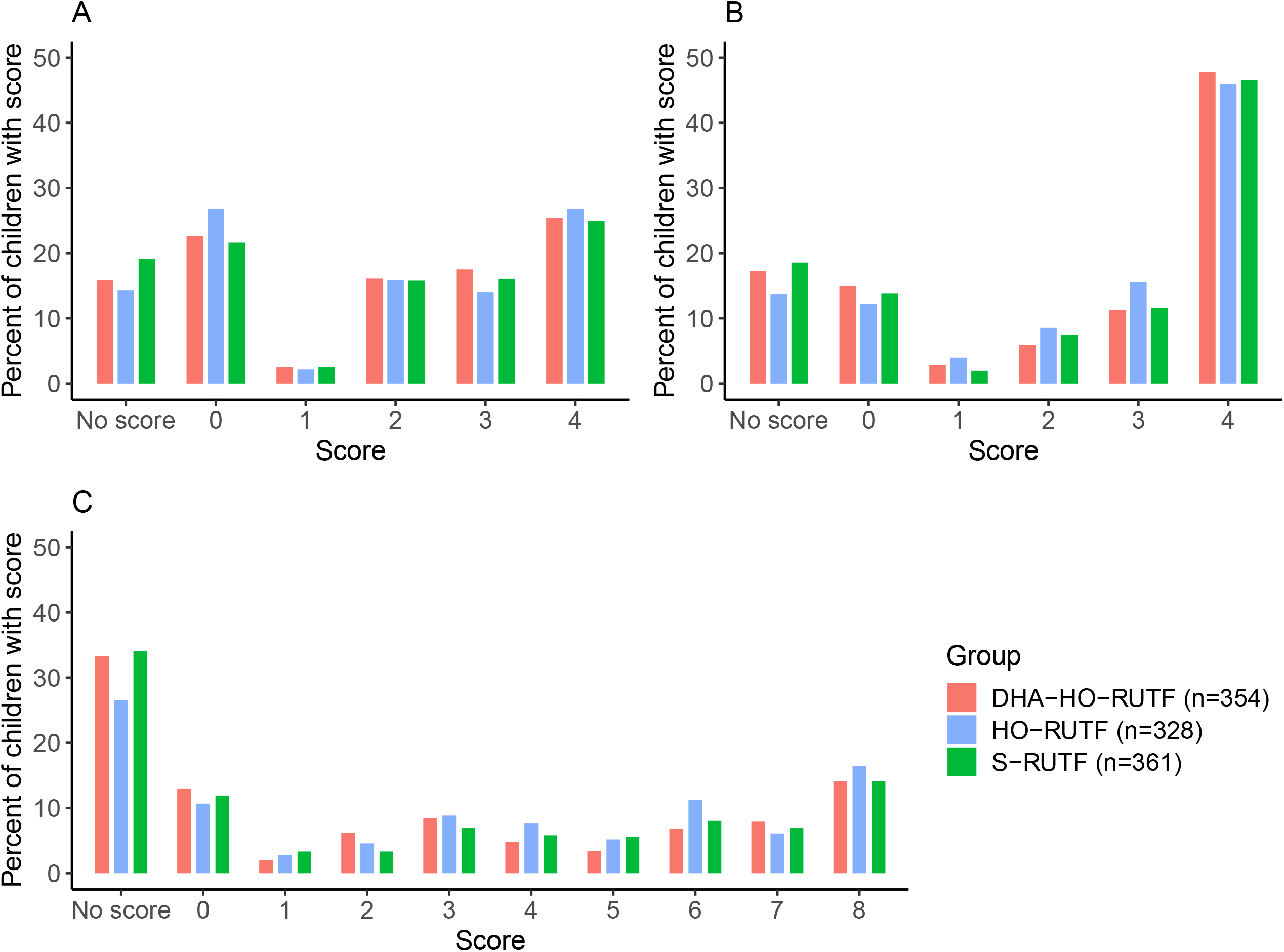
Problem solving assessment intention scores of children receiving RUTF made with high oleic acid peanuts with added docosahexaenoic acid (DHA-HO-RUTF), RUTF made with oleic acid peanuts but without added DHA (HO-RUTF), or standard RUTF (S-RUTF). Percentages of children in each intervention group with each score are shown. Overall, children in each study food had similar intention scores in all three problems; the differences between food groups were not significant. The lowest and highest scores possible in each problem were the most common results among children with scores. Children with “No Score” were unable to engage in the task.

Neither intervention RUTF led to superior eye-tracking results, anthropometric recovery, rates of gain in weight, length, or MUAC compared to S-RUTF (Table 3). Potential adverse events, including reports of fever, diarrhea, food intolerance, and poor appetite, were not higher in the intervention groups compared to S-RUTF, nor were default rates different between groups (Table 3).

Summary statistics for enrollment characteristics and analysis of primary and key secondary outcomes were repeated after excluding those children enrolled during each of the three stock-out periods (271 children excluded, **Supplementary Tables 5-6**). The groups remained balanced at baseline and the point estimates were similar for all primary and secondary outcomes. Exploratory analysis of MDAT results by key subgroups is shown in **Supplementary Figure 4**. Linear and ordinal logistic regression models assessed the effects of DHA and HO in RUTF on MDAT global z-score and PSA intentions scores, respectively, is shown in **Supplementary Table 7**.

Six key plasma PUFA concentrations were selected to describe the biochemical changes induced by HO-RUTF and DHA-HO-RUTF (**Figure 4**). LA and ALA are the primary dietary omega-6 and omega-3 fatty acids respectively, EPA and DHA are bioactive omega-3 fatty acids that affect mood and cognition, and arachidonic acid and docosapentaenoic acid are bioactive omega-6 fatty acids. Consumption of HO-RUTF and DHA-HO-RUTF resulted in greater amounts of plasma ALA, as well as more EPA, while DHA was increased only in the DHA-HO-RUTF group. S-RUTF resulted in more LA than DHA-HO-RUTF, as well as greater amounts of the bioactive omega-6 fatty acids. The full plasma fatty acid concentrations from SAM children fed different RUTFs can be found in **Supplementary Table 8**.

**Figure 4.**
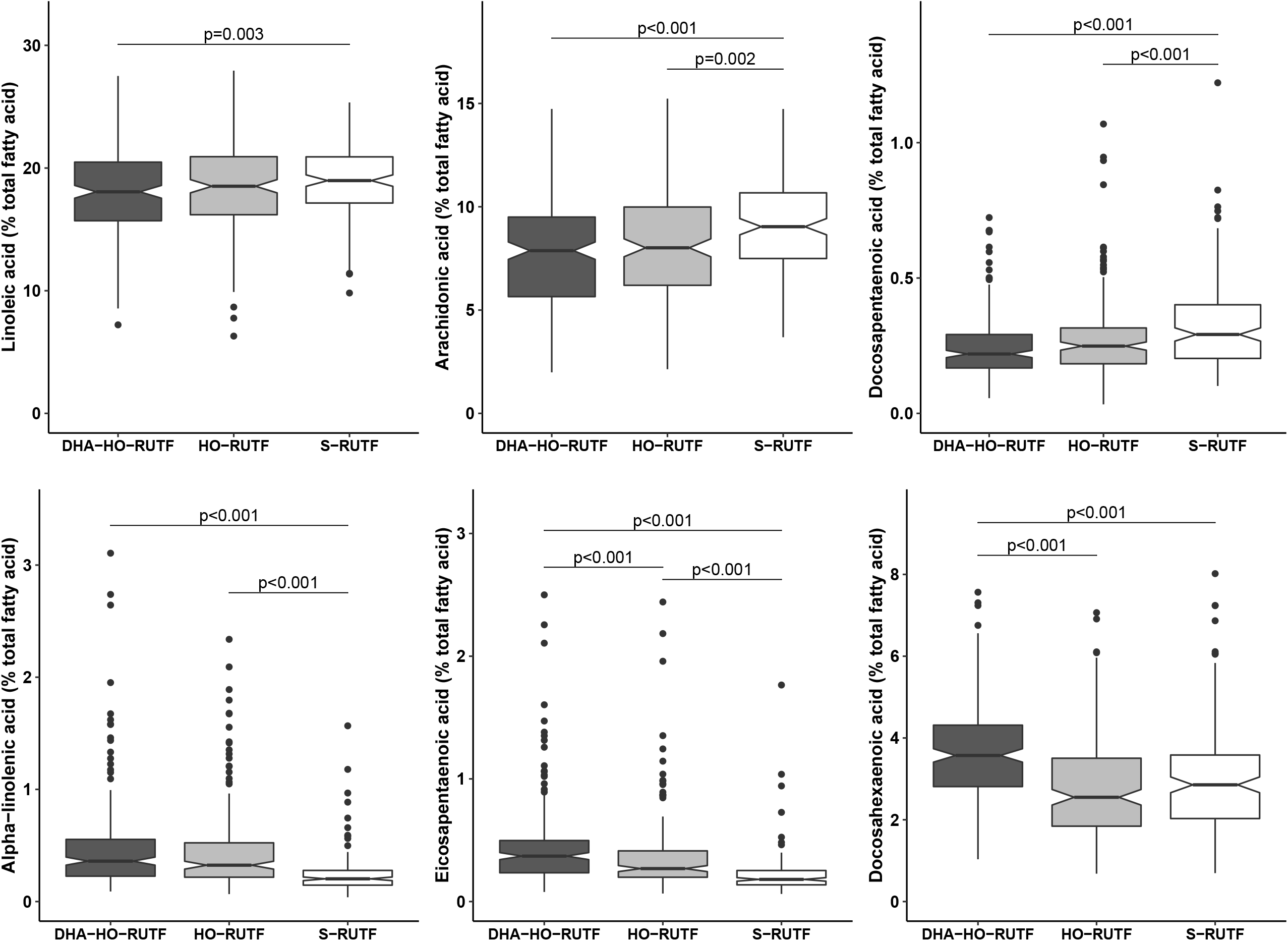
Box and whisker plot of plasma fatty acid content of 6 selected fatty acids in severely malnourished children receiving RUTF made with high oleic acid peanuts with added DHA (DHA-HO-RUTF), RUTF made with high oleic acid peanuts (HO-RUTF), or standard RUTF (S-RUTF). The boxes represent the interquartile range of the distribution with a heavy midline median value. The whiskers extend to 1.5 times the IQR, values outside of these designations are plotted as points. The statistical comparisons were made using a Wilcoxon Rank Sum test, the numbers of samples in each group were DHA-HO-RUTF=208 HO-RUTF=189 S-RUTF=162.

## Discussion

This trial found that SAM children who consumed DHA-HO-RUTF achieved superior global MDAT scores than children who consumed S-RUTF. DHA-HO-RUTF caused a positive shift in the distribution of MDAT scores measured about 6 months after a SAM outcome with no intervening experimental treatment. However, PSA scores were similar in all dietary groups. The expected improvements in plasma fatty acid content were observed; children receiving DHA-HO-RUTF had greater concentrations of DHA. Anthropometric recovery was similar; 72% for DHA-HO-RUTF and 75% for S-RUTF. No adverse effects or negative preferences against DHA-HO-RUTF were observed in this large trial.

This trial demonstrated a clear discrepancy between anthropometric recovery and cognitive recovery in SAM. Children fed S-RUTF show inferior cognitive performance when compared with population norms, an insult of 0.9 MDAT z-scores. This deleterious effect is observed in all domains of the MDAT, except for the social domain. The magnitude of the insult suggests that a frameshift in thinking and approach may be warranted with respect to SAM treatment and recovery. The focus on cognitive recovery must rise to equal importance as anthropometric recovery to enable the child to thrive. The distribution of MDAT scores in children < 1 year of age mirrors a healthy population (Supplementary Figure 2) and offers hope that improvements in treatment may avert lifelong disabilities. The increase seen in MDAT by the inclusion of less LA and more DHA in RUTF is encouraging and actionable. Differences seen *between* dietary groups were not dramatic (Supplemental Figure 1) when compared to the SAM insult. Our results report a functional benefit that extends the importance of our previous biochemical results, indicating that children recovering from SAM require a source of DHA to restore cognition (17,34).

The MDAT was developed in a rural Malawian setting, is fully validated, reliable, and predictive of later intellectual performance. This is why MDAT was chosen was as a primary outcome, and why Malawi was chosen as the study location. MDAT has revealed cognitive deficits in acutely malnourished populations in Malawi and Burkina Faso. In a previous study, MDAT domain scores were reported in 150 Malawian SAM children upon discharge from the hospital (21). All 4 domain scores were remarkably lower among hospitalized SAM children than in our population; differences of -1.7 z-scores seen in gross motor, -1.0 z-scores in fine motor, -0.7 in language, and -3.0 z-scores seen in social. The MUAC of these hospitalized children was 11.5 cm, similar to that in our population, although the clinical status due to infectious complications was likely worse. These findings lead us to speculate that significant improvements in MDAT occur after discharge, especially in the gross motor and social domains, and that maximizing this “cognitive recovery” should be a primary goal in malnutrition programs

The PSA scores did not differ between food groups. PSA problem 3 has been used to demonstrate cognitive differences among healthy infants aged 9-12 mo receiving DHA enriched infant formulas (25). In this study, half of the healthy infants achieved a perfect score on the PSA on all 3 attempts. This is in contrast to our results, wherein only 15% of children achieved a perfect score on any attempt. This suggests the presence of a substantial cognitive deficit at the time of anthropometric recovery. Our PSA was conducted using this very same problem and protocol, and with a sample size increased three-fold, as previously reported, so it is unlikely the failure to detect differences is the result of methodological flaws. Rather, PSA problem 3 was beyond the intellectual capacity of most children tested. Subsequent use of PSA in SAM should explore its timing relative to anthropometry and the potential utility of repeated testing.

Two previous, smaller studies in SAM children were done using blood measures of fatty acid content (18,35). Our findings are consonant with these; increases in DHA were only seen when fish oil was added to the diet, and reduction in dietary LA resulted in greater EPA content.

The food formulations of HO-RUTF and DHA-HO-RUTF were achieved by Project Peanut Butter by ingredient changes, without alterations in the mixing or packaging processes. HO peanuts can be purchased in the major peanut production markets worldwide. DHA is now encapsulated and available from the same ingredient producers as the micronutrient premix. Encapsulation is very durable and obscures the taste of the fish oil exceedingly well, in addition to limiting oxidative degradation. Non-genetically modified, high oleic vegetable oils are available at a cost of about 10% more than traditional vegetable oils. They were developed for purposes of increasing the shelf-life of the oil and have been shown to have a reduced risk of heart disease in adults.

Our study has multiple limitations. The study population did not habitually consume fish, and the positive effect of DHA-HO-RUTF in a fish consuming population might differ. However, most children recovering from SAM worldwide consume little else but RUTF, and RUTF should meet the needs of such children. Our study population developed SAM largely because of food insecurity. Children in whom chronic illness or excessive inflammation precipitated SAM might not realize similar cognitive benefits of DHA-HO-RUTF. The use of different cognitive assessments at single time points limited the ability to interpret the dynamic nature of cognitive recovery. Food stock-outs resulted in imbalanced randomization, potentially introducing bias. Re-analysis of the sample enrolled outside of these stock-outs did not show evidence of bias.

We estimate that during treatment, SAM children were consuming about 240g RUTF/d. For the DHA-HO-RUTF group, DHA intake averaged 173 mg/d or about 0.24%w/w DHA. The global breastmilk DHA reference level is 0.32%w/w.(35) Fat constitutes half the energy in breastmilk and RUTF. The DHA content of DHA-HO-RUTF and breastmilk was similar. Importantly, the effective intervention was to supply RUTF with DHA and with limited LA when recovery food was supplying calories and protein needed to restart a normal or even accelerated trajectory of neurocognitive development. The brain substitutes omega-6 docosapentaenoic acid (DPA6) for DHA when dietary LA is surfeit and omega-3 is limiting. DPA does not support neurocognitive function similarly to DHA, thus our results may point to slow replacement of DHA for DPA in the months after recovery (11).

LA antagonism of omega-3 PUFAs is understood from a molecular and genetic basis (36-38). Large dietary amounts of LA and little ALA given to pregnant animals results in offspring with increased aggression and impulsivity, reduced executive function, and impaired visual function (39). Confusion over the interrelationship of dietary LA and ALA has led to futile attempts to increase circulating/tissue DHA by increasing dietary ALA without reducing LA. While the dietary omega-6 to omega-3 *ratio*, usually cast as [LA]/[ALA], is widely quoted as a parameter defining omega-3 tissue accretion, it is the excess of omega-6 over omega-3, i.e. [LA]-[ALA], that controls DHA availability (40,41). Clinical studies show that additional ALA does not increase circulating or breastmilk DHA (42). Moreover, no amount of supplementary omega-3 EPA increases DHA. To increase DHA, two dietary interventions are well established: lowering dietary LA and increasing dietary DHA.

This study is the first to provide direct evidence that reduction in LA and addition of DHA in RUTF enhances cognition in SAM children. This finding is consonant with a body of scientific evidence that extends over many decades, methodologies, and species. Perhaps the most compelling evidence is found in the composition of breast milk. The need to enhance cognitive recovery in SAM is substantial, even crucial, as this insult affects tens of millions of children worldwide annually. At present, about 85M children worldwide will develop SAM in the first 5 years of their life, a number roughly equal to the estimate of all other causes of childhood developmental delay. Changing the composition of RUTF to reduce the insult of SAM is safe, feasible, and effective. Further research is needed to optimize the amount and duration of DHA supplementation, but in the meantime, there is a clear course of action to help SAM children worldwide: require that RUTF have reduced amounts of LA and include preformed DHA.

## Supporting information

Trial Protocol

## Data Availability

Data described in the manuscript, code book, and analytic code will be made available upon request pending application to the corresponding author and approval.

## Abbreviations used

ALA: α-linolenic acid;
DHA: docosahexaenoic acid;
DHA-HO-RUTF: high oleic acid RUTF with added DHA;
EPA: eicosapentaenoic acid;
HAZ: height-for-age z-score;
HO-RUTF: high oleic acid RUTF;
LA: linoleic acid;
MDAT: Malawi Developmental Assessment Tool;
MUAC: mid-upper arm circumference;
PSA: problems solving assessment;
PUFA: polyunsaturated fatty acid;
RUTF: ready-to-use therapeutic food;
SAM: severe acute malnutrition;
S-RUTF: standard RUTF;
WHZ: weight-for-height z-score

## Acknowledgment

MJM is an associate editor at AJCN. The other authors have no conflict of interest to declare. JTB, MCG, KM, DRW, RC, and MJM designed the research; RC and KM designed the neurocognitive testing and scoring; MN, MG, RL, IHC, KM, and MCG conducted the clinical research, assessed the children’s cognition, and collected the samples; JTB, HGP, and RJSL conducted the basic laboratory work, KS, MCG, JTB, RJSL, RC, and MJM analyzed and interpreted the data; KS and MCG created the figures and graphically analyzed the data; MCG, KS, and MJM wrote the first draft of the manuscript with input from JTB; MJM has primary responsibility for final content. All authors read and approved the final manuscript. The trial was registered at NCT03094247.

We are grateful to the Wiley Companies for the generous gift of AlaskOmega® omega-3 concentrate derived from Alaskan Pollock (Gadus chalcogrammus).

