## Supplementary Tables and Figures for "Low linoleic acid foods with added DHA given to Malawian children with severe acute malnutrition improves cognition: a randomized, triple blinded, controlled clinical trial"

1

0

-1

-2

-3

Malawi Developmental Assessment Tool domain

MDAT z-score

Global

Gross motor

Fine motor

Language

Social

DHA-HO-RUTF

HO-RUTF

S-RUTF

p=0.044

p=0.031

p=0.047

p=0.002

**Supplementary Figure 1*.*** Malawi Developmental Assessment Tool (MDAT) global and domain z-scores of children receiving RUTF made with high oleic acid peanuts with added docosahexaenoic acid (DHA-HO-RUTF), RUTF made with oleic acid peanuts but without added DHA (HO-RUTF), or standard RUTF (S-RUTF). Symbols represent means. Z-scores were compared pairwise using Student’s *t* test and p-values <0.05 are reported with bars overlying comparisons*.* Global n*_DHA-HO-RUTF_* = 332, n*_HO-RUTF_* = 312, n*_S-RUTF_* = 342. Gross motor domain n*_DHA-HO-RUTF_* = 331, n*_HO-RUTF_* = 312, n*_S-RUTF_* = 342. Fine motor n*_DHA-HO-RUTF_* = 323, n*_HO-RUTF_* = 306, n*_S-RUTF_* = 337. Language n*_DHA-HO-RUTF_* = 331, n*_HO-RUTF_* = 312, n*_S-RUTF_* = 342, n*_DHA-HO-RUTF_* = 332, n*_HO-RUTF_* = 311, n*_S-RUTF_* = 342

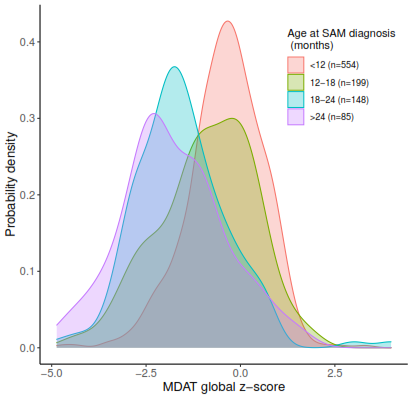

**Supplementary Figure 2.** Probability density plots of Malawi Developmental Assessment Tool (MDAT) global z-scores based on age in months of participant when diagnosed with severe acute malnutrition (SAM). Probability densities were constructed using kernel density estimation.

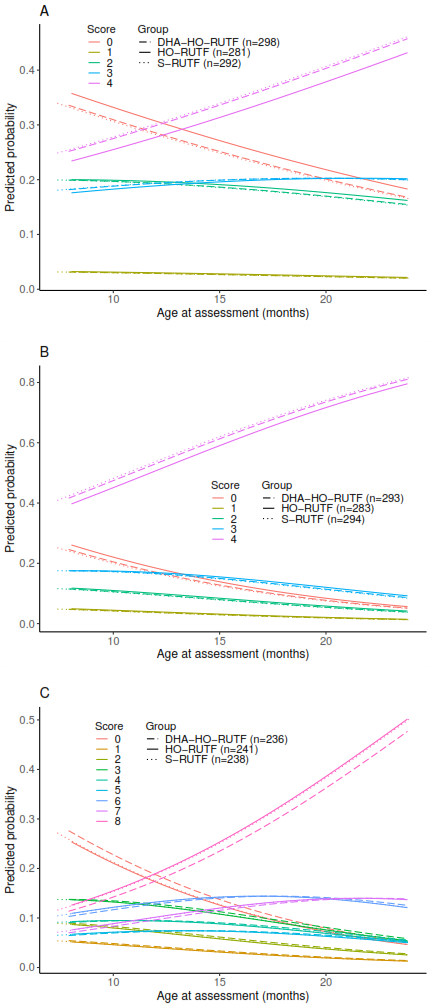

**Supplementary Figure 3.** Predicted probabilities of problem solving assessment (PSA) intention scores and age at assessment of children receiving RUTF made with high oleic acid peanuts with added docosahexaenoic acid (DHA-HO-RUTF), RUTF made with oleic acid peanuts but without added DHA (HO-RUTF), or standard RUTF (S-RUTF). Predicted probabilities generated from the ordinal logistic regression model. Colors represent intention score and line dash denote food group. Plot A is problem 1 where the highest intention score given was 4 and lowest intention score was 0; DHA-HO-RUTF n=298; HO-RUTF n=281; S-RUTF n=292. Plot B is problem 2 where the highest intention score given was 4 and lowest intention score was 0; DHA-HO-RUTF n=293; HO-RUTF n=283; S-RUTF n=294. Plot C is problem 3 where the highest intention score was 8 and lowest was 0; DHA-HO-RUTF n=236; HO-RUTF n=241; S-RUTF n=238.

All (N = 674)

Marasmus (N = 468)

Kwashiorkor (N = 206)

HAZ ≤ -2 (N = 574)

HAZ > -2 (N = 100)

Age* < 12 mo (N = 372)

Age* ≥ 12 mo (N = 302)

Female (N = 411)

Male (N = 263)

Recovered (N = 525)

Improved to MAM (N = 108)

Remained SAM (N = 41)

-1.0

-0.5

0.0

0.5

1.0

Z-score difference (95% CI)

Favors S-RUTF

Favors DHA-HO-RUTF

**Supplementary Figure 4.** Subgroup differences in Malawi Developmental Assessment Tool Global z-score between of children receiving RUTF made with high oleic acid peanuts with added docosahexaenoic acid (DHA-HO-RUTF) and standard RUTF (S-RUTF). Global z-score differences calculated by subtracting z-score means in S-RUTF group from those in DHA-HO-RUTF group. HAZ, height-for-age z-score; MAM, moderate acute malnutrition; MDAT, Malawi Developmental Assessment Tool; SAM, severe acute malnutrition. Kwashiorkor defined by presence of bilateral pedal pitting edema. Marasmus defined by either WHZ < -3 or MUAC < 11.5 cm.

*Age at time of episode of severe acute malnutrition.

**Supplementary Table 1.** Nutrient composition of study ready-to-use therapeutic foods per 100g of RUTF

| Nutrient | DHA-HO-RUTF | HO-RUTF | S-RUTF | Specification^1^ |
| --- | --- | --- | --- | --- |
| Energy, kcal | 532 | 531 | 541 | 520-550 |
| Protein, % TE | 11.4 | 11.5 | 11.6 | 10-12 |
| Lipids, % TE | 51.1 | 50.0 | 54.4 | 45-60 |
| omega-6 fatty acids, % TE | 5.0 | 5.0 | 10.0 | 3-10 |
| omega-3 fatty acids, % TE | 4.0 | 3.0 | 1.0 | 0.3-2.5 |
| Potassium, mg | 1271.0 | 1271.0 | 1229.7 | 1,100-1,400 |
| Calcium, mg | 469.9 | 469.9 | 488.1 | 300-600 |
| Phosphorus, mg | 380.4 | 380.4 | 374.8 | 300-600 |
| Magnesium, mg | 85.3 | 85.3 | 80.7 | 80-140 |
| Iron, mg | 12.5 | 12.5 | 12.4 | 10-14 |
| Zinc, mg | 11.8 | 11.8 | 11.9 | 11-14 |
| Copper, mg | 1.8 | 1.8 | 1.8 | 1.4-1.8 |
| Selenium, µg | 31.6 | 31.6 | 31.4 | 20-40 |
| Iodine, µg | 79.7 | 79.7 | 79.7 | 70-140 |
| Vitamin A, mg RE | 1.7 | 1.7 | 1.7 | 0.8-1.1 |
| Thiamin, mg | 0.7 | 0.7 | 0.7 | ≥0.5 |
| Riboflavin, mg | 4.2 | 4.2 | 4.1 | ≥1.6 |
| Niacin, mg | 5.6 | 5.6 | 5.6 | ≥5 |
| Pantothenic acid, mg | 4.2 | 4.2 | 4.1 | ≥3 |
| Vitamin B6, mg | 0.7 | 0.7 | 0.7 | ≥0.6 |
| Biotin, µg | 65.5 | 65.5 | 65.5 | ≥60 |
| Folate, µg | 278.9 | 278.9 | 277.7 | ≥200 |
| Vitamin B12, µg | 2.8 | 2.8 | 2.9 | ≥1.6 |
| Vitamin C, mg | 54.2 | 54.2 | 54.5 | ≥50 |
| Vitamin D, µg | 15.9 | 15.9 | 15.9 | 15-20 |
| Vitamin E, mg | 20.0 | 20.0 | 20.0 | ≥20 |
| Vitamin K1, µg | 20.7 | 20.7 | 20.7 | 15-30 |

^1^Joint Statement on Community-Based Management of Severe Acute Malnutrition by the World Health Organization, the World Food Programme, the United Nations System Standing Committee on Nutrition and the United Nations Children’s Fund, 2007. DHA-HO-RUTF, docosahexaenoic acid added to ready-to-use therapeutic food made with high oleic acid peanuts; HO-RUTF, RUTF made with high oleic acid peanuts; RE, retinol equivalents; S-RUTF, standard RUTF; TE, total energy.

**Supplementary Table 2.** Amino acids provided by the study ready-to-use therapeutic foods^1^

| Amino Acid | DHA-HO-RUTF | HO-RUTF | S-RUTF | Requirement for catch-up weight gain of 10 g/kg/d^*^ |
| --- | --- | --- | --- | --- |
| Histidine, mg | 26 | 26 | 26 | 23 |
| Isoleucine, mg | 49 | 49 | 51 | 34 |
| Leucine, mg | 83 | 83 | 85 | 70 |
| Lysine, mg | 61 | 61 | 63 | 65 |
| Sulfur amino acids^†^, mg | 31 | 31 | 31 | 31 |
| Phenylalanine + tyrosine, mg | 94 | 94 | 96 | 63 |
| Threonine, mg | 41 | 41 | 41 | 37 |
| Tryptophan, mg | 12 | 12 | 12 | 10 |
| Valine, mg | 56 | 56 | 57 | 46 |

^1^ Amino acid expressed mg /g protein. DHA-HO-RUTF, docosahexaenoic acid added to ready-to-use therapeutic food made with high oleic acid peanuts; HO-RUTF, RUTF made with high oleic acid peanuts; S-RUTF, standard RUTF.

^*^ Target protein requirement to achieve a catch-up weight gain of 10 g/kg/d was calculated by considering a compositional weight gain of 73:27, lean/fat equivalent to 14% protein and 27% fat, 14% deposited tissue adjusted for a 70% efficiency of utilization, and a safe level of maintenance at 1.24 x 0.66 g/kg/d = 0.82, with 0.66 g/kg/d being the adult maintenance protein needs (<http://www.fao.org/3/CA2487EN/ca2487en.pdf>).

^†^ Sulfur amino acids = (methionine + cysteine)

|  | DHA-HO-RUTF | HO-RUTF | S-RUTF |
| --- | --- | --- | --- |
|  | (N = 332) | (N = 312) | (N = 342) |
| **Enrollment and nutritional outcomes** |  |  |  |
| Age, mo, median (IQR) | 11.1 (7.3 - 16.8) | 10.2 (7.1 - 18.0) | 10.7 (7.7 – 18.0) |
| Female sex, n (%) | 203 (60.2) | 182 (58.3) | 212 (61.1) |
| Edematous, n (%) | 110 (32.6) | 93 (29.8) | 101 (29.1) |
| Mid-upper arm circumference, cm | 11.6 ± 1.1 | 11.4 ± 1.1 | 11.5 ± 1.0 |
| MUAC < 11.5 cm, n (%) | 231 (68.5) | 212 (67.9) | 246 (70.9) |
| Weight-for-height z-score | -1.7 ± 1.1 | -1.8 ± 1.1 | -1.9 ± 1.0 |
| WHZ < -3, n (%) | 45 (13.4) | 51 (16.3) | 53 (15.3) |
| Height-for-age z-score | -3.2 ± 1.3 | -3.3 ± 1.5 | -3.3 ± 1.4 |
| HAZ ≤ -2, n (%) | 288 (85.5) | 266 (85.3) | 294 (84.7) |
| HAZ ≤ -3, n (%) | 193 (57.3) | 181 (58.0) | 185 (53.3) |
| Recovered, no. (%) | 260 (77.2) | 243 (77.9) | 271 (78.1) |
| Improved to MAM, no. (%) | 59 (17.5) | 52 (16.7) | 52 (15.0) |
| Remained SAM, no. (%) | 18 (5.3) | 17 (5.4) | 24 (6.9) |
| Duration of treatment, weeks, median (IQR) | 6 (4 - 12) | 6 (4 - 12) | 6 (4 - 12) |
| **Demographics at time of Malawi Developmental Assessment Tool test** |  |  |  |
| Age at MDAT, mo, median (IQR) | 18.9 (15.5 – 24.4) | 17.7 (15.3 - 25.4) | 18.9 (15.6 – 25.4) |
| Time between outcome and MDAT, mo | 5.8± 0.5 | 5.8 ± 0.5 | 5.9 ± 0.6 |
| Primary caregiver mother, no. (%) | 311 (92.3) | 289 (92.6) | 324 (93.4) |
| Primary caregiver at least some primary school education, no. (%) | 248 (73.6) | 232 (74.4) | 270 (77.8) |
| Secondary caregiver at least some primary school education, no. (%) | 233 (69.8) | 218 (71.5) | 257 (75.6) |
| Number of siblings, median (IQR) | 2 (1 - 3) | 2 (1 - 4) | 2 (0 - 4) |

**Supplementary Table 3.** Enrollment characteristics, nutritional outcomes, and characteristics at time of testing of the children who underwent Malawi Developmental Assessment Tool testing^1^

^1^ Plus-minus values are means ± SD. DHA-HO-RUTF, docosahexaenoic acid added to ready-to-use therapeutic food made with high oleic acid peanuts; HAZ, height-for-age z-score; HO-RUTF, RUTF with high oleic acid peanuts; MAM, moderate acute malnutrition; MDAT, Malawi Developmental Assessment Tool; MUAC, mid-upper arm circumference; SAM, severe acute malnutrition; WHZ, weight-for-height z-score.

Summary

Median age at time of MDAT was similar between groups, and mean duration between SAM outcome and MDAT was similar, 5.8 – 5.9 months. 10 (1.0%) were excluded from analysis, 9 participants due to outlier MDAT global z-scores < -5 and 1 who was unable to be scored for > 2 domains. Several children were unable to be scored for one or more sub-domains due to inadequate item completion: one child had no gross motor score (DHA-HO-RUTF), twenty did not have fine motor scores (nine DHA-HO-RUTF, six HO-RUTF, five S-RUTF), one did not have a language score (DHA-HO-RUTF), and one did not have a social score (HO-RUTF).

|  | DHA-HO-RUTF | HO-RUTF | S-RUTF |
| --- | --- | --- | --- |
|  | (N = 354) | (N = 328) | (N = 361) |
| **Enrollment characteristics and nutritional outcomes** |  |  |  |
| Age, mo, median (IQR) | 10.5 (7.7 – 15.1) | 10.0 (7.4 – 14.8) | 10.6 (7.4 – 14.2) |
| Female sex, n (%) | 212 (59.9) | 183 (55.8) | 219 (60.7) |
| Edematous, n (%) | 102 (28.8) | 98 (29.9) | 91 (25.2) |
| Mid-upper arm circumference, cm | 11.4 ± 1.0 | 11.4 ± 1.0 | 11.3 ± 0.8 |
| MUAC < 11.5, n (%) | 254 (71.8) | 235 (71.6) | 274 (75.9) |
| Weight-for-height z-score | -1.9 ± 1.1 | -1.9 ± 1.1 | -2.0 ± 1.0 |
| WHZ < -3, n (%) | 60 (16.9) | 46 (14.0) | 59 (16.3) |
| Height-for-age z-score | -3.1 ± 1.4 | -3.3 ± 1.5 | -3.2 ± 1.4 |
| HAZ ≤ -2, n (%) | 283 (79.9) | 277 (84.5) | 311 (86.1) |
| HAZ ≤ -3, n (%) | 185 (52.3) | 183 (55.8) | 201 (55.7) |
| Recovered, no. (%) | 268 (75.7) | 252 (76.8) | 267 (74.0) |
| Improved to MAM, no. (%) | 66 (18.7) | 52 (15.9) | 68 (18.8) |
| Remained SAM, no. (%) | 20 (5.6) | 24 (7.3) | 26 (7.2) |
| Duration of treatment, weeks, median (IQR) | 6 (4 - 12) | 6 (4 - 12) | 6 (4 - 12) |
| **Demographics at time of problem solving assessment** |  |  |  |
| Age at PSA, mo, median (IQR) | 12.2 (9.8 – 16.9) | 12.0 (9.6 – 16.4) | 12.1 (9.6 – 15.7) |
| Time between outcome and PSA, weeks | 0.5 ± 1.1 | 0.6 ± 1.1 | 0.6 ± 1.1 |
| Primary caregiver mother, no./total no. (%) | 325/351 (92.6) | 305/328 (93.0) | 336/359 (93.6) |
| Number of siblings, median (IQR) | 2 (0 - 3) | 2 (0 - 4) | 2 (1 - 4) |

**Supplementary Table 4.** Enrollment characteristics, nutritional outcomes, and characteristics at time of assessment of the children who underwent modified Willatts problem solving assessments^1^

^1^Plus-minus values are means ± SD. HAZ, height-for-age z-score; MAM, moderate acute malnutrition; MDAT, Malawi Developmental Assessment Test; MUAC, mid-upper arm circumference; SAM, severe acute malnutrition; WHZ, weight-for-height z-score.

Summary

Median age at time of problem solving assessment was similar between groups, as was mean time between SAM outcome and assessment, 0.5 – 0.6 weeks. A minority of children were unable to engage in each problem sufficiently for scoring: for problems 1 and 2, 14-19% had no score, while for problem 3, 27-34% had no score (Figure 3).

|  | DHA-HO-RUTF | HO-RUTF | S-RUTF |
| --- | --- | --- | --- |
|  | (N = 774) | (N = 764) | (N = 756) |
| Female sex, no. (%) | 434 (56.1) | 417 (54.6) | 435 (57.5) |
| Age, mo, median (IQR) | 11.8 (7.9 – 20.3) | 12.1 (7.7 - 21.1) | 12.0 (7.8 - 21.1) |
| Edematous, no. (%) | 257 (33.2) | 263 (34.4) | 230 (30.4) |
| Mid-upper-arm circumference, cm | 11.5 ± 1.2 | 11.6 ± 1.2 | 11.6 ± 1.2 |
| MUAC < 11.5 cm, no. (%) | 516 (66.7) | 486 (63.6) | 505 (66.8) |
| Weight-for-height z-score | -1.9 ± 1.2 | -1.9 ± 1.3 | -2.0 ± 1.2 |
| WHZ ≤ -3, no. (%) | 156 (20.2) | 161 (21.1) | 165 (21.8) |
| Height-for-age z-score | -3.3 ± 1.5 | -3.3 ± 1.5 | -3.2 ± 1.4 |
| HAZ ≤ -2, no./total no. (%) | 639/774 (82.6) | 645/764 (84.4) | 637/756 (84.3) |
| Fever in past 2 weeks, no./total no. (%) | 441/770 (57.0) | 457/763 (59.8) | 464/753 (61.4) |
| Diarrhea in past 2 weeks, no./total no. (%) | 428/770 (55.3) | 420/763 (55.0) | 405/751 (53.6) |
| Child breastfed, no./total no. (%) | 502/773 (64.9) | 506/762 (66.4) | 498/753 (66.1) |
| HIV-seropositive, no./total no. tested (%) | 18/254 (7.1) | 22/271 (8.1) | 20/262 (7.6) |
| Mother alive, no./total no. (%) | 748/772 (96.9) | 734/764 (96.1) | 737/753 (97.9) |
| Number of siblings, median (IQR) | 2 (0-3) | 2 (1-3) | 2 (0-3) |
| Thatch roof, no./total no. (%) | 583/769 (75.8) | 584/763 (76.5) | 574/750 (76.5) |
| Radio in home, no./total no. (%) | 169/772 (21.9) | 180/758 (23.7) | 146/754 (19.4) |
| Clean water source, no./total no. (%) | 728/772 (94.3) | 704/762 (92.4) | 702/755 (93.0) |

**Supplementary Table 5.** Enrollment characteristics of children enrolled outside of the three time periods of study food stock-out^1^

^1^ Plus-minus values are means ± SD. DHA-HO-RUTF, docosahexaenoic acid added to ready-to-use therapeutic food made with high-oleic acid peanuts; HAZ, height-for-age z-score; HIV, human immunodeficiency virus; HO-RUTF, RUTF made with high oleic acid peanuts; MUAC, mid-upper arm circumference; WHZ, weight-for-height z-score.

**Supplementary Table 6.** Cognitive, programmatic, and growth outcomes, and adverse events of children enrolled outside of the three time periods of study food stock-outs^1^

|  | DHA-HO-RUTF | | HO-RUTF | | S-RUTF | | DHA-HO-RUTF vs. S-RUTF | HO-RUTF vs.  S-RUTF |
| --- | --- | --- | --- | --- | --- | --- | --- | --- |
|  | N |  | N |  | N |  | Comparison† | Comparison† |
| **Cognitive Outcomes** |  |  |  |  |  |  |  |  |
| MDAT z-scores |  |  |  |  |  |  |  |  |
| Global | 312 | -0.65 ± 1.17 | 312 | -0.77 ± 1.25 | 288 | -0.85 ± 1.26 | 0.19 (0.00 to 0.39) | 0.07 (-0.13 to 0.28) |
| Gross motor domain | 311 | -1.07 ± 1.66 | 312 | -1.32 ± 1.66 | 288 | -1.34 ± 1.77 | 0.27 (0.00 to 0.54) | 0.02 (-0.26 to 0.30) |
| Fine motor domain | 303 | -1.77 ± 1.70 | 306 | -1.81 ± 1.81 | 283 | -1.87 ± 1.83 | 0.10 (-0.19 to 0.39) | 0.06 (-0.24 to 0.35) |
| Language domain | 311 | -0.69 ± 1.33 | 312 | -0.93 ± 1.53 | 288 | -0.89 ± 1.41 | 0.20 (-0.02 to 0.42) | -0.03 (-0.27 to 0.21) |
| Social domain | 312 | 0.42 ± 1.00 | 311 | 0.51 ± 0.98 | 288 | 0.27 ± 1.06 | 0.15 (-0.02 to 0.31) | 0.24 (0.08 to 0.41) |
| PSA intention scores |  |  |  |  |  |  |  |  |
| Problem 1, no. (%) with perfect score | 280 | 86 (31) | 265 | 83 (31) | 243 | 75 (31) | 0.95 (0.70 to 1.30)‡ | 0.85 (0.62 to 1.16)‡ |
| Problem 2, no. (%) with perfect score | 275 | 157 (57) | 265 | 145 (55) | 244 | 141 (58) | 0.92 (0.65 to 1.29)‡ | 0.88 (0.63 to 1.23)‡ |
| Problem 3, no. (%) with perfect score | 224 | 46 (21) | 226 | 52 (23) | 200 | 45 (23) | 0.87 (0.62 to 1.23)‡ | 1.04 (0.74 to 1.45)‡ |
| Eye-tracking |  |  |  |  |  |  |  |  |
| Infant Oriented with Attention response time, ms | 205 | 430 ± 107 | 211 | 434 ± 107 | 209 | 417 ± 97 | 13 (-12 to 37) | 17 (-3 to 37) |
| Visual paired comparison novelty preference score | 224 | 0.60 ± 0.1 | 226 | 0.58 ± 0.1 | 237 | 0.59 ± 0.1 | 0.01 (-0.01 to 0.03) | -0.01 (-0.02 to 0.01) |
| Mean fixation duration, ms | 232 | 331 ± 152 | 232 | 354 ± 146 | 243 | 348 ± 165 | -16 (-50 to 18) | 7 (-28 to 41) |
| **Programmatic Outcomes** |  |  |  |  |  |  |  |  |
| Recovered, no. (%) | 774 | 551 (71.2) | 764 | 570 (74.6) | 756 | 558 (73.8) | -2.6 (-7.1 to 1.9) | 0.8 (-3.6 to 5.2) |
| Remained malnourished, no. (%) | 774 | 162 (20.9) | 764 | 145 (19.0) | 756 | 161 (21.3) | -0.4 (-4.5 to 3.7) | -2.3 (-6.3 to 1.7) |
| Improved to moderate acute malnutrition, no. (%) | 774 | 117 (15.1) | 764 | 100 (13.1) | 756 | 111 (14.7) | 0.4 (-3.2 to 4.0) | -1.6 (-5.1 to 1.9) |
| Remained severely malnourished, no. (%) | 774 | 45 (5.8) | 764 | 45 (5.9) | 756 | 50 (6.6) | -0.8 (-3.2 to 1.6) | -0.7 (-3.1 to 1.7) |
| Died, no. (%) | 774 | 19 (2.5) | 764 | 16 (2.1) | 756 | 11 (1.5) | 1.0 (-0.4 to 2.4) | 0.6 (-0.7 to 1.9) |
| Defaulted, no. (%) | 774 | 42 (5.4) | 764 | 33 (4.3) | 756 | 25 (3.3) |  |  |
| Anthropometric Outcomes |  |  |  |  |  |  |  |  |
| Rate of weight gain, g/kg/day | 751 | 3.7 ± 4.0 | 745 | 3.8 ± 3.8 | 740 | 3.9 ± 3.6 | -0.3 (-0.7 to 0.1) | -0.2 (-0.5 to 0.2) |
| Rate of mid-upper arm circumference gain, mm/day | 751 | 0.27 ± 0.27 | 745 | 0.25 ± 0.26 | 740 | 0.28 ± 0.27 | -0.01 (-0.04 to 0.02) | -0.02 (-0.05 to 0.01) |
| Rate of length gain, mm/day | 751 | 0.37 ± 0.36 | 745 | 0.35 ± 0.39 | 738 | 0.35 ± 0.33 | 0.02 (-0.01 to 0.06) | 0.00 (-0.04 to 0.04) |
| **Adverse events, first 2 weeks of intervention** |  |  |  |  |  |  |  |  |
| Fever, no. (%) | 771 | 202 (26) | 763 | 185 (24) | 755 | 227 (30) | -4.0 (-8.5 to 0.5) | -6.0 (-10.5 to -1.5) |
| Diarrhea, no. (%) | 772 | 236 (31) | 763 | 214 (28) | 755 | 245 (32) | -1.0 (-5.7 to 3.7) | -4.0 (-8.6 to 0.6) |
| Eating well, no. (%) | 773 | 738 (95) | 761 | 730 (96) | 751 | 726 (97) | -2.0 (-4.0 to 0.0) | -1.0 (-2.9 to 0.9) |

^1^ Plus-minus values are means ± SD. Significance testing was performed for primary outcomes only. Malawi Developmental Assessment Tool and modified Willatts Problem Solving Assessment. DHA-HO-RUTF, docosahexaenoic acid added to ready-to-use therapeutic food made with heigh-oleic acid peanuts; HO-RUTF, high-oleic acid ready-to-use therapeutic food; S-RUTF, standard ready-to-use therapeutic food.

† All values are differences with 95% confidence intervals unless otherwise noted.

‡ This value is the odds ratio with 95% confidence interval.

|  |  | DHA vs. no DHA | HO vs. no HO |
| --- | --- | --- | --- |
| **Outcome** | N | Comparison (95% CI)† | Comparison (95% CI)† |
| **MDAT** |  |  |  |
| Global z-score | 986 | 0.11 (-0.8 to 0.30)‡ | 0.08 (-0.11 to 0.27)‡ |
| **Problem solving assessment** |  |  |  |
| Problem 1 intention score | 871 | 1.11 (0.83 to 1.49) | 0.89 (0.66 to 1.19) |
| Problem 2 intention score | 870 | 1.10 (0.80 to 1.51) | 0.88 (0.64 to 1.21) |
| Problem 3 intention score | 715 | 0.91 (0.66 to 1.24) | 1.01 (0.74 to 1.38) |

**Supplementary Table 7.** The effects of derived variables for exposure to docosahexaenoic acid- containing ready-to-use therapeutic food (DHA) or RUTF produced with high oleic acid peanuts (HO) on MDAT global-score and modified Willatts problems solving assessment intention scores.

Derived variables were produced by dummy coding for presence or absence of each exposure. Effects of docosahexaenoic acid (DHA) in RUTF and use high oleic peanuts (HO) in RUTF on MDAT global z-score were assessed using linear regression. Effects of DHA in RUTF and use of HO peanuts in RUTF on modified Willatts problems solving assessment intention scores were assessed using ordinal logistic regression. DHA, docosahexaenoic acid; HO, high oleic acid peanuts used to produce ready-to-use therapeutic food; MDAT, Malawi Developmental Assessment Tool; RUTF, ready-to-use therapeutic food.

† All values are odds ratios with 95% confidence intervals unless otherwise indicated.

‡ This value is the beta coefficient with 95% confidence interval.

**Supplementary Table 8.** Fatty acid concentrations in plasma from SAM children fed different PUFA diets^1^

| Common name | Shorthand Designation | DHA-HO-RUTF  n = 208 | HO-RUTF  n = 189 | S-RUTF  n = 162 |
| --- | --- | --- | --- | --- |
| Myristic Acid | 14:0 | 0.76 (0.45-1.14) | 0.81 (0.45-1.19) | 0.67 (0.39-1.08) |
| Palmitic Acid | 16:0 | 28.08 (26.37-29.81) | 28.18 (26.39-29.91) | 27.91 (26.47-29.93) |
| Hypogeic Acid | 16:1 n-9 | 0.09 (0.07-0.12) | 0.10 (0.07-0.14) | 0.09 (0.07-0.13) |
| Palmitoleic Acid | 16:1 n-7 | 0.32 (0.21-0.54) | 0.33 (0.22-0.57) | 0.35 (0.21-0.52) |
| Stearic Acid | 18:0 | 18.05 (15.17-24.30) | 18.75 (15.27-24.54) | 17.17 (14.49-21.04)* |
| Oleic Acid | 18:1 n-9 | 13.07 (10.13-16.81) | 11.98 (9.77-15.99) | 12.78 (10.31-16.63) |
| Vaccenic Acid | 18:1 n-7 | 1.22 (0.98-1.47) | 1.17 (0.96-1.48) | 1.29 (0.99-1.51) |
| Linoleic Acid | 18:2 n-6 | 18.06 (15.70-20.48) | 18.52 (16.19-20.9) | 18.98 (17.15-20.91) |
| γ-Linoleic Acid | 18:3 n-6 | 0.04 (0.03-0.05) | 0.04 (0.03-0.06) | 0.04 (0.03-0.06) |
| α-Linolenic Acid | 18:3 n-3 | 0.36 (0.23-0.55) | 0.32 (0.22-0.52) | 0.20 (0.15-0.28)* |
| Arachidic Acid | 20:0 | 0.23 (0.19-0.27) | 0.24 (0.20-0.29) | 0.23 (0.19-0.29) |
| Eicosenoic Acid | 20:1 n-9 | 0.15 (0.11-0.20) | 0.15 (0.11-0.19) | 0.17 (0.13-0.21) |
| Eicosadienioic Acid | 20:2 n-6 | 0.32 (0.25-0.38) | 0.33 (0.26-0.40) | 0.32 (0.26-0.38 |
| Mead Acid | 20:3 n-9 | 0.04 (0.03-0.08) | 0.05 (0.03-0.10) | 0.06 (0.03-0.11) |
| Sciadonic acid | 5,11,14-20:3 | 0.08 (0.07-0.11) | 0.10 (0.08-0.13) | 0.11 (0.08-0.14) |
| Dihomo-γ-linolenic Acid | 8,11,14-20:3 | 1.59 (1.29-1.92)* | 1.68 (1.32-2.08)* | 1.77 (1.46-2.15) |
| Arachidonic acid | 20:4 n-6 | 7.87 (5.65-9.50) | 8.01 (6.20-9.99) | 9.04 (7.50-10.67)* |
| Behenic Acid | 22:0 | 0.32 (0.21-0.43) | 0.31 (0.20-0.47) | 0.36 (0.24-0.49) |
| Eicosapentaenoic acid EPA | 20:5 n-3 | 0.37 (0.24-0.50)* | 0.27 (0.20-0.41)* | 0.18 (0.14-0.25) |
| Adrenic Acid | 22:4 n-6 | 0.24 (0.17-0.31) | 0.28 (0.20-0.39) | 0.37 (0.25-0.47)* |
| Lignoceric Acid | 24:0 | 0.22 (0.12-0.32) | 0.20 (0.09-0.35) | 0.26 (0.12-0.45) |
| Docosapentaenoic Acid | 22:5 n-6 | 0.22 (0.17-0.29) | 0.25 (0.18-0.32) | 0.29 (0.20-0.40) |
| Nervonic Acid | 24:1 n-9 | 0.47 (0.28-0.65) | 0.45 (0.23-0.62) | 0.52 (0.32-0.67) |
| Docosapentaenoic Acid | 22:5 n-3 | 0.47 (0.37-0.61) | 0.54 (0.37-0.72) | 0.45 (0.34-0.59) |
| Docosahexaenoic Acid (DHA) | 22:6 n-3 | 3.57 (2.81-4.31)* | 2.55 (1.84-3.51) | 2.85 (2.03-3.58) |

^1^Values expressed in %wt./total fatty acids as median (25^th^%tile-75%tile)

*Values in differ significantly from others in their row, as tested by Kruskal-Wallis Test.

Abbreviations: DHA-HO-RUTF, docosahexaenoic acid added to ready-to-use therapeutic food made with high oleic acid peanuts; HO-RUTF, RUTF made with high oleic acid peanuts; S-RUTF, standard RUTF.
