## Supplementary material for "Low linoleic acid foods with added DHA given to Malawian children with severe acute malnutrition improves cognition: a randomized, triple blinded, controlled clinical trial": Trial Protocol

| TRIAL FULL TITLE | Low linoleic acid foods with added DHA given to Malawian children with severe acute malnutrition improves cognition: a randomized, triple blinded, controlled clinical trial |
| --- | --- |
| TRIAL PRINCIPAL INVESTIGATOR | Mark Manary, MD  Helene B Roberson Professor of Pediatrics, Washington University in St. Louis |
| INVESTIGATORS | Kenneth Maleta MBBS  Department of Community Health, College of Medicine, University of Malawi  J Thomas Brenna PhD  Department of Pediatrics, Dell Medical School,  University of Texas at Austin  Richard Lane Canfield PhD  Department of Psychology, Cornell University |
| INSTITUTIONS | Washington University in St. Louis  University of Texas, Austin  College of Medicine, University of Malawi  Cornell University |
| AUTHORS | Kevin Stephenson, Sophia Agapova, Meghan Callaghan-Gillespie, Donna Wegner, J. Thomas Brenna, Scott Lacombe, Reginald Lee, Richard Canfield, Kenneth Maleta, Mark J. Manary |
| FUNDING | Unorthodox Philanthropy  Open Philanthropy |

*The funding agencies have played no role in the design or conduct of this study and do not retain any ownership of the data or other materials collected during the study. There shall also be no input from the funders about the data analysis, manuscript, or decision to publish any data. All responsibility for the trial design, methods, data analysis, conclusions, and decisions regarding publication rest ultimately with Drs. Manary, Maleta and Brenna.*

### 1 Table of Contents

Enrollment	38

Allocation	39

Follow-Up	39

Analysis	39

### 2 Abbreviations and Definitions

| AE | Adverse Event |
| --- | --- |
| ALA | Alpha-linolenic acid |
| ARA | Arachidonic acid |
| DHA | Docosahexaenoic acid |
| DHA-HO-RUTF | High oleic acid RUTF with added DHA |
| EPA | Eicosapentaenoic acid |
| HAZ | Height-for-age z-score |
| HO-RUTF | High oleic acid RUTF |
| LA | Linoleic acid |
| MAM | Moderate acute malnutrition |
| MDAT | Malawi Development Assessment Tool |
| MUAC | Mid-upper arm circumference |
| PUFA | Polyunsaturated fatty acid |
| RUTF | Ready-to-use therapeutic food |
| SAM | Severe acute malnutrition |
| SRT | Saccadic reaction time |
| S-RUTF | Standard ready-to-use therapeutic food |
| WAZ | Weight-for-age z-score |
| WHZ | Weight-for-height z-score |

### 3 Trial Summary

#### 3.1 Type of study

Prospective, randomized controlled blinded clinical trial

#### 3.2 Problem to be studied

Cognition in children with severe acute malnutrition (SAM) treated with ready-to-use therapeutic food (RUTF).

#### 3.3 Interventions

Three different RUTF formulations will be compared: standard RUTF (S-RUTF), high oleic RUTF (HO-RUTF) made with high oleic peanuts and vegetable oils, HO-RUTF with added docosahexaenoic acid (DHA-HO-RUTF).

#### 3.4 Methodology

2700 children in Southern Malawi aged 6-59 months with uncomplicated SAM were randomized to receive S-RUTF, HO-RUTF, or DHA-HO-RUTF that provides 175 kcal/kg/day for up to 6 bi-weekly follow-up visits (or 12 weeks absent defaults). Anthropometry, demographics, symptom assessment, and past health information were recorded at enrollment. Anthropometry, compliance, and symptom assessments were repeated fortnightly until an outcome was reached. A plasma sample was obtained from a subset of children after 4 weeks of treatment. Recovery was defined based on the anthropometric criteria that were used to diagnose SAM. Default was defined by missing 3 consecutive visits. Death was recorded when reported by the caregiver. Children who did not recover after 6 follow-up visits were designated as “remained malnourished,” specified as to whether they had reached criteria for moderate acute malnutrition (MAM), or remained with SAM.

Neurocognition was assessed in a subset of participants using three tests:

- Modified Willatts problem solving assessments: intentionality was assessed in constructed scenes in which a child could retrieve an object with and without barriers between them and the object. This was assessed within 4 weeks of completion of nutritional therapy.
- Malawi Developmental Assessment Tool (MDAT): a standardized 4-part exercise, assessing gross motor, fine motor, language, and social skills. MDAT was assessed 6 months after completion of nutritional therapy.
- Saccadic eye-tracking assessment: an electronic, laptop-based tool that administers a visual paired comparison memory task and infant orienting with attention saccade task. This was assessed 6 months after completion of nutritional therapy.

#### 3.5 Primary outcomes

MDAT global z-scores and modified Willatts problem solving assessment intention scores

#### 3.6 Secondary outcomes

Saccadic eye-tracking assessments, recovery, fatty acid percentages in plasma, MDAT sub-domain z-scores, rates of gain in weight, mid-upper arm circumference (MUAC), and length, death, default, adverse reactions, adverse events

#### 3.7 Hypothesis

Children with SAM treated with HO-RUTF or DHA-HO-RUTF will have superior cognitive performance than those treated with S-RUTF.

### 4 Introduction

4.1 **General Background**

Undernutrition is a global problem implicated in nearly 50% of the approximately 5.6 million deaths of children under 5 years worldwide each year.^1^ Despite the success of humanitarian interventions, over 200 million children under 5 years continue to suffer from stunting and wasting as a consequence of undernutrition.^1,2^ Of the children who survive, lifelong morbidities including neurodevelopmental insults may persist despite the apparent physical recovery.^2,3^ Long-term follow-up of those who recover from undernutrition has revealed lower educational achievement, lower cognitive scores, and intellectual and learning disabilities throughout adolescence and into adulthood.^4-6^ Sustained cognitive and psychological deficits stemming from early life malnutrition contribute to lower human capital and adult productivity, which may further aggravate social and economic challenges for individuals in disadvantaged communities, contributing to a cycle of intergenerational poverty.^4,5,7^ These findings highlight the importance of understanding the neurocognitive changes associated with undernutrition and developing interventions that optimize brain development along with promoting physical growth.

Currently, community-based treatment for uncomplicated severe acute malnutrition (SAM) is the prescribed feeding of ready-to-use therapeutic food (RUTF). RUTF is a micronutrient-fortified, lipid-rich (45-60% of total energy), high-energy food (520-550 kcal/100 g) formulated to support rapid growth and promote weight gain in affected children. Formulations of RUTF commonly consist of four main ingredients: peanut butter, milk powder, vegetable oil, and sugar, with added vitamins and minerals. Although there are standards set by the World Health Organization, RUTF ingredients will vary depending on local availability and cost. Currently, the WHO standards stipulate that RUTF’s nutritional composition is to match the F-100 therapeutic milk, a “catch up growth” formula, used to treat SAM in hospitals.^3^ While macronutrient profiles are well described, specific fatty acid requirements of RUTF formulations have not been standardized. As a result, the use of locally available oils and conventional peanuts often result in RUTF formulations with fatty acid profiles that do not support optimal brain development.

4.2 **RUTF fatty acid composition suppresses omega-3 synthesis**

When RUTF was originally formulated, energy density was maximized by the inclusion of large amounts of lipid. This strategy also created a product with low water activity to avoid bacterial proliferation. Due to their wide availability and high-fat content (78% of total energy from fat), peanuts were chosen as the base of RUTF. At the time of its development, the only fatty acid composition specification given for RUTF was that the omega-6 polyunsaturated fatty acid (PUFA) should make up 3-10% of total energy and omega-3 PUFA make up 0.3-2.5% of total energy. The United Nations Children’s Funds (UNICEF) has additionally specified that the omega-6 to omega-3 PUFA ratio to be less than 10:1, as a dietary recommendation made for all healthy humans.

Most RUTF formulations are made with peanuts and additional readily available processed oils. Conventional peanuts, the base of RUTF, are high in the omega-6 PUFA linoleic acid (LA, 18:2n-6; 25-40% of total fatty acids) and contain negligible amounts of the inefficient precursor of active omega-3 PUFA, alpha-linolenic acid (ALA, 18:3n-3; 0.01-0.4% of total fatty acids).^8,9^ Additional oils from sources such as soy, sunflower, corn, cottonseed, and safflowers are common ingredients in processed foods, including RUTF. Most common oils are similarly high in LA and have low or negligible ALA, ^9-11^ whereas palm oil is predominantly comprised of saturated and monounsaturated fatty acids.^9^ As a source of ALA, canola oil (3-10% of total fatty acids) may be added to RUTF to improve the fatty acid profile, but even in canola oil the greater amount of LA compared to ALA further increases LA.^11^ Our calculations of oil composition of conventional RUTF formulations find omega-6 to omega-3 ratio to be always greater than 10:1, and as high as 53:1. Notably missing from the vegetable oils in RUTF are long-chain PUFA. As a result, children treated for SAM with RUTF rely completely on the endogenous synthesis and secretion to maintain circulating levels of conditionally essential long-chain PUFA; however, this very mechanism is inhibited by the high omega-6 LA content of standard RUTF.

The long-chain omega-6 PUFA arachidonic acid (ARA, 20:4n-6) and omega-3 PUFA docosahexaenoic acid (DHA, 22:6n-3) are synthesized by a series of elongation and desaturation reactions from shorter chain precursors LA and ALA, respectively. These 18-carbon precursor fatty acids compete for the same enzymes catalyzing the elongation and desaturation steps, and as a result the overabundance of omega-6 competitively inhibits the synthesis of the omega-3.^12^ As evidenced from experimental animal studies, excessive dietary intake of LA compared to ALA overwhelms the pathway and suppresses circulating levels of DHA.^13^ The same is true for conventional formulations of RUTF.^14-16^

In two separate blinded, randomized clinical trials measuring circulating DHA levels in children with SAM being treated with various RUTFs, those on conventional RUTF formulations experienced dramatic decreases in DHA from baseline measures.^14,16^ In a study of 61 children with SAM, 21 randomized to receive a standard RUTF with a LA: ALA ratio of 11.2:1, Jones et al. reported a significant 25% decrease in red blood cell DHA levels after 4 weeks of treatment with the standard RUTF.^16^ Similarly, Hsieh et al. reported a significant 25% decrease in plasma phospholipid DHA levels after 4 weeks of treatment with a control RUTF (LA: ALA ratio of 53:1).^14^ However, with a specially formulated, balanced dietary provision of LA and ALA (LA: ALA ratio = 1:1) using high-oleic peanuts, Hsieh et al. found plasma phospholipid DHA concentrations after 4 weeks of treatment remained unchanged from baseline. Thus children’s endogenous synthesis of DHA was sufficient to maintain an adequate amount of DHA when not challenged with overwhelming omega-6 LA.^14^ The substantial decreases in circulating DHA levels in children treated with standard RUTF formulations are of serious concern, as DHA supply to the brain is critical to support optimal neurocognitive development.

4.3 **Brain development and docosahexaenoic acid**

Brain development is vulnerable to environmental insults, such as dietary deficiency and more specifically insufficient omega-3 fatty acid intakes.^6,17^ Surprising to many, the brain is one of the fattiest organs in the human body with a lipid content exceeding 50% by dry weight.^18^ As is the case with virtually all tissues, the fatty acid environment of the brain is highly dynamic and supports much more than merely providing the structural components of cell and organelle membranes.

Docosahexaenoic acid comprises over 40% of total brain PUFA and has been shown to participate in the regulation of cell-survival and signal transduction, mediate neuroinflammation and neurogenesis and regulate blood-brain barrier permeability.^19,20-22^ During the third trimester of gestation, the brain reaches its maximal accretion rate for DHA, which continues to be high through approximately age two, a critical period of brain development.^23^ The brain continues to rapidly accrue DHA through childhood before plateauing during adolescence and young adulthood.^24^ A constant and adequate supply of circulating DHA is critical during this period as the brain cannot efficiently synthesize DHA from shorter chain fatty acid precursors. Although ALA rapidly enters the brain, ≤ 0.2% is converted to DHA.^25^ Furthermore, during this period the brain undergoes substantial structural and functional changes. Advanced neuroimaging studies have shown the morphological changes and rapid development of functional neurocognitive systems occurring during the first 5 years of life.^26^ The functional networks and connectome patterns established early in childhood influence cognitive function and behavioral skills throughout life.

Animal models of omega-3 deficiency demonstrate the significant negative functional and behavioral consequences of inadequate DHA during brain development. Diets used to establish omega-3 deficiency are often formulated with sunflower, safflower, corn, or peanuts oils and exhibit similar fatty acid profiles as some standard RUTF formulations.^11,14^ In the absence of dietary DHA, brain DHA levels may be supported by synthesis from ALA; at levels ≤0.8% of total fatty acid on a background of 23-24% LA, however, brain DHA levels decrease dramatically.^27^ At these levels of omega-3 deprivation, the brain reacts to preserve DHA through the widespread reduction in the activity of key DHA metabolizing enzymes (e.g. calcium-independent phospholipase A2 IV, cyclooxygenase 1), significantly reducing brain DHA turnover.^28^ Brain metabolic changes underlie the observed widespread functional neurological deficiencies associated with omega-3 deprivation, such as deficits in reflexes, maze learning, motor development, aggression, impulse control, and learning, among others.^11^

The importance of DHA in brain development is further supported through functional neuroimaging studies. Studies in children show that DHA supplementation has measurable effects on brain activity which may translate to improved learning and behavioral skills.^29,30^ Lacking from the literature, however, is any study on the neurocognitive support of DHA in a population suffering from malnutrition. Given that children with SAM have a high nutrient demand during recovery, which coincides with active brain development, they are likely to be at high risk of neurocognitive deficits resulting from inadequate omega-3 intake and conversely might experience the greatest benefits from supplementation.

4.4 **Dietary strategy to increase circulating DHA levels**

Increasing intake of ALA or other shorter chain omega-3 fatty acids such as eicosapentaenoic acid contributes to altering blood DHA levels.^31^ In healthy humans, targeted increases in circulating DHA levels can only be achieved through two approaches: providing preformed DHA in the diet, or by reducing dietary LA levels.^15,32^

In the last 20 years, plant breeding with a focus on oil composition has produced high oleic soy and peanut with lower omega-6 fatty acids.^33,34^ The use of high oleic variety ingredients over commodity alternatives results in reduced LA levels and a more balanced fatty acid profile of RUTF. In a study by Hsieh et al., the balance of omega-6 and omega-3 fatty acid profile of RUTF was achieved through the use of high oleic peanut and flaxseed oil.^14^ After 4 weeks of feeding high oleic-RUTF, plasma DHA levels, and weight-for-height z scores were higher than controls receiving a standard formulation. There were no significant differences in anthropometry, recovery, and growth rates between groups, demonstrating that high oleic-RUTF was well tolerated and did not compromise recovery.^14^

In addition to increasing omega-3 content of RUTF through added flaxseed oil, supplemental fish oil appears to be well tolerated by children with SAM.^16^ The addition to fish oil as a source of dietary DHA at 0.42% of total fatty acids on top of a flaxseed-based RUTF resulted in a 17% increase over baseline red blood cell DHA levels after 12 weeks and no significant differences were measured with respect to safety outcomes.^16^

Although previous studies have proven that alternative RUTF formulations with high oleic peanut and added DHA can successfully target circulating DHA levels, none have evaluated the associated impact on neurocognitive development. The current study aims to investigate the effect of a novel high oleic-RUTF and high oleic RUTF with added DHA on neurocognitive performance in children with uncomplicated SAM living in rural southern Malawi.

4.5 **Cognitive assessments**

The primary endpoints of this study are cognitive developmental outcomes which are challenging to assess in infants, especially in rural and low-resource settings. We will use three separate tests to capture different facets of development and cognition, aiming for a comprehensive assessment. One test, the Malawi Developmental Assessment Tool, was developed in Malawi as a culturally sensitive test of global development and was validated in over 1,500 children. A second, a modified Willatts problem solving assessment, has low technological demands, focuses on executive function and intentionality, and has been shown to respond to long-chain polyunsaturated fatty acid supplementation. The third, an automated saccade eye-tracking assessment, uses looking behavior as a window into attention and novelty preference, and is technologically advanced but has been successfully used in rural, low-resource settings. As detailed above, there is reason to suspect the fatty acid profile of S-RUTF is not optimal for promoting cognitive development. In this study, we will investigate the effects of novel HO-RUTF and DHA-HO-RUTF on neurocognitive performance in children following treatment of uncomplicated SAM living in rural areas in southern Malawi. Details of the tests are below.

4.5.1 **Modified Willatts problem solving assessment**

Problem solving, or “means-end behavior”, is an important component of cognitive development in infants. Means-end behavior specifically involves “the deliberate and planful execution of a sequence of steps to achieve a goal.”^35^ The key cognitive element drawn out by means-end testing is intentionality, wherein children must complete subgoals to allow the possibility of achieving their final goal. Planning and persistence are essential to success. Testing this domain in infants often involves the use of barriers placed between them and their ultimate goal, such that they must plan and intentionally act to achieve the goal. Examples of barriers used in prior research in this field include removing a cover or pulling a cloth to obtain an object resting on it.^36,37^

The problem solving assessments used in this study are modified from a protocol created by Willatts et al. ^35^ These tests aim to quantify problem solving such that test scores will reflect the infant’s level of cognitive sophistication. They do so by presenting a protocolized series of problems to the infants wherein different barriers are placed between the child and their goal of obtaining a toy. The purpose of the test is to assess intentionality in solving the problems, which involve pulling the toy closer by grabbing a cloth, uncovering the toy, or a combination of both of these.

This specific problem solving assessment tool was chosen for multiple reasons, including prior validation, low technological requirement, ease of training assessors, standardized coding procedures, and its use in research wherein the investigators assessed neurocognitive development in response to long-chain polyunsaturated fatty acid supplementation.^38^ In this study, 3-month-old infants were provided with one of two infant foods, one of which contained long-chain polyunsaturated fatty acids while the other did not. Follow-up intention testing at 9 months of age identified a trend toward improved intention scores and number of solutions.

4.5.2 **Malawi Development Assessment Tool**

Developmental assessment tools produced in Europe and the United States have been translated and adapted for use in other settings but are constrained by multiple factors, including culture-specific tasks and items, technological requirements, and limited validation. Investigation of several of these tools in a rural Malawian setting showed good reliability for some aspects of development, such as gross and fine motor skills, but poor reliability for social items.^39^ The Malawi Developmental Assessment Tool has been developed and studied over the past decade to address these limitations.

MDAT is composed of four domains that aim to summarize key aspects of development: language, social, gross motor, and fine motor. To perform the test, a trained assessor elicits a series of actions by the child and asks the caregiver questions about their child. These tasks and questions have pictorial representations to aid in understanding. Assessment items were generated via qualitative studies including focus groups of villagers and professionals in Malawi.^40^ Pilot studies were done to operationalize and test these items. Thereafter, a large-scale community study was performed, including over 1500 children, to test the reliability and validity of MDAT and to establish normal reference ranges.^41^ It showed excellent reliability across different observers and over time, as well as high sensitivity (97%) and specificity (82%) for neuro-disability.

MDAT has been tested in undernourished children in multiple studies. In a field study in Malawi, children with marasmus were tested and found to have substantially higher rates of failure, with lower scores in all domains except social development.^41^ Stunting and wasting were associated with significant reductions in approximately half of the MDAT tools, suggesting MDAT is sensitive to undernutrition. A study of children hospitalized with SAM in Malawi identified profound delays across all domains, including significantly worse language scores in those with kwashiorkor compared to marasmus.^42^ A study in Burkina Faso investigated the impact of different supplemental foods in the treatment of moderate acute malnutrition (MAM) using the MDAT and found improvements in multiple domains.^43^ A separate study in Burkina Faso investigated correlates with MDAT in MAM and found that higher anthropometrics and omega-3 PUFA levels correlated with higher MDAT scores.^44^ Taken together, MDAT has been used by a host of different research groups in various settings over the past decade and has been shown to be reliable, valid, and sensitive to undernutrition.

4.5.3 **Saccade eye-tracking assessment**

Infant looking behavior has been studied as a developmental measure for decades across many contexts. An infant’s ability to orient their gaze to novelty and hold their gaze on key sources of information offers insight into their attention and memory, is important for their ability to interact with their environment, and emerges during the first six months of life.^45-48^ Visual orienting speed in infancy has been associated with future executive function and performance IQ, and reduced orienting to faces has been associated with social-behavioral problems at 4 years of age.^49-51^ Prior research has suggested looking behavior may be more sensitive than global developmental measures.^52^ Our research group recently studied visual orienting in the context of moderate acute malnutrition in Sierra Leone and found a preliminary association between therapeutic feeding and reduction in saccade reaction time (manuscript in press).

The replacement of manual methods of eye-tracking assessment with automated, technologically advanced methods has benefited the field. These methods take advantage of the fact that the cornea of the human eye reflects infrared light, which can thus be used to track eye movement using a camera. In general, these automated tests use a display screen to show the child's attention-grabbing images in different locations across the screen. Software calibrates the location of the child’s eyes in relation to the monitor and links the display monitor to the eye tracker, thereby allowing the software to identify the location of the stimulus on the display and whether the infant’s gaze has fixed on that location. While initially these techniques were limited to high-resource settings, more recently they have been expanded for use in low or middle-income settings.^53,54^ A recent study in Malawi used a fully automated system for eye-tracking to assess the effect of supplemental feeding on development in 6-9-month-old infants. We have adopted this system with the help of its primary developer for the improved PUFA study.

### 5 Study objectives and endpoints

#### 5.1 Study objectives

5.1.1 **Broad**

To compare cognitive-developmental measures in children treated for SAM with 3 formulations of RUTF with different fatty acid compositions.

5.1.2 **Specific**

To compare problem solving assessment intention scores, Malawi Developmental Assessment Tool (MDAT) scores, saccade eye-tracking assessment scores, biochemical plasma fatty acid profiles, recovery, growth, infectious symptoms, mortality, and adverse events in 6-59 month-old children with SAM treated with S-RUTF, HO-RUTF, or DHA-HO-RUTF in a 7 visit / 12-week home-based therapeutic feeding program.

#### 5.2 Endpoints

5.2.1 Primary
- Malawi Developmental Assessment Tool global z-score
- Modified Willatts problem solving assessment intention scores
  5.2.2 Secondary
- MDAT sub-domain z-scores: gross motor, fine motor, language, and social
- Nutritional recovery, defined as resolution of nutritional edema for at least 2 weeks, and WHZ > -2 or MUAC ≥ 12.5 cm depending on criteria used to diagnose SAM at enrollment
- Serum levels of fatty acids, including oleic acid, eicosapentaenoic acid (EPA), docosahexaenoic acid (DHA), alpha-linolenic acid (ALA), linoleic acid (LA), and arachidonic acid (AA)
- Saccade eye-tracking assessment scores
- Rates of gain in weight, mid-upper arm circumference (MUAC), and length
- Mortality
- Defaults
- The average number of days of fever, diarrhea, poor intake during treatment

### 6 Study methods

#### 6.1 General study design and plan

This was a prospective, individually randomized controlled clinical trial. All endpoints were compared using a superiority framework. The control type was standard of care, S-RUTF with amoxicillin for 7 days. The intervention arms were HO-RUTF with 7 days of amoxicillin and DHA-HO-RUTF with 7 days of amoxicillin. The participants, caregivers, investigators, and outcomes assessors were blinded. Participants were randomized individually before treatment initiation 1:1:1 to the three study arms. Screening, inclusion, and active treatment started in October 2017 and ended December 2020. Each participant was followed until a programmatic outcome (recovery, death, default, transfer to inpatient facility) or 6 bi-weekly follow-up visits if they remained acutely malnourished despite treatment, whichever occurred first. If the participant remained malnourished after 6 follow-up visits, they were referred to their local health center for evaluation.

#### 6.2 Study setting

The study was initially planned to take place at 20 rural health clinics: Chamba, Chikonde, Chikweo, Chipolonga, Makhwira, Matiya, Mauwa, Mayaka, Mbiza, Milonde, Mitondo, Mlomba, Muloza, Namasalima, Naphimba, Ndakwera, Nkalo, Nkhate, Nsanama, and Zombasalima. These sites are located in 5 districts: Chikwawa, Chiradzulu, Machinga, Mulanje, and Zomba. Most people who live in these areas farm for subsistence, with maize as the staple crop and main source of nutrition. Water is collected from boreholes or rivers several times per day. Homes are built of mud bricks, often single-room, and roofed with thatch or occasionally corrugated metal. Electricity and plumbing are unavailable, and defecation occurs outdoors or in pit latrines. With rare exceptions, women perform household labor and child-rearing, and most are illiterate.

Sites were accessible by vehicle and contained a building with a roof. Sites were chosen in consultation with district health officials and village elders and leaders, including chiefs and health surveillance assistants. In general, these sites had limited access to other treatment programs and were known from prior research experience to contain populations at risk for severe acute malnutrition.

*Protocol addendum*

In 2018, 5 enrollment sites were added: Kasinthula, Makokola, Milepa, Nasfan, and Thumbwe. In 2018, study enrollment stopped at Nasfan. In 2019, 2 enrollment sites were added, Dolo and Nkumaniza. In 2019, enrollment was discontinued at Chamba, Kasinthula, Mauwa, Nkalo, and Thumbwe. In 2020, Bondo was added as an enrollment site. In total, study participants were enrolled from 28 clinic sites: Bondo, Chamba, Chikonde, Chikweo, Chipolonga, Dolo, Kasinthula, Makhwira, Makokola, Matiya, Mauwa, Mayaka, Mbiza, Milepa, Milonde, Mitondo, Mlomba, Muloza, Namasalima, Naphimba, Nasfan, Ndakwera, Nkalo, Nkhate, Nkumaniza, Nsanama, Thumbwe, and Zombasalima (map below).

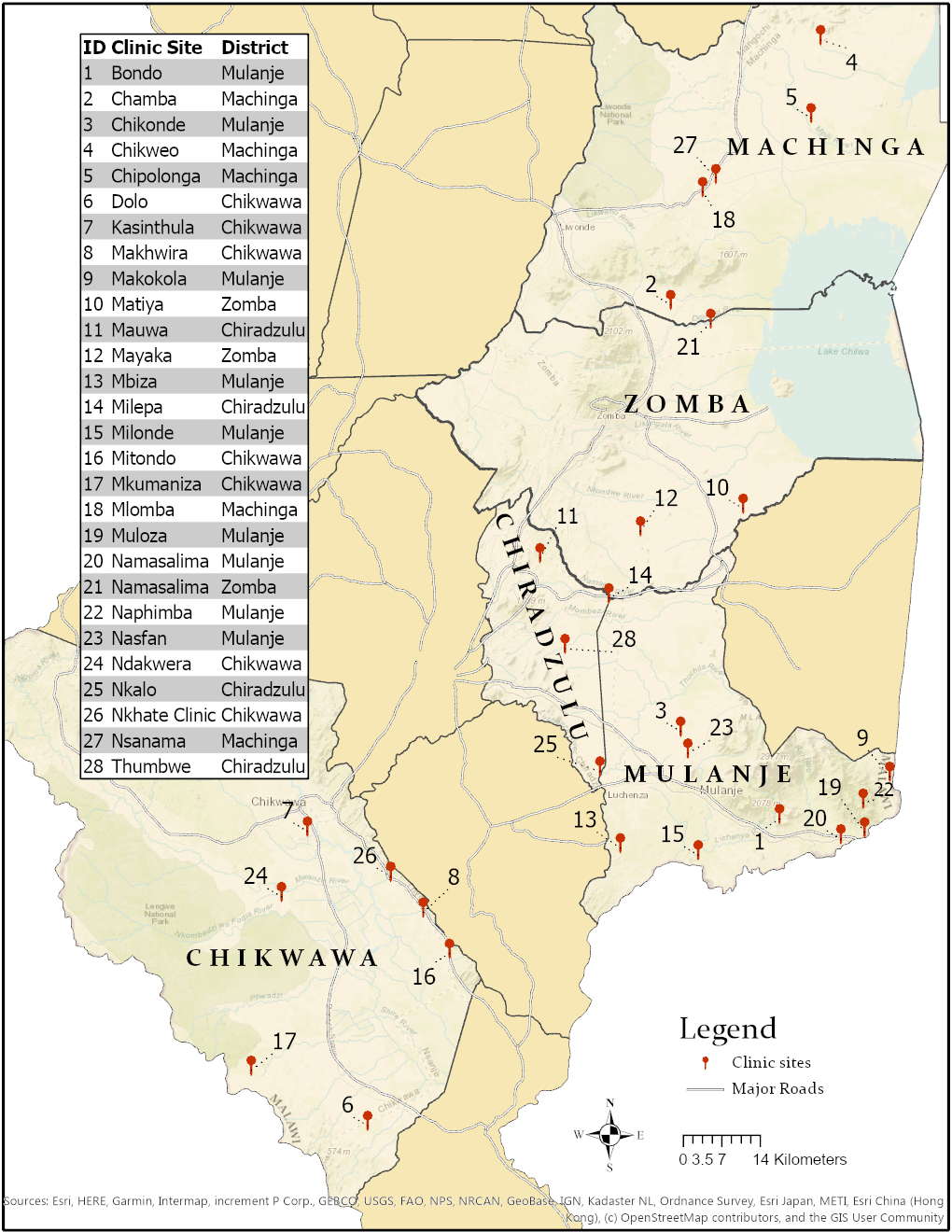

#### 6.3 Study population

Malawian children aged 6-59 months with uncomplicated SAM (defined by the presence of bipedal pitting edema, and/or MUAC < 11.5 cm, and/or WHZ < -3, no active fever, rapid breathing, or altered mental status, adequate appetite) who live in the vicinity of the study sites in rural Southern Malawi.

#### 6.4  Inclusion and exclusion criteria

6.4.1 **Inclusion criteria**
- Age 6-59 months, rounded to the nearest month
- MUAC <11.5, and/or weight-for-height z-score < -3, and/or bilateral pitting edema on the dorsum of feet
- An acceptable appetite, as defined by the ability to consume 30 grams of RUTF within 20 minutes
- Presentation to one of the included feeding clinics during the recruitment period
- Permanent residents in the vicinity of one of the feeding clinics
  6.4.2 **Exclusion criteria**
- Participation in a therapeutic or supplementary feeding program in the past 1 months
- Participation in any other ongoing study or supplementary feeding program
- Children with a chronic debilitating medical condition, including cerebral palsy, static encephalopathy, or congenital heart disease.
- Children with peanut allergy
- Children with vision or hearing problems

#### 6.5 Interventions

The interventions were specially prepared RUTFs, made by Project Peanut Butter in Blantyre, Malawi.^55^ In this study, three RUTF formulations were compared: DHA-HO-RUTF, HO-RUTF, and S-RUTF. The formulations of these foods are shown in Table 1, while Tables 2 and 3 detail the macronutrient and micronutrient contents, respectively.

6.5.1 **PUFA optimized RUTF development**

The goal was to produce two alternative RUTF formulations to achieve improved polyunsaturated fatty acid (PUFA) profiles, one with preformed DHA and one without. A linear programming (LP) tool was used to consider local commodities while maintaining the desired nutrient profile, with a focus on the omega-3 alpha-linolenic acid (ALA) and omega-6 linoleic acid (LA) nutrient constraints.^56^ The LP tool is designed to aid in the development of alternative RUTF formulations by pulling upon a database of candidate ingredients, with associated nutrient contents. It then calculates potential formulas which meet RUTF nutrient requirements of the joint United Nations agency statement in addition to the optimized omega-3 and omega-6 constraints.^3,57^ The UN agency specifications for FA in RUTF are omega-6 FA should be 3-10% of total energy and omega-3 FA should be 0.3-2.5% of total energy.

The first objective of the formulation process was to generate RUTF with improved omega-6 and omega-3 content. The candidate ingredient list included milk powders, vegetable oils including high-oleic oils, high oleic peanuts, and sugar. The paper formulation process started with an adjustment in the constraint for omega-3 FA to target a minimum of 1.5% of total energy. To further optimize the formulation, this was followed by dropping the maximum percent of omega-6 from energy to meet a favorable threshold for an ideal fatty acid balance and acceptable palatability. Potential alternative PUFA optimized RUTF formulations were prepared at the bench top level in the food lab at Washington University in St. Louis. Bench top production of improved PUFA formulas was evaluated primarily on organoleptic characteristics and production feasibility.

After organoleptic and production feasibility optimization, the optimal high-oleic RUTF (HO-RUTF) formulation was selected. Then food-grade DHA and EPA found in MEG-3 DHA K powder were added to the ingredient list with a target ingredient constraint of 1.5%. The MEG-3 is derived from fish oil but is encapsulated to reduce the organoleptic properties. Encapsulation is achieved with the seaweed-derived hydrocolloid carrageenan in combination with starch. The encapsulation utilizes a double-wall barrier to preserve the omega-3 EPA and DHA while preventing tastes or smells to permeate the food matrix. The encapsulation also prevents oxidation of the PUFA, rendering a shelf life similar to S-RUTF. Additional refinement of both formulations occurred, ensuring they were as similar as possible except for the addition of the DHA powder.

6.5.2 **High oleic RUTF (HO-RUTF)**

HO-RUTF was formulated according to sample ingredients and with nominal LC-PUFA levels shown in **Tables 1 and 2.** The formula was chosen from about 20 sample formulas developed with our in-house food component/nutrient database and optimization system as described above.^58^ The primary ingredient differences in lipid sources compared to the standard RUTF are the replacement of regular peanuts with high oleic (HO) peanuts, the addition of a small amount of perilla oil, and the removal of canola oil. Because the objective of the formulation process was to lower the LA as much as possible, the formulations were pulling less overall lipid from vegetable oils and more from the HO peanut, which also added protein. To ensure the formulation remained within the specifications for other macronutrients such as protein, a small quantity of the dried nonfat milk was replaced with sweet whey powder, which is lower in protein and higher in lactose. The amount of emulsifier was decreased to establish optimal suitability for large-scale production. The final formulation featured a simple set of readily available ingredients and produced a palatable product.^59^ It achieved the lowest LA practically achievable with common ingredients, was well within the UN agency specifications for both LA and ALA, and therefore was a completely balanced food raising no concerns over nutritional adequacy.

6.5.3 **DHA-High Oleic-RUTF (DHA-HO-RUTF)**

DHA content was formulated based on the natural intake of DHA from breast milk. The mean 24 h output of DHA in breast milk was 49 mg over the first year of life in five carefully followed mothers.^60^ Other literature estimated global mean breast milk DHA output to be 110 mg/d.^61^ Breast milk eicosapentaenoic acid (EPA) is approximately 25% of the DHA levels across nine countries.^62^ Using these benchmark natural intake levels, we formulated DHA-HO-RUTF to deliver nominal levels of 100 mg DHA and 25 mg EPA per day. Food grade DHA and EPA are readily available from many suppliers, and with care, in formulation and storage, the RUTF resisted oxidation.

6.5.4 **Standard RUTF (S-RUTF)**

Standard RUTF (S-RUTF) is composed of 4 ingredients in roughly equal amounts; milk powder, peanuts, vegetable oils, and sugar. S-RUTF is made from conventional, high omega-6 PUFA peanuts, as well as palm and canola oils. It meets the UN specifications for omega-3 and omega-6 PUFA content; omega-3 PUFA constituted 0.3-2.5% of the energy in RUTF and omega-6 PUFA constituted 3-10% of the energy in RUTF.^63^ A multiple micronutrient powder was added to this mixture to provide a full complement of vitamins and minerals. These powdered ingredients were mixed into a lipid-rich paste to create a soft, semi-solid consistency that can safely and easily be consumed by infants.^55^ RUTF has very low water activity making it difficult to contaminate because it does not support the growth of bacteria.^64^ This low moisture, energy-dense, homogenous lipid matrix, therefore, can safely be used at home even if hygiene conditions are not optimal. It is also shelf-stable, requiring no refrigeration.

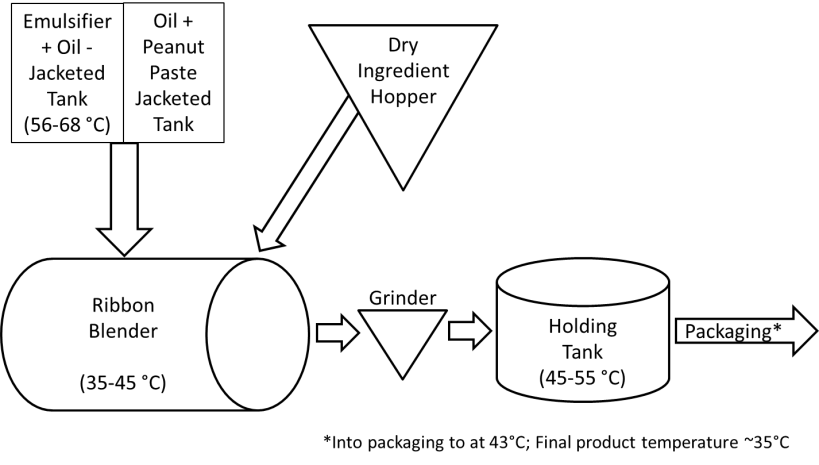
To achieve a homogeneous mixture, a specific mixing procedure was followed. In large-scale production, the process illustrated in the figure below summarizes the required steps and processing limits.

Oil and powdered hydrogenated vegetable oil emulsifier, specifically Caravan Trancendim 180, are melted to about 64°C. The remaining oil and peanut paste are released into the ribbon blender and mixed for 6 minutes. The oil/emulsifier mixture is added and mixed for 2 minutes. The dry ingredients are then dispensed and mixed with the paste ingredients for 10 min. Heating the oil and the mixing process warms the mixture, thus the temperature in the ribbon blender can reach 45°C. A grinding step follows, and depending on the equipment set up, the RUTF may be passed through the grinder three times to homogenize and reduce particle size. The RUTF is held in a tank until it is pumped to packaging with slow mixing using a rotating paddle. The temperature in the holding tank may reach 55°C, and the packing temperature is around 43°C.

The World Health Organization, World Food Programme, United Nations System Standing Committee, and the United Nations Children’s Fund in 2007 jointly published specifications for RUTF to ensure a worldwide supply of effective, safe RUTF products.^3^ Nutritional and microbiological requirements were also subsequently established.^63^ RUTF must also comply with the Recommended International Code of Hygiene Practice for Foods for Infants and Children of the Codex Alimentarius, and all mineral salts and vitamins must be listed on the Advisory List of Mineral Salts and Vitamin Compounds for Use in Foods for Infants and Children, also from Codex.^63^ All foods were locally manufactured by Project Peanut Butter in Blantyre, Malawi, which is fully certified as a RUTF producer by the United Nations Children’s Fund (UNICEF). Batches were certified by a battery of 12 microbiological tests for which the sampling methodology, test method, and test laboratory are specified by UNICEF, primarily testing for *Salmonella* and *Enterobacter spp.*^65^

HO-RUTF underwent acceptability testing before the aforementioned pilot study.^14^ A double-blind, randomized assessment of 148 children ages 6-59 months was done wherein half received S-RUTF and half HO-RUTF. All children were given 30g of the assigned food and timed during consumption. If after 40 minutes not all food was consumed, the remaining amount was weighed, the time recorded, and caregivers completed a survey regarding the child’s appetite and likeability of the food. In a second stage, 57 of these children (28 S-RUTF, 29 HO-RUTF) were provided with 270g of the assigned food, to be consumed in 90g increments over 3 days. On the 3rd day, the caregivers completed an additional likeability assessment. The results indicated high likeability, with survey results of 5/5 (highest on the scale) for 64/74 of those who received S-RUTF and 59/74 of those who received HO-RUTF. All 57 caregivers indicated 5/5 scores after the second stage of testing.

The food scientist working for the study team made a variety of prototype recipes for HO-RUTF and DHA-HO-RUTF from which the final recipes were chosen. These underwent informal acceptability testing at the time of bench top preparation, and all were deemed equivalent to S-RUTF.

**Table 1. Ingredient content of study RUTFs**

| **Ingredient*** | **High Oleic RUTF** | **DHA High Oleic RUTF** | **Standard RUTF** |
| --- | --- | --- | --- |
| Milk powder, non-fat, dry | 22.0 | 22.0 | 27.3 |
| Milk, sweet whey powder | 6.1 | 6.1 | 0.0 |
| Palm oil | 14.0 | 14.0 | 16.0 |
| Canola oil | 0.0 | 0.0 | 5.1 |
| Sugar | 26.0 | 24.5 | 23.0 |
| High oleic peanuts | 25.0 | 25.0 | 0.0 |
| Standard, high omega-6 peanuts | 0.0 | 0.0 | 23.7 |
| Perilla oil | 3.0 | 3.0 | 0.0 |
| Soy lecithin & a hydrogenated vegetable oil emulsifier** | 1.0 | 1.0 | 2.0 |
| Micronutrient powder | 2.9 | 2.9 | 2.9 |
| DHA powder | 0.0 | 1.5 | 0.0 |

*Content expressed in g/100g RUTF

**This was a proprietary commercial product, Trancendim 180 (Caravan Foods).

**Table 2. Macronutrient content of study RUTFs**

| **Nutrient*** | **High Oleic RUTF** | **DHA High Oleic RUTF** | **Standard RUTF** |
| --- | --- | --- | --- |
| Protein, total | 15.2 | 15.2 | 15.7 |
| Protein, dairy | 8.9 | 8.9 | 9.8 |
| Fat total | 29.5 | 30.2 | 32.7 |
| Omega-6 fats (LA) | 2.7 | 3.1 | 6.3 |
| Omega-3 fats | 1.7 | 2.2 | 0.5 |
| DHA, mg | 0.0 | 198.0 | 0.0 |
| EPA, mg | 0.0 | 40.5 | 0.0 |
| Energy, total | 531.0 | 532.0 | 541.0 |

*Content expressed in g/100g RUTF, except for DHA and EPA which are mg

**Table 3. The micronutrient content of RUTF**

| **Nutrient** | **RUTF** |
| --- | --- |
| Vitamin A, mg/100g | 1.07 |
| Vitamin C, mg/100g | 55.90 |
| Vitamin D, µg/100g | 28.00 |
| Vitamin E, mg/100g | 44.50 |
| Vitamin B6, mg/100g | 2.00 |
| Vitamin B12, µg/100g | 32.70 |
| Riboflavin, mg/100g | 3.88 |
| Niacin, mg/100g | 23.90 |
| Thiamine, mg/100g | 1.46 |
| Iron, mg/100g | 14.20 |
| Zinc, mg/100g | 14.80 |
| Calcium, mg/100g | 658.00 |
| Phosphorus. mg/100g | 651.00 |
| Magnesium, mg/100g | 172.00 |

#### 6.6 Randomization

Participants were randomized individually in a 1:1:1 ratio to the three intervention arms. Computer-generated block randomization lists were created in blocks of 54, designed using 6 colors, 2 of which were randomly assigned to each study group using a random number generator. One color was inserted each into a small, opaque envelope which was then sealed. These opaque envelopes were indistinguishable externally. Larger manila envelopes were then each filled with a block of 54 smaller envelopes, such that 9 envelopes were contained for each of the 6 colors.

At the time of randomization, the caregiver of each participant was asked to randomly select one small opaque envelope from the larger manila envelope, thereby assigning the study group. Once the group was assigned, this color was written on the top of the participant’s study card.

#### 6.7 Blinding

Study foods were dispensed in sachets containing 92g of study food and were identical in external appearance aside from a colored sticker added to indicate the intervention group. Boxes contained 1 of 6 colors corresponding to the randomization group and were labeled with that color. As previously described, a pilot study was completed before the initiation of the present study to assess acceptability, appearance, and taste of each intervention, with attempts made to mask any changes related to HO peanuts and DHA to preserve participant and caregiver blinding.^14^

Team members responsible for test feeding, nutrition counseling, questionnaire completion, anthropometric measurements, determination of programmatic outcomes, and neurocognitive assessments were unaware of and did not have access to the allocation code. Team members responsible for dispensing the intervention therapeutic foods did so by matching the color on each participant’s study card with the box color label. These study members also did not have access to the allocation code.

The senior study investigators (Drs. Manary, Maleta, Stephenson) will not have access to the allocation code until the study is finished and data analysis has been completed. The investigator responsible for statistical analysis (Dr. Stephenson) will take no part in the study otherwise, including no clinic visits, measurements, or outcome determinations. Dr. Stephenson will be provided with a code that is linked to the study assignment in a separate file to which he will not have access. The investigators responsible for calculating the problem solving assessment scores, MDAT scores, and saccade eye-tracking assessment scores will take no part in the study otherwise.

#### 6.8 Consent

Nurses were responsible for describing the purpose and nature of the study to caregivers. They would sit with the caregivers away from the center of the clinic and speak confidentially. They would describe SAM, RUTF, anthropometric measurements, blood draws, and cognitive tests. They would also explain how the study foods were different and prior testing that demonstrated acceptability and safety of all study foods. They would describe the role of antibiotics in the treatment of SAM. They would assure the caregivers that participation was voluntary and that the caregivers could withdraw their children at any time for any reason if they so choose, without repercussion and with the assurance of continued SAM treatment if indicated. They would inform the caregivers that additional children in their household could be treated if malnourished, but not in the research study. They described how their child’s data will be kept confidential and protected, with de-identification when first possible, and assured that any private information they provide would be secured and used only for the study in question. They described the tokens of appreciation that would be provided to caregivers at study milestones and that these tokens were not contingent on continued participation after receipt. They described the potential impact of the research and the role caregivers and their children will play in generating new knowledge. They described the process of default tracing if the caregiver were not to arrive at the clinic, as well as the hope that caregivers would either return to the clinic or send a message if their child were hospitalized or had passed away. Finally, they answered any questions and then proceeded to explain the mechanics of consent, whether by signature or inked thumbprint (consent form attached at end of protocol).

#### 6.9 Study Team

Team members included nurses, drivers, research assistants, and neurocognitive assessors. Nurses were responsible for assessment for inclusion and exclusion criteria other than anthropometric measurements and edema assessment, explanation of study procedures including blinding process, appetite test, consenting caretakers, nutritional counseling of caregivers at enrollment, and each follow-up with special attention to those who were not recovering, identification of children who required transfer to inpatient care, blood draws, explanation of follow-up schedule. Drivers were responsible for the distribution of intervention foods, distribution of pre-determined tokens of appreciation to caregivers during study procedures, and monitoring compliance to study foods. Research assistants were responsible for the evaluation of age criteria, measurement of MUAC and length, identification of SAM by WHZ and/or MUAC, and pedal edema assessments. Neurocognitive assessors were responsible for problem solving assessment, MDAT, and saccade eye-tracking assessments.

#### 6.10  Clinic Flow

6.10.1 **New child and caregiver arrive at study clinic**
- The child is undressed by the caregiver and weight is measured to the nearest 5 grams using a seated digital scale (Seca 334)
- MUAC measured to nearest 1mm using standard insertion tape
- Length measured in triplicate to nearest 2mm using rigid length board (Seca 417)
- WHZ score identified using a WHZ field chart (attached)
- Pedal edema assessment
- If the child meets the criteria for SAM, the age eligibility is checked using their health passport
  - If the WHZ criterion is met, target weight is calculated using WHZ field chart and added to the child’s health passport
  - If they meet the criteria for MAM, they are enrolled in a standard MAM feeding program
  - If they are not malnourished, caregivers are told the child’s well-nourished status
- If the child meets age criteria, “PUFA?” is written on their health passport and they are directed to the study nurse station
- A research nurse assesses eligibility based on additional inclusion and exclusion criteria
  - If the child appears to have severe acute illness including but not limited to rapid breathing, high fever, or altered mental status, they will be transported to the closest inpatient nutritional rehabilitation unit
- The nurse performs test feeding of 30g of RUTF
  - Consumption of the test dose within 20 minutes is required for study admission.
  - If the child is unable to consume the 30g, they will be transported to the closest inpatient nutritional rehabilitation unit
- If eligible, the nurse explains study procedures and asks the caregiver if they will consent
  - If not eligible but deemed to be safe for community-based management of SAM, they will be given 2 weeks of RUTF and asked to return in 2 weeks for non-research SAM follow-up
- The caregiver can either sign their name or place an inked thumb print on the consent sheet if they have chosen to participate
  - If they have chosen not to participate, the child will be provided with a 2-week provision of RUTF and asked to return in 2 weeks for non-research SAM follow-up
- A nurse explains the randomization procedure and holds out a larger manila envelope that contains smaller, individually packaged opaque envelopes that each contain one of six study colors, two of which correspond to each intervention
- Caregiver selects one card at random and opens it to reveal their allocation
- Intervention allocation is added to a study card, which is then filled out with demographic, anthropometric, health, and illness symptom information
- The date of the follow-up visit is filled into the card and explained to the caregiver
- A nurse explains the role of amoxicillin and teaches the caregiver how to administer it for seven days
- The caregiver is directed to study driver station, where the driver determines the amount of intervention study food to provide based on a chart with pre-calculated sachets to provide 180 kcal/kg/day
  6.10.2 **Returning caregiver and child arrive at the clinic with a small identification study card**
- A full study card is identified in the site binder
- Weight, MUAC, and length measurements are performed
- Edema assessment done
- Research assistant assesses for the fulfillment of recovery criteria
  - If enrolled based on edema alone, resolution of edema for at least 2 visits
  - If enrolled based on WHZ, target weight achievement
  - If enrolled by MUAC, MUAC ≥ 12.5cm
  - If enrolled by both WHZ and MUAC, whichever criterion is met first
  - If enrolled by edema and anthropometric criteria, resolution of edema for at least 2 visits and achievement of threshold for recovery for anthropometry
- If the child has recovered:
  - They are directed to the nursing station for symptom assessment
  - If they are not in the subset to undergo neurocognitive testing, they are directed to the driver’s station to receive a token of appreciation for their participation
- If the child has not reached criteria for recovery and they have not yet had 7 visits:
  - They are directed to the nursing station for counseling
  - If the child continues to appear safe to continue community-based treatment, symptom assessment performed and a follow-up date given
  - Caregiver directed to driver’s station
  - Intervention food distributed
- If the child has not reached criteria for recovery and they have had 7 visits:
  - They are given the outcome of “remained malnourished”
  - They are counseled about the next steps and offered a ride to the nearest inpatient nutritional rehabilitation unit if the mother consents
- If the child has reached an outcome as above and is part of the subset to undergo MDAT and eye-tracking neurocognitive testing
  - The caregiver is given a date to return for MDAT and saccade eye-tracking assessments
- If the child has reached an outcomes as above and is part of the subset to undergo PSA neurocognitive testing
  - They are then directed to the neurocognitive assessor station for problem solving assessment or, if unable to take part that day, are given a date to return within 4 weeks of SAM treatment outcome date
  - Following neurocognitive assessments, they are directed to the driver station for the provision of tokens of appreciation
    6.10.3 **Returning caregiver arrives to inform staff their child has passed away or has been hospitalized**
- The nurse speaks with the caregiver about the time and cause, if known, of the child’s death or reason for hospitalization

#### 6.11  Defaulter Tracing

Children enrolled in the research study who were not brought back for their scheduled follow-up visit were sought at home by the Community Health Workers via a “send-the-message” card on which the Research Assistant would transcribe the child’s caretaker’s name and directions to their domicile. The Community Health Worker would be instructed to locate the child and ascertain the child’s clinical status by MUAC, presence of edema, and general clinical assessment. Acutely ill children would be referred to the health center. The child’s caretaker would be encouraged to return to the clinic the next time that the research team would be present there.

If the child missed a second follow-up visit, a second “send-the-message” card would be sent. If the child missed a third follow-up visit, the entire study team would attempt to locate the child after clinic with the aid of the project 4x4 vehicle. If the child was still unable to be located, they would be classified as a defaulter for the study.

Caregivers were assured that they could withdraw their child from the study at any point and for any reason if they so choose. Study staff discussed the decision with the caregiver if the caregiver had concerns. The mother was assured her child would continue to receive treatment for SAM if indicated regardless of their decision to withdraw.

Community Health Workers received a small monetary gift and/or cell phone units for any successful tracing of children or information obtained via “send-the-message” cards.

#### 6.12  Study Assessments

Below is a table detailing the timing of study assessments and procedures if an individual were to remain in treatment for the full 7 visits. If alternatively, a participant reached a programmatic outcome before visit 7, that outcome week would be the last week of assessment and they would not receive therapeutic food at that visit.

**Table 4. Study assessments**

| **Visit** | **1** | **2** | **3** | **4** | **5** | **6** | **7** |
| --- | --- | --- | --- | --- | --- | --- | --- |
| **Week*** | **0** | **2** | **4** | **6** | **8** | **10** | **12** |
| Test feeding** | X |  |  |  |  |  |  |
| Weight | X | X | X | X | X | X | X |
| Target weight calculation | X |  |  |  |  |  |  |
| Length | X | X | X | X | X | X | X |
| MUAC | X | X | X | X | X | X | X |
| Edema assessment | X | X | X | X | X | X | X |
| Symptom questionnaire | X | X | X | X | X | X | X |
| Socioeconomic questionnaire | X |  |  |  |  |  |  |
| Appetite assessment | X | X | X | X | X | X | X |
| Gross motor milestone assessment | X | X | X | X | X |  |  |
| Therapeutic food provision | X | X | X | X | X | X |  |
| Amoxicillin 7-day prescription | X |  |  |  |  |  |  |
| Blood test |  |  | X |  |  |  |  |

*Approximate, as default visits can extend the number of weeks in the study

**Can be done any week at the discretion of study nurses if there are concerns about the child’s ability to continue community-based treatment

Below is a table of cognitive-developmental assessment timing. These assessments were performed on a subset of study participants as determined by sample size requirements.

**Table 5. Neurocognitive assessment timing**

| Visit | Study outcome* | 6 months post-outcome** |
| --- | --- | --- |
| Problem solving assessment | X |  |
| Malawi Developmental Assessment Tool |  | X |
| Saccade eye-tracking assessment |  | X |

*Could fall within 4 weeks of outcome

**Could fall between 5-7 months post SAM program outcome

#### 6.13 Subgroup enrollment

6.13.1 **Neurocognitive testing**

Subsets of the study population underwent modified Willatts problem solving assessment testing, MDAT, and saccade eye-tracking assessments, as the required samples sizes for each were smaller than that of the entire study. For the problem solving assessment, beginning in February 2018, all participants under two years of age were assessed with a PSA within four weeks of their clinical outcome. Beginning in March 2018, all participants under 30 mo of age were asked to return to clinic five to seven months later for testing with the Malawi Developmental Assessment Tool and eye-tracking testing. This method allowed for adequate power of the neurocognitive outcomes, while not unduly subjecting children to the testing procedures.

6.13.2 **Plasma sampling**

A subset of the study population underwent a blood draw for plasma fatty acid testing. This subset was determined by participation during two designated periods of the study, from specified dates in January 2018 to October 2018 and from December 2019 to August 2020. During these intervals, all caretakers were asked at all study sites for permission to draw a blood sample. It was expected that many caretakers would opt out of the blood draw. The unusual choice for this subset was guided by a local fear in Malawi at the time that health workers who were drawing blood were using the blood to spiritually curse the child. This fear was so strong that in a context outside of our study, two health workers were murdered for blood collection.

#### 6.14 Intervention adherence and side effect assessments

6.14.1 **Adherence**

When the child was brought back for their first follow-up visit after enrollment in the study, the nurse would assess adherence to the intervention from the caretakers. All assessments were conducted in a non-judgmental way and it was made clear to the caretakers that we were only asking about adherence for the sake of data collection, and that the child’s continued care and feeding and the caretaker’s prizes for participation in the study would not be affected by the answers to the adherence questions. Assessment of adherence was done using 2 criteria: 1) verbal report from the caretaker; 2) examination of empty RUTF sachets. Based on these criteria, the nurse would document the number of days of intervention the child received on the child’s study data card.

6.14.2 **Side Effects**

The nurses also asked the caretakers about any potential associated side effects, including (but not limited to) abdominal pain, emesis, rash, and diarrhea. Specific questions about the timing of these side effects relative to entry into the study and the duration and severity would be asked. If there was any concern that a child may have had a side effect, an adverse drug reaction would be reported and discussed with the research team.

### 7 Ethical considerations

In accordance with the Declaration of Helsinki, we aimed to complete the study using the highest ethical standards for clinical trials research.^66^ Ethical approval was obtained from the University of Malawi College of Medicine Research and Ethics Committee and the Human Research Protection Office at Washington University in St. Louis. Approval was obtained from the University of Texas at Austin and Cornell University. Letters of support were obtained from the district health officers in each district where the study would be conducted. Local village chiefs surrounding the study sites gave their consent and support for the studies. Each child’s caregiver was briefed on the study procedures and interventions. Verbal and written informed consent was obtained from all caregivers. Caregivers who were unable to sign or write their names were asked to document consent using their thumbprints.

On the day of enrollment, caretakers were informed about the process of randomization, the duration and requirements of participation in the study, and the benefits of participation in the study through an interactive oral explanation. Only caretakers who expressed continued interest in the study after being fully informed were enrolled; otherwise, they were enrolled in the standard therapeutic feeding programs administered by Project Peanut Butter.

Children who were deemed ineligible to participate in the research study were treated in a standard SAM therapeutic feeding program unless they were too ill for community management, in which case they were transported to the closest inpatient nutritional rehabilitation unit.

Discussions with district health officers and health service assistants at each site occurred before the initiation of the study to address any ethical concerns they had. Individual written or thumb print consent was obtained at the time of enrollment.

### 8 Sample Size

The primary outcomes in this study are MDAT and problem solving assessment results, while the key secondary outcome is recovery. To determine the sample size needed for this study, we first assessed what sizes would be needed for each primary outcome as well as the key secondary outcome.

MDAT z-scores were expected to have a standard deviation of approximately 1.1 in this population, based on prior research.^67-69^ An effect size of 0.25 SD would represent a clinically meaningful result and using this difference with 25% attrition, parameters of 80% power, and two-sided 95% significance for superiority testing, 400 participants were required per group.

The problem solving assessment is a modified version of the Willatts test.^36^ In research testing the effect of DHA on infant cognition, Willatts et al. obtained a standardized effect size of 0.51 ((7.7 – 6) / 3.35). We did not anticipate an effect size this large in our trial for several reasons, including study environment (less well-controlled setting), participants will have either just recovered from or will continue to have malnutrition, the supplementation period is shorter, and the age distribution will be wider, likely increasing variance. Additionally, we could not predict with confidence the distribution of the PSA intention scores in our population because of the wider age range and presence of, or recent treatment for, SAM. Thus, this sample size calculation was uncertain. We estimated 20% will be unable to complete testing despite repeated attempts for a variety of reasons (including acute effects of SAM). Using parametric assumptions, and assuming an effect size one-half as large as that found in prior research and using parameters of 80% power and two-sided 95% significance for superiority testing, 300 participants would be required per group.

Based on prior experience, recovery rates from SAM in this setting are expected to be approximately 89%. If present, we aimed to detect superiority at 4%. Using parameters of 80% power and two-sided 95% significance for superiority testing, 800 participants per group were required to detect superiority at 4%. To account for defaults and other data collection issues, 900 participants per group were sought for recruitment. Because this secondary outcome was deemed essential, and because it required the largest sample size, this determined the total study sample size. The cognitive and developmental outcome sample size calculations above determined the number of participants included for each test.

*Protocol addendum:* The protocol initially aimed to enroll 3800 participants, as indicated in the initial clinicaltrials.gov listing. This large sample size was chosen to detect a difference in anthropometric recovery of 3 percentage points with 95% sensitivity and 80% power. The sample size was reduced to 2700 children in March 2019, on the basis that available resources would not allow for the achievement of the original sample size.

### 9 Protocol deviations

#### 9.1 Food group stock-outs

In 2020, there were three stock-outs of study RUTFs due to ingredient importation restrictions related to the COVID-19 pandemic. These three periods were March 9, 2020 – March 31, April 8 - May 8 and November 9 – December 2. As a result of these stock-outs, randomization was unbalanced during these periods, with participants only randomized to the remaining available foods. One remote member of the study team was unblinded as a result of these periods. This unblinded individual did not evaluate any of the study subjects, nor did she analyze the primary outcomes.

The March 2020 stock-out occurred when the Malawi Bureau of Standards seized the study DHA stock due to exceeded expiration date 1 month prior. Following this, we obtained DHA oil from a donation from Wiley. On the following two pages the technical data sheet for this fish oil is presented. We used it as a direct replacement for the DHA in DHA-HO-RUTF.

The material was received packed in a metal barrel under argon as an air freight shipment, these precautions maintained its stability. When produced into RUTF in May-June 2020, ample amounts were made and promptly packaged to avoid problems with degradation of the omega-3 PUFAs from storing the opened barrel.

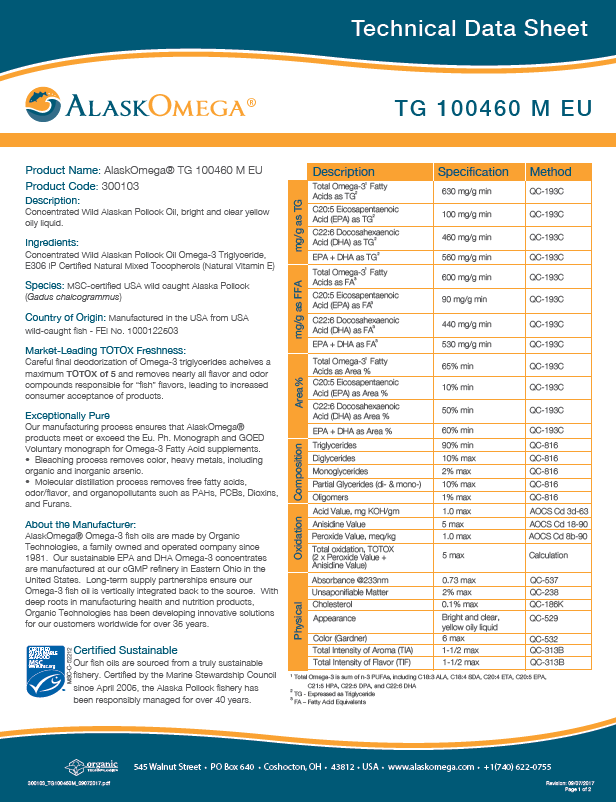

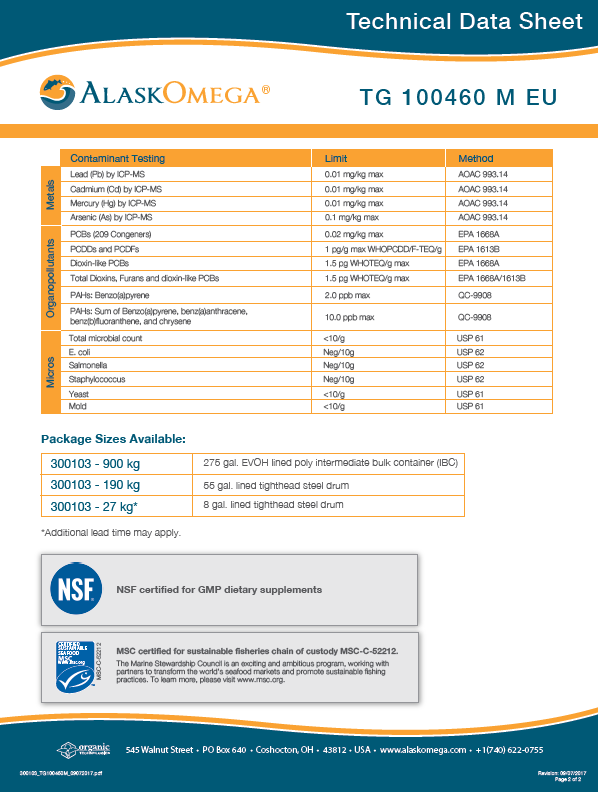

#### 9.2 Missing Problem Solving Assessment videos

On 12 August 2019 study staff discovered that an external hard drive that contained video recordings of study participants was missing from its storage space at our study office. The drive contained video recordings of infants participating in the problem solving assessment in the company of their mother or a care giver. The video files in the hard drive are linked to study site but not to participant ID. The staff in Malawi tore apart the office to search for the missing hard drive with no success in locating the hard drive. A total of 453 study participants videos files were completely lost as the files were too large to store on the field computers and they had not been backup on any other devices since March 2019. The PSA files lost were collected between end of March 2019 and 9 August 2019.

This incident was reported to all necessary institutional review boards and it was determined that the risk to the participant was minimal as the participant information that was contained in the hard drive is not linked to participant ID although the participant and caretaker faces are identifiable. The information in these videos, however, is non-sensitive in nature and deemed unlikely to cause significant social harm to the participants and their caretakers.

Following this incident, a series of corrective actions and protocols were put in place to avoid further data loss and ensure protection of study participants. These procedures included, password encrypting the external hard drives, keeping external hard drives stored in a digital code-restricted safe with limited access and video files would be uploaded to the College of Medicine Public Health Nutrition Group (COM PHNG) in Malawi server and uploaded to the Washington University cloud-based server on a weekly basis.

Documentation of this is available upon request.

### 10 Study pause

On 23 September 2019, we received notice from the institutional review board in Malawi, the College of Medicine Research Ethics Committee (COMREC), that the study’s approval had lapsed since April 2019 and that due to a misunderstanding of the College of Medicine field manager approval for study continuation had not been sought. As a result of this lapse in in approval and other citations during an audit conducted in August 2019, the study was suspended from enrolling participants until all items were addressed and study suspension lifted. On 25 November 2019, COMREC lifted suspension and approval was given to resume study enrollment.

Documentation of this is available upon request.

### 11 Procedures for neurocognitive testing

At the start of the study and periodically throughout, all neurocognitive assessors underwent training in the administration of the modified Willatts problem solving assessment, Malawi Developmental Assessment Tool, and the saccade eye-tracking evaluation by a qualified trainer.

#### 11.1 Modified Willatts problem solving assessment

After arriving at the clinic, the neurocognitive assessor would set up the problem solving assessment administration station in a room across from the clinic site that would allow for a sufficiently quiet environment for testing. The station included a table, two chairs, a supply box with toys, cloth covers, and a video camera to capture the entire room. As caregivers arrived, eligibility would be confirmed by comparison with a master list and the date would be cross-checked to confirm the child was meant to be tested that day. The assessor would discuss the child’s current health with their caregiver and if any concerns were raised, the caregiver would be directed to a study nurse and asked to return to the clinic at the next visit for testing.

The assessor would start by establishing rapport with the child, invoking the caregiver’s assistance. The goal was to establish a playful and comfortable environment for the child. The assessor would explain the testing procedure to the caregiver, including its purpose and the steps involved, as well as the need for recording the test. They would then instruct the caregiver to place the child on their lap facing forward across the table and to keep the child in this position as best they were able.

Next, the assessor would show the child three different toys, playing with them and offering them to the child to play, noting which one the child seemed to prefer best. The assessor would allow the child to play with the toys for at least 3 minutes to become comfortable, and would then provide the child with the toy they seemed to prefer. The assessor would then attempt to retrieve the toy from the child, asking them to return it and reaching out for it, and continue with playful attempts until they were successful in assuring the child’s cooperation.

Test 1: cloth and toy

The assessor would present the cloth to the child, moving it around the table and allowing the child to play with it until their interest waned. They would then bring up the toy and assure the child was attending to it, after which they would place the toy on the cloth and move them around the table together. Next, they would slide the cloth with the toy on top toward the child, placing it within 5 cm of the child’s edge of the table, and withdraw to their side. The assessor would then watch the child’s response. If the child did not reach for the cloth to pull it and the toy toward them to grab the toy, the assessor could say general phrases such as “go ahead,” “you can do it,” or “it’s ok,” but not specific phrases like “grab the toy.” The child was given 30 seconds for the task. If the child performed the problem solving maturely, the assessor could move on to problem 2. If they did not, the assessor could repeat problem 1 one more time at most before moving on to problem 2 regardless of success.

Test 2: cover and toy

The assessor would present the cover to the child in the same manner as above. They would then bring up the toy and, confirming the child was watching them, would place the toy under the cover. They would then gently slide the cover and toy toward the child to within 2 cm of the child’s edge of the table, with the toy under the middle of the cover, before withdrawing to their side. The child was given 30 seconds for the task. If the child readily removed the cover and grabbed the toy, the assessor would then move on to problem 3. If the child did not readily react, similar phrases as above could be used for encouragement, without direct instruction. The assessor could repeat problem 2 one more time at most. Reasons for repeating could be lack of interaction with cover or toy, knocking both off the table, seeming to swipe at both and accidentally uncovering the toy, or if there was a long delay between the presentation of cover and toy and the child’s action. Assessors would stop at problem 2 only if the child did not succeed on either problem 1 or problem 2.

Test 3: cloth, toy, and cover

The assessor would then present the cloth and place the toy on top, confirming the child was watching them. They would then place the cover on top of the toy, again confirming the child has seen, before sliding the cloth/toy/cover over to the child, within 5 cm over the child’s edge of the table. If the child solved the problem in the mature manner (readily pulled the cloth toward themself, removed the cover and grabbed the toy), the assessment was complete. If the child did not readily solve the problem, words of encouragement could be used similar as those above, without specific direction or pointing. If the child did not solve the problem in a mature manner, the assessor could repeat problem 3 at most one more time.

The tests were video recorded and sent to the co-principal investigator at Cornell University, who oversaw training of coders there. Coders were required to complete training and achieve 90% agreement by kappa statistic on test video intention scores before coding for the trial. Using software, Behavioral Observation Research Interactive Software (BORIS), assessors coded tests and tabulated intention scores. In a modification of the procedure of Willatts et al,^70^ problems were coded for two primary components, each on a scale of 0-2 and each assessing an aspect of intentionality: orientation and execution. Orientation was defined as the quality of shift in attention (visual fixation, bodily orientation) from the toy itself to the subgoal in question (e.g., cloth to pull closer, or cover to remove), or means by which the ultimate goal could be obtained. Orientation was scored on three levels: direct shift, slow/indirect/fumbling shift, and no shift. Primarily in older children, it was possible for a child to demonstrate a phenomenon known as “covert attention,” in which the child executed the sub-goal without clear change in visual fixation or body position, or did so too fast for observation. If the execution was smooth/perfect, the child would receive credit for orientation. Execution is defined as the manner in which the child performs motor actions on the task materials when confronted with the barrier, including how they manipulate subgoals. Execution is scored on three different levels: smooth/perfect execution, fumbling execution, or no execution.

For problems 1 and 2, intention scores ranged from 0-4. For problem 3, intention scores ranged from 0-8.

#### 11.2 Malawi Developmental Assessment Tool

After arriving at the clinic, the neurocognitive assessor would set up the MDAT administration station, often in a room across from the clinic site that allowed for a sufficiently quiet environment for testing. As caregivers arrived, eligibility would be confirmed by comparison with a master list and dates would be cross-checked for accuracy. The assessor would discuss the child’s current health with their caregiver and if any concerns were raised, the caregiver was directed to a study nurse and asked to return to the clinic at the next visit for testing. Included as an appendix is a list of all the MDAT assessments.

The assessor would open the tablet and enter their initials, participant ID, date of collection, date of birth of the participant, and caregiver details. The questionnaire automatically shifted to the appropriate starting point for the participant’s age and the assessor would proceed item-by-item. The MDAT contained 136 items in total. For evaluation of gross motor, fine motor, and language development items, the assessor would directly observe the child. Social development is evaluated by questions asked of the caregiver. Each item was scored as “pass,” “fail,” or “NA.” “NA” was used if the child did not engage sufficiently for the assessor to make a determination. If a participant scored “fail” on six consecutive items in any single domain, the questionnaire would automatically change to the next domain.

#### 11.3 Saccade eye-tracking assessments

The setup for the eye-tracking assessment required several steps to be completed before participants arrived. The laptop, monitor, external processing unit, and webcam all were connected to power, the external processing unit was connected to the eye-tracker and laptop, the laptop was connected to the monitor and webcam, and the study USB drive was connected to the laptop. The software was password protected. The assessor then mapped the display monitor and eye tracker such that they aligned spatially.

Next, the caregiver and child would be brought into the testing station. Their ID, name, visit, the caregiver’s name, and assessor ID would be written on the whiteboard and the participant’s ID was entered into the computer. The assessor would align the lab window and webcam window so they were able to see both the video of the child and the video of the eye-tracking test simultaneously.

Next, the assessor would explain the process and purpose of testing to the caregiver. The caregiver would be asked to adjust their baby carrier so the child faced forward and sit in the study chair. The assessor would then hold the whiteboard in front of the camera, click record, and read the information aloud, before removing the whiteboard from view. The first video would be played to attract the child’s attention, and different videos could be tried until the child was seen fixing their attention on the screen. The caregiver was then asked either to look away or close their eyes, and the assessor would adjust the screen to be at eye level and 55-65cm away from the child’s face.

The assessor would then initiate the eye-tracking calibration, which aimed to match where images appeared on the monitor and where the eye-tracking software calculated their presence. The software first calculates the distance of the child’s eyes from the monitor, following which the assessor would adjust the monitor to be as close to 60 cm away as possible. The assessor would then click “start calibration,” which presented an image at pre-specified locations. The assessor compared the known image location with eye fixation and adjusted the screen as needed, re-calibrating after each change. There were multiple videos available for calibration if the initial options did not work.

Next, the assessor would begin testing by showing an “attention grabber,” meant to focus the child’s eyes on the center of the screen. They could shake a rattle behind the screen if the child would not attend there but could not use a rattle or any other external stimulus except during an “attention grabber” step. The software would then run through testing with interspersed attention grabbers. The recording could be stopped if the child was not able to attend to the screen.

Testing included visual paired comparison task and infant orienting with attention task. Identical procedures to those previously described were used.^69^ The visual paired task consisted of 4 trials of African faces. The infant orienting task measured saccadic reaction time in a standardized manner. The testing sequence for paired comparison task and orienting attention task was randomized.

After the testing was complete, the assessor would show a sample video to the caregiver to explain what was done. They would release the caregiver and then save the file in a prespecified format. The assessor would then complete a form that included information about the child’s perceived mood and activity level and contained a free comments section where they could record technical issues or anything else unusual that occurred during testing.

12 **Biochemical fatty acid profile analysis**

To evaluate the influence of study RUTF formulations on circulating fatty acid profiles, gas chromatography-flame ionization detection (GC-FID) of fatty acid methyl esters (FAME) derived from isolated plasma phospholipids was carried out.

At the 4-week study visit, blood samples for fatty acid analysis were collected into ethylenediaminetetraacetic acid treated vacutainer tubes. Plasma was separated from red blood cells through centrifugation at 1000-2000 x g for 10 mins, following which it was transferred to clean tubes and held at -20°C until samples were received at the Dell Pediatric Research Institute.

The choice of analysis of plasma fatty acids levels over other fractions, such as whole blood or red blood cells, was twofold. Firstly, plasma phospholipid DHA levels reach a plateau after approximately 4 weeks of supplementation or feeding.^31^ Second, plasma fatty acids are stable for over 1 year when stored at -20°C, whereas whole blood and red blood cell PUFA are liable to degradation by iron initiated peroxidation when stored at -20°C.^71^

From plasma collected at 4-week blood draws, phospholipids were isolated using a three-phase liquid-liquid extraction similar to the methods described by Vale et al. and adapted in house for semiautomated extractions by a Hamilton Microlab STARlet robot (Hamilton Robotics, Reno, NV).^72^ Briefly, an aliquot of plasma is weighed out, to which a mixture of hexane: methyl acetate: acetonitrile: water (4:4:3:4, by volume) is added. Samples are mixed then incubated at room temperature for one hour. The middle phospholipid containing phase is then collected into a new tube, dried under nitrogen, and reconstituted in heptane.

After isolation of plasma phospholipid, samples underwent transmethylation to generate FAME for analysis by GC-FID. Transmethylation is required to cleave fatty acids from their esterified position on the glycerol backbone of their native phospholipid and results in a volatile derivative with good chromatographic properties. For the purposes of this study, isolated plasma phospholipids were transmethylated via a one-step reaction adapted from the methods of Garcés and Mancha.^73^ Briefly, a mixture of methanol: 2,2-dimethoxypropane: sulfuric acid (85:11:4, by volume) to the solvated phospholipid, followed by a mixture of heptane: toluene (63: 37, by volume). Samples are then heated at 80°C for 2 hrs. A saturated solution of NaCl is then added, and samples are incubated at 4°C for 3 hrs. The top FAME-containing layer is then transferred to a clean glass tube, evaporated under nitrogen, and reconstituted in heptane for analysis by GC-FID.

Gas chromatography is a classical analytical technique routinely used for the analysis of fatty acids. A complex mixture of FAME is injected onto a capillary column coated with a stationary phase. An inert carrier gas serves as a mobile phase as gaseous compounds interact with the stationary phase of the capillary column, and specific analyte species elute off of the column with a characteristic retention time. With respect to FAME, species are separated based on carbon chain length and degree of unsaturation. The area of the corresponding chromatographic peaks are related to the abundance of FAME in the sample and are put on a quantitative basis using experimentally determined response factors.

For the proposed study, FAME were separated on a 20 m $\times$ 0.22 mm i.d. $\times$ 0.25 µm film thickness BPX70 capillary column (Trajan, Austin, TX) configured in a Shimadzu GC2010 equipped with a flame ionization detector (Shimadzu Corporation, Kyoto, Japan)^74^ Samples were injected into a splitless inlet held at 250°C using a Shimadzu AOC-20i/s autosampler. FAME were separated using a GC temperature program as follows, 80°C hold for 2 min, 10°C/min to 174°C hold for 2 min, 10°C/min to 240°C hold for 10 min, and 5°C/min to 245°C hold for 5 min. The flame ionization detector was held at 260°C. Peak area response factors were generated from a reference mixture of equal weight FAME (Nu-Chek Prep, Inc., Elysian, MN) and applied to peak areas under the curve to yield calibrated FAME profiles. FAME structural identity is positively identified by covalent adduct chemical ionization mass spectrometry^75^. Fatty acid data will be reported as weight percentages (%wt) of total fatty acids identified. A complete profile of 25 fatty acids will be collected for each sample.

The test formulations of RUTF, HO-RUTF, and HO+DHA-RUTF were modified to enhance the distribution of omega-6 and omega-3 fatty acids. Moreover, the use of high-oleic peanut varieties as the base ingredient contributes to increasing the abundance of oleic acid in study RUTF. As such, the plasma phospholipid fatty acids we propose to compare across dietary treatments are oleic acid, omega-6, and omega-3 series fatty acids. Specific fatty acids of interest are indicated and described below:

12.1 **Oleic acid**

Oleic acid is an 18-carbon monounsaturated fatty acid found abundantly in vegetable oils, notably olive oil. Within the brain, oleic acid is found predominately in white matter where it comprises over 30% of the total fatty acids.^18^ Oleic acid is a critical component of brain myelin which is rapidly accumulated following birth.^23^

12.2 **Omega-6 series**

**Linoleic acid** an 18-carbon PUFA containing double bonds at carbon nine and twelve from the carbonyl end. Humans lack delta-12 desaturase and consequently are unable to endogenously synthesize LA, lending to LA being considered an essential fatty acid. Beyond acting as a precursor for longer chain omega-6 polyunsaturated fatty acids, such as ARA, LA is incorporated into the brain where it plays a role in the neuroinflammatory processes through the production of oxidized linoleic acid metabolites (OXLAMs).^76^ Experimental evidence from rodent studies suggests that diets with high LA contribute to a greater pro-inflammatory response to brain insult.^77^ In humans, reducing dietary LA intakes in combination with increasing EPA and DHA intake is associated with reduced headache duration and severity in patients with chronic daily headaches.^76,78,79^

**Arachidonic acid** is a 20-carbon omega-6 PUFA synthesized from the desaturation and elongation of LA, primarily taking place in the liver. During the first two years of life, ARA is the most abundant PUFA in the brain.^23,80^ Over 90% of ARA in the brain is esterified into phospholipids; however, free-ARA is released in response to the activation of dopaminergic, cholinergic, serotonergic, or N-methyl D- aspartate (NMDA) activation.^21^ Free-ARA can be metabolized by cyclooxygenases, lipoxygenases, and cytochrome P450 to produce bioactive eicosanoids which have been shown to mediate neuroinflammatory pathways.^21,77,81^ Additionally, ARA is necessary for the synthesis of the two most abundant endocannabinoids in the brain, anandamide and 2-arachidnoyl glycerol, which have been shown to regulate synaptic function.^21^

**Adrenic acid** (AdrA) is the 22-carbon omega-6 PUFA synthesized from elongation of ARA. It is present in various tissues including the brain and its concentration increases in the first years of life.^23^ It particularly increases when DHA is low.

**Docosapentaenoic acid** omega-6 (DPA6) is the 22-carbon omega-6 PUFA synthesized from desaturation of AdrA. DPA6 is a structural analog of DHA, with the sole difference being that DPA6 has one fewer double bond than DHA. Because DPA6 can be made from dietary LA, its concentration increases in DHA deficiency and it has long been known as a marker for DHA deficiency.

12.3 **Omega-3 series**

**Alpha-linolenic acid** is the 18-carbon essential omega-3 PUFA. As is the case with LA, humans do not possess the required desaturase enzymes for *de novo* synthesis of ALA; therefore, it must be supplied by the diet. As a precursor to longer-chain omega-3 PUFA such as eicosapentaenoic acid (EPA, 20:5n-3) and DHA, ALA competes with LA for elongase and desaturase enzymes. Excess dietary intakes of LA have been shown to suppress circulating DHA levels from ALA by outcompeting ALA for elongase and desaturase enzymes.^13^ Although ALA rapidly enters the brain, it does not appreciably contribute to local within-brain production of DHA.^25^ Despite sharing a similar incorporation rate as DHA, brain ALA levels are maintained very low (≤0.5% of total fatty acids) likely due to rapid β-oxidation.^25^ ALA has been proposed as a potential substrate for the production of ketones within the brain, which may serve as an important energy source to support rapid brain growth and development during infancy and young childhood.^82,83^

**Eicosapentaenoic acid** is a product of ALA elongation and desaturation. Despite being relatively abundant in the blood, EPA levels are exceedingly low in the brain, about 250-300 fold lower than DHA levels.^84^ Similar to ALA, EPA is rapidly β-oxidized after entering the brain. Although levels are maintained very low, EPA has been suggested to play a role in the regulation of mood.^21^ Results from randomized clinical trials suggest that supplementation with EPA dominant (EPA ≥ 60% of EPA+DHA) omega-3 is effective against primary prevention of major depression.^85^ Additionally, EPA is converted to oxygenated metabolites and specialized pro-resolving mediators that have been shown to promote the resolution of inflammation following brain injury.^84,86^

**Docosahexaenoic acid** is the most abundant PUFA in the brain beyond about 2 years of age, and has been shown to play an important role in the regulation of neuroinflammation, cell survival, synaptogenesis, and signal transduction.^20,21^ Increasing levels of DHA in the blood and tissues can only be achieved through either decreasing the intake of LA while maintaining an adequate intake of ALA, or consuming dietary preformed DHA.

### 13 General analysis considerations

#### 13.1  Data cleaning

All data were double-entered into password-protected Microsoft Access databases and differences will be reconciled by reassessing the original data cards and using these as the authoritative version.

#### 13.2  Outliers

Extreme values of anthropometrics and z-scores will be evaluated individually and assessed for obvious recording errors. If no error is found, previously identified criteria will be used to exclude these outliers: 20 cm < MUAC < 6.5 cm, weight change > 25g/kg/day, or MUAC change > 1.5 cm per week.^87^

Extreme MDAT z-scores were evaluated individually and assessed for obvious recording errors. Outliers that were clearly impossible would be corrected if possible or recoded to missing if correction was not possible. Based on observation of the distribution, global MDAT z-scores <-5 were excluded.

#### 13.3  Framework

All outcomes will be tested using a two-sided superiority framework.

#### 13.4  Timing of analyses

We will conduct all analyses blinded to intervention groups by creating a code for actual group assignments. The final analysis will be performed after all study participants have achieved a programmatic outcome and all data have undergone cleaning.

#### 13.5  Confidence intervals and p-values

In all analyses, confidence intervals will be two-sided and calculated at the 95% confidence level, and all testing will be two-sided and considered significant at the p < 0.05 level.

#### 13.6  Analysis Populations

13.6.1 **Modified Intention-to-treat (ITT)**

This population will include all randomized study subjects with one exception: all participants in whom a pre-existing exclusion criterion is found after randomization will be excluded from the study and the modified intention-to-treat analysis. This is justified by the blinded nature of the experiment and the type of exclusion criteria in question – determining age rounded to nearest month < 6 or > 59 mo, presence of a chronic debilitating medical condition, vision or hearing problem – which are unlikely to be affected by allocation group even in the absence of blinding. All superiority analyses will be performed as modified intention-to-treat.

13.6.2 **Protocol deviation subgroup**

We will perform a secondary analysis including only those participants who were enrolled outside the windows of the three food group stock-outs that led to unbalanced randomization. We will characterize the enrollment characteristics of these children and repeat the primary and key secondary analyses to assess for bias introduced by unbalanced randomization.

#### 13.7  Covariates and Subgroups

Multiple variables are hypothesized to exert an influence on neurocognitive assessments.

- Child age at the time of assessment
- Child sex
- Type of SAM (kwashiorkor, marasmus, marasmic-kwashiorkor)
- Enrollment HAZ and WAZ
- Presence of active infection: reported days with fever, diarrhea

The above list will be assessed but is not meant to be exclusionary such that the study investigators may generate additional observations and hypotheses during the study.

#### 13.8  Missing Data

The percent of participants lost to follow-up between enrollment and outcome assessment will be tabulated.

Missing data will be identified using summary statistics and quantified using % of participants with the missing variable, and % of measurements, where applicable. Missing data will not be imputed. In some cases, if the missing data are essential for inclusion in the analysis, e.g., date of birth for age-based z-score calculations, these participants will not be included in these analyses.

#### 13.9  Multiple Testing

There are two primary endpoints in the study and three intervention arms. To reduce the family-wise error rate, only two pairwise comparisons will be made each for the MDAT z-score and problem solving assessment scores, HO-RUTF vs. S-RUTF and DHA-HO-RUTF vs. S-RUTF. These two comparisons have been chosen because of their primary importance for the study hypothesis.

Multiple secondary endpoints and analyses will be completed, but we will not plan to generate *P*-values for these comparisons, but rather differences with 95% confidence intervals.

13.10 **Reporting conventions**

*P*-values ≥ 0.001 will be reported to 3 decimal places. *P*-values less than 0.001 will be reported as < 0.001. Percentages will be reported to the tenths digit. Z-scores will be reported to the hundreds digit. Estimated parameters, such as regression coefficients, will be reported to 3 significant figures. Confidence intervals will be 95%.

### 14 Subject disposition

The CONSORT flow diagram (skeleton below) will detail the number of children at enrollment, allocation, follow-up, and analysis stages, with details of exclusions between each stage listed.

Assessed for eligibility (n = )

Randomized (n = )

Allocated to HO-RUTF (n = )

Excluded (n =	)

- Under 6 months of age (n = )
- Over 59 months of age (n = )
- Transfer from inpatient nutrition unit (n = )
- Recently treated as outpatient for acute malnutrition (n = )
- Obvious chronic/debilitating illness (n = )
- Unable to obtain consent (n = )
- Not a resident of catchment area (n = )

#### Enrollment

#### Allocation

Allocated to HO-RUTF (n = )

Allocated to HO-RUTF (n = )

#### Follow-Up

Same as central branch

Same as central branch

Subsequently excluded (n = )

- Under 6 months of age (n = )
- Over 59 months of age (n = )
- Incomplete data (n = )
- Actually moderately malnourished (n = )
- Failed test feeding (n = )
- Received incorrect food (n = )
- Transfer from inpatient hospital nutrition unit (n =)

#### Analysis

Analyzed (n = )

- Recovered (n = )
- Failed (n = )
  - Died (n = )
  - Hospitalized (n = )
  - Remained acutely malnourished (n = )
  - Default (n = )

### 15 Demographic and derived variables

#### 15.1 Baseline variables

| **Category** | **Definition** |
| --- | --- |
| **Demography** |  |
| Age | Child age in months; continuous |
| Sex | Male or female; categorical |
| Child still being breastfed | Yes or no; categorical |
| Caregiver relation to child | Mother, father, sibling, grandparent, aunt, uncle |
| Mother alive | Yes or no; categorical |
| Father alive | Yes or no; categorical |
| Number of siblings of participant | Discrete |
| Number of siblings of participant that have died | Discrete |
| Number of radios in the home | Discrete |
| Number of bikes in the home | Discrete |
| Latrine use | Yes or no, categorical |
| Water source | Borehole, well, or river; categorical |
| How many times per day water retrieved | Discrete |
| Animals sleep in the home | Yes or no; categorical |
| Number of individuals that sleep in the same room as the child | Discrete |
| Material of household roof construction | Thatch or metal sheets; categorical |
| **Health questionnaire** |  |
| Child with good appetite | Yes or no; categorical |
| Presence of fever in past 2 weeks | Yes or no; categorical |
| Days of fever in the past week if present | Discrete |
| Presence of diarrhea in past 2 weeks | Yes or no; categorical |
| Days of diarrhea in the past week if present | Discrete |
| Participant had HIV testing done | Yes or no; categorical |
| If above yes, result | Positive or negative; categorical |
| If positive, on antiretroviral therapy | Yes or no; categorical |
| Caregiver had HIV testing done | Yes or no; categorical |
| If above yes, result | Positive or negative; categorical |
| If positive, on antiretroviral therapy | Yes or no; categorical |
| Participant on tuberculosis treatment | Yes or no; categorical |
| Adult in participant’s home on tuberculosis treatment | Yes or no; categorical |
| **Adherence** |  |
| Child eating RUTF well | Yes or no; categorical |
| **Gross motor development questionnaire** |  |
| Sitting without support | Yes or no; categorical |
| Hand-and-knees crawling | Yes or no; categorical |
| Standing with assistance | Yes or no; categorical |
| Walking with assistance | Yes or no; categorical |
| Standing alone | Yes or no; categorical |
| Walking alone | Yes or no; categorical |
| **Anthropometry** |  |
| Weight | Measured to nearest 5g; continuous |
| Length | Measured to nearest 2mm; continuous |
| MUAC | Measured to nearest 1mm; continuous |
| Weight-for-height z-score | Continuous |
| Weight-for-age z-score | Continuous |
| Length-for-age z-score | Continuous |
| Target weight | Calculated using z-score chart; continuous |
| Edema | Present or absent; categorical |
| Edema severity | 1-3+; discrete |

#### 15.2 Derived variables

- Weight gain will be calculated as g/kg/d over the treatment course for all children for whom data is available. MUAC gain and length gain will be total mm/wk over the treatment course for all children for whom data is available.
- Because the two intervention arms have overlapping components, i.e. the DHA-HO-RUTF arm contains DHA in addition to HO (the identifying component of the HO-RUTF intervention), in a secondary analysis, we will create variables for DHA and HO for regression analyses rather than a variable for the intervention group.
- We will assess the distribution of saccade reaction time results, which have been identified as right-skewed in prior analyses. If we identify a right-skewed distribution, we will attempt transformations, first log, then square root, and assess for improvement in distribution symmetry.

### 16 Outcome analysis

#### 16.1 List of outcomes with variable type

| **Primary** |  |
| --- | --- |
| Problem solving assessment intention scores | Ordinal |
| Malawi Development Assessment global z-score | Continuous |
| **Key secondary outcomes** |  |
| Recovery | Yes or no; categorical |
| MDAT sub-domain z-scores | Continuous |
| **Other secondary outcomes** |  |
| **Neurocognitive outcomes** |  |
| Saccade eye-tracking assessment score | Continuous |
| **Anthropometry** |  |
| Weight change | g/kg/d, continuous |
| Length change | mm/wk, continuous |
| MUAC change | mm/wk, continuous |
| **Programmatic Outcomes** |  |
| Recovery | Yes or no; categorical |
| Death | Yes or no; categorical |
| Default | Yes or no; categorical |
| Remained malnourished | Yes or no; categorical |
| Transferred to hospital | Yes or no; categorical |
| **Biochemical testing, serum levels** |  |
| Oleic acid | %; continuous |
| Docosahexaenoic acid | %; continuous |
| Eicosapentaenoic acid | %; continuous |
| Alpha-linolenic acid | %; continuous |
| Linoleic acid | %; continuous |
| Arachadonic acid | %; continuous |
| Docosapentaenoic acid | %; continuous |
| Adrenic acid | %; continuous |

#### 16.2 Outcome variable definitions

16.2.1 **Primary outcomes**
- MDAT global z-score for gross motor, fine motor, language, and social domains 5-7 mo after completion of SAM treatment.
  - We will calculate the raw score on each MDAT sub-domain as the number of items the child was observed or reported to be able to perform out of 38 gross motor items, 34 fine motor items, 35 language items, and 29 social items.
  - We will then calculate global and sub-domain z-scores using the standard published norms.
  - For children in whom one sub-domain is unable to be completed, the remaining sub-domains will be used to calculate the global z-score.
  - We will require at least two sub-domains to be completed for a global score to be included in analysis
- Problem solving assessment intention scores
  - These scores are ordinal. The distribution was unknown prior to the study because the age range was wide and the children had recently been treated for SAM. We considered the possibility of combing the three problem scores into a composite score, but the meaning of each problem is different, specifically problem 3 is more than a mere combination of problems 1 and 2, and so each problem will be analyzed separately.
  - Problems 1 and 2 had scores of 0-4, while problem 3 was scored 0-8.
  - NA was scored when children were unable to engage with the problem sufficiently for a score to be obtained, and the reasons for NA outcome were documented.
    16.2.2 **Secondary outcomes**
- MDAT sub-domain z-scores
- Recovery: recovery from SAM was defined based on the criteria used for diagnosis:
  - Recovery must have occurred on or before the 6^th^ follow-up visit.
  - In those with nutritional edema only, recovery was the resolution of nutritional edema for two consecutive follow-up visits.
  - In those with WHZ < -3, a target weight was calculated on enrollment using the child’s length and a WHZ chart separated by sex, such that obtaining this weight would result in WHZ ≥ -2. Recovery was determined when the child reached the target weight.
  - In those with MUAC < 11.5 cm, recovery was be determined when MUAC ≥ 12.5 cm
  - In those with nutritional edema and WHZ/MUAC criteria, recovery was defined by resolution of nutritional edema for two consecutive visits and reaching either of the WHZ or MUAC criteria above.
  - In those with WHZ < -3 and MUAC < 11.5 cm, recovery was determined when the child reached either WHZ or MUAC criteria for recovery
- Default: determined when a child missed three consecutive visits
- Remained malnourished: the child did not reach an alternative outcome (recovery, transfer to hospital, death, default) and remained malnourished after 6 follow-up visits
- Death: caregiver reported the child died
- Transfer to hospital: criteria for uncomplicated SAM no longer reached and field team determined child required transfer to inpatient nutritional rehabilitation unit
- Saccade eye-tracking assessment score: automated output of the program, in units of milliseconds

#### 16.3 Primary outcome analysis

There are two primary endpoints to be compared in this study: MDAT global z-score and the modified Willatts problem solving assessment intention scores. The null hypothesis for MDAT z-score will be that the sample means are equal between DHA-HO-RUTF vs. S-RUTF and HO-RUTF vs. S-RUTF. The null hypothesis for the problem solving assessment intention scores will be that the odds ratios for higher scores will be equal between DHA-HO-RUTF vs. S-RUTF and HO-RUTF vs. S-RUTF. Mean differences and 95% CIs will be calculated for pairwise comparisons in MDAT z-score, while ORs with 95% CI will be calculated for pairwise comparisons in problem solving assessment intention scores.

16.3.1 **Global MDAT z-score**

Student’s *t* test will be used to compare global MDAT z-scores. Differences in mean scores with 95% CIs will be reported alongside *P-*values.

16.3.2 **Problem solving assessment scores**

Age is a known predictor of problem solving assessment results and so will be included *a priori* as a covariate in the regression analysis. We will assess for sparse cells and, if present, will combine groups to satisfy this assumption. We will test for proportional odds using graphical comparison of the beta coefficients each group resulting from binary logistic regressions at cut-points of scores >2. An interaction term between age and group will be offered into the model and included if it demonstrates a statistically significant effect. Multicollinearity is not expected, as only the intervention group and age will be entered as covariates into the regression model, and the intervention group is randomly assigned. Odds ratios with 95% CIs will be reported alongside *P*-values.

#### 16.4  Secondary outcome analysis

Programmatic outcomes will be summarized as n (%). Anthropometric growth rates will be assessed for distribution using histograms; if normally distributed, they will be summarized with mean and SD. If the data do not appear normally distributed, they will be summarized as median (IQR). Mean differences and 95% CIs will be calculated for all continuous variable comparisons. Proportion differences and 95% CIs will be calculated for all categorical variable comparisons.

16.4.1 **Neurocognitive outcomes**

Mean MDAT sub-domain z-scores will be compared across all pairs using Student’s *t* test. Saccade eye-tracking assessment score will be compared across all pairs using Student’s *t* test. Differences with 95% CIs will be reported.

16.4.2 **Biochemical outcomes**

Fatty acid percentages will be assessed for distribution shape. If they do not appear to approximate the normal distribution, they will be compared using Wilcoxon rank sum test; if they do approximate the normal distribution, they will be compared using Student’s *t* test.

16.4.3 **Programmatic outcomes**

Chi-square will be used to compare proportions who have recovered, remained malnourished, died, and defaulted between DHA-HO-RUTF and S-RUTF, and between HO-RUTF and S-RUTF. Differences with 95% CIs will be reported.

16.4.4 **Anthropometric outcomes**

Rates of gain in weight, MUAC, and length will be compared using Student’s *t* test. Differences with 95% CIs will be reported.

16.4.5 **Analysis by intervention component**

Linear regression will be performed to compare the effects of HO and DHA on MDAT global z-scores. Beta coefficients with 95% CI will be reported. Ordinal logistic regression will be performed to compare the effects of HO and DHA on the PSA composite intention score. ORs and 95% CI will be reported.

### 17 Safety Analyses

There were no predicted safety issues with either of the experimental fatty acid formulations of RUTF used in this study. Following study completion, we will compare adherence, defaulting, and symptoms, including diarrhea and fever, between DHA-HO-RUTF and S-RUTF, and between HO-RUTF and S-RUTF.

### 18 Quality assurance of statistical programming

Windows operating system will be used for statistical analysis. Data will be stored in locked Microsoft access databases until analysis. Data cleaning will be performed using Microsoft Excel and SPSS. At the time of writing, we plan to perform analyses using R and SPSS. All R syntax will be dated, saved, and stored.

### 19  Attachments

19.1 **Consent document**

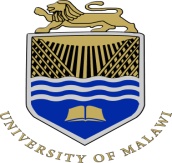

**COLLEGE OF MEDICINE**

**Principal	College of Medicine**

**M. H. C. Mipando MSc PhD	Private Bag 360**

**Chichiri**

**Blanytre 3**

**Our Ref:	Malawi**

**Your Ref:	01 874107**

“Randomized, double-blind, placebo-controlled trial to evaluate the effectiveness of different fats in food in the community-based treatment of severe acute malnutrition in Malawian children”

Your child is invited to participate in a research study conducted by Dr. Maleta and colleagues.

1. The overall purpose of this research is to see if your child will grow and gain weight while achieving a more normal neurocognitive status when using therapeutic foods with different fats as he/she recovers from severe malnutrition.

2. You will be given one of three peanut butter foods to feed him/her at home. One of these foods is the standard RUTF, the other two RUTFs are experimental. They contain all of the same nutrients that are recommended by WHO, and include some different fats. One of the fats is from a different variety of peanut and the other from fish oil. You will be given enough food to feed the child for 2 weeks, and asked to return every 2 weeks to be weighed, measured and given more food. You will be asked to feed your child this food until his/her weight has returned to what is considered normal for the child’s height.

After 4 weeks of treatment a small blood sample, 1.5 mL will be taken from his/ her arm to measure the amount of essential fatty acids in the body.

After 6 weeks, if your child is one of the following ages, 10-11 mo, 14-15 mo or 18-19 mo he/ she will be asked to participate in a special test. The test has two parts and takes about 30 minutes total. The first part of the test is conducted as you sit with your child in your lap watching a computer screen. The computer will show your child some pictures and determine how long the child looks at any one picture and how quickly your child looks at new pictures. The second part of the test involves showing your child an interesting object that is enclosed in a plastic box with a hole in it. Your child will try to remove the object from the box with his/ her hand. This exercise will be recorded by a video camera. Experts will review these videos and determine how well your child was able to remove the interesting object. Both of these exercises are known to be safe.

A 6 months after recovery and when your child reaches about 3 years of age, he/ she is asked to return to participate in a testing exercise in which the child is asked to complete certain small tasks and solve certain small problems. This test takes about 30-60 minutes. Three types of tests will be conducted namely: pattern analysis which involves a set of form-board problems testing visual-constructional abilities; copying which involves a set of block building problems also testing visual-constructional abilities and bead memory involving a set of visual memory tests.

Your (your child’s) participation in this study is expected to last 8 weeks of therapy.

Other malnourished children may benefit because we may learn whether these different therapeutic foods are necessary to promote full nutritional recovery from severe malnutrition.

6. Other than non-participation in the research, there are no alternatives. If you decide not to participate, your child will still receive the peanut butter food without any extra fats to help him/her recover from the malnutrition.

7. All reasonable measures to protect the confidentiality of your child’s medical records. There is a possibility that your child’s medical record, including identifying information, may be inspected and photocopied by government or University officials or members of the Human Studies Committee.

8. You will receive soap at every 2-week follow-up visit and a chitenje upon graduation.

9. If you have any questions or concerns regarding this study, if any problems arise, or there any feelings of pressure to participate, you may call the Principal Investigator Dr. Maleta at College of Medicine, Blantyre on 0888 232 202. You may also ask questions or state concerns regarding your (your child's) rights as a research subject to Dr. YB Mlombe at COMREC’s secretariat, College of Medicine on 01 871 911.

10. University of Malawi investigators and their staffs will try to reduce, control, and treat any complications from this research. If you feel that you are injured because of the study, please contact the Investigator and/or the Human Studies Committee Chairman.

11. You (your child) will be informed of any significant new findings developed during the course of participation in this research that may have a bearing on your (your child's) willingness to continue in the study. The investigator may withdraw you (your child) from this research if circumstances arise (such as non-compliance with the protocol and non-tolerance of a study food) which warrant doing so.

12. This research is not intended for the purpose of diagnosing or treating any medical problems not specifically stated in the purpose of the research.

I have heard this consent and have been given the opportunity to ask questions. I will also be given a copy of this consent form for my records. I hereby give my permission (or give permission for my child) to participate in the research described above,

Parent’s name: _____________________Parent’s signature/thumbprint:__________	Date:________

Name of child ___________________

Witness’ name: _____________________Witness’ signature/thumbprint:__________	Date:________

Study staff’s name:___________________study staff’s signature:_________________	Date:________

19.2 **WHZ charts**

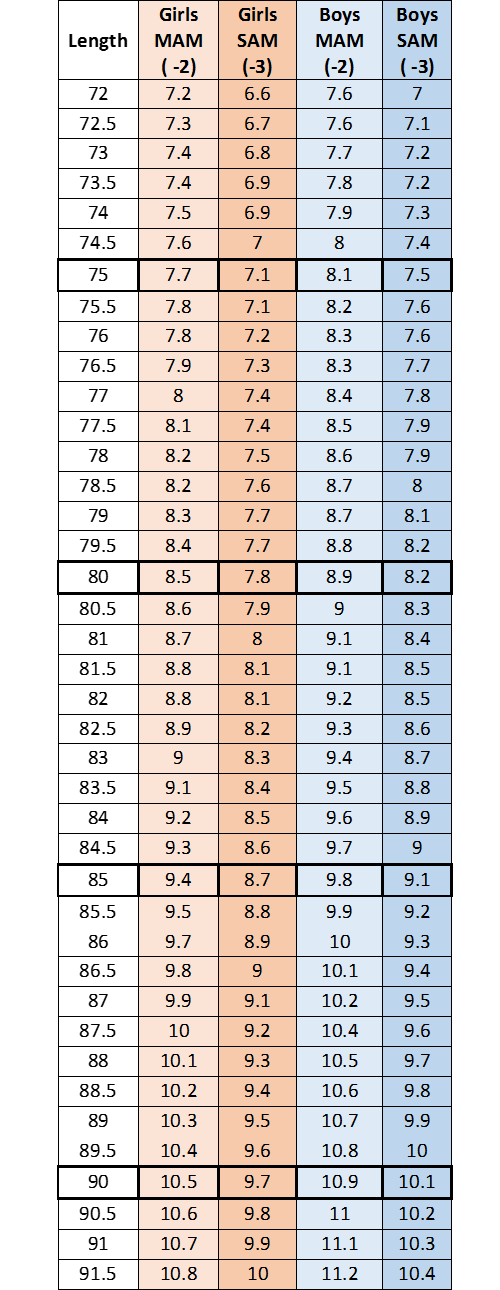

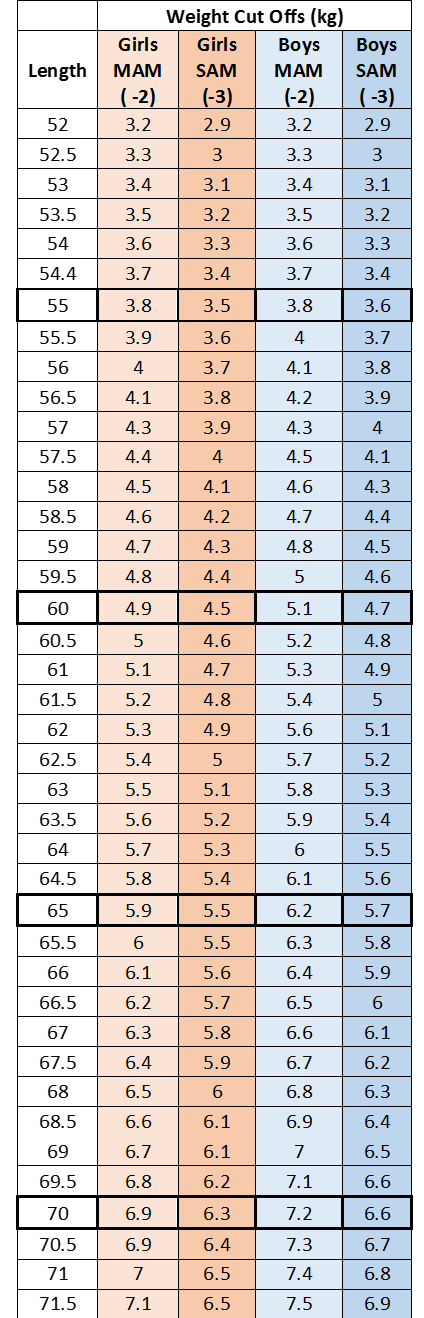

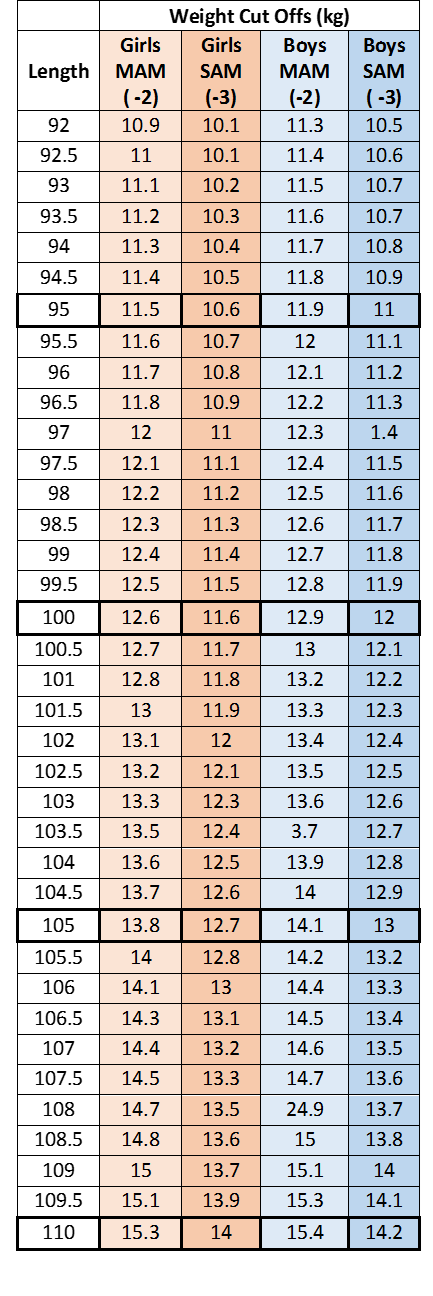

19.4 **MDAT items sheet**

**GROSS MOTOR**

| **1. Lifts chin up off the floor for a few seconds***  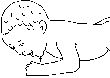 | 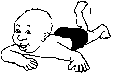**2. Prone (on his or her tummy), can lift head up to 90 degrees*** | **3. Lifts head, shoulders and chest when lying on stomach***  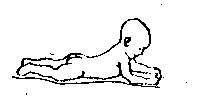 | 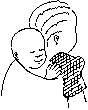**4. Holds head upright for a few seconds*** | 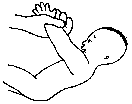**5. Pulls to sit with no head lag*** | **6. Sits with help (with a nice straight back)***  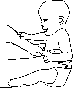 |
| --- | --- | --- | --- | --- | --- |
| 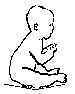**7. Holds head erect continuously when sitting.*** | 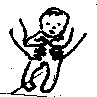**8. Bears weight on legs (holds legs strongly when put in standing position)*** | 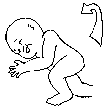**9. Rolls over from back to front*** | **10. Sits by self well.****  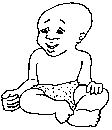 | 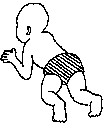**11. Crawls (in any way)**** | 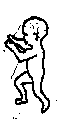**12. Pulls self to stand**** |
| **13. Able to stand if holding on to things****  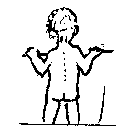 | 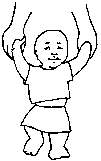**14. Walks using both hands of someone.** | **15. Walks with help using someone’s hand or furniture**  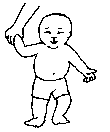 | **16. Walks but falls over at times**  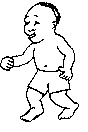 | **17. Stoops over and gets back up**  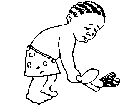 | **18. Walks well**  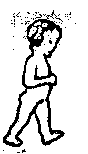 |
| **19. Runs (basic running)**  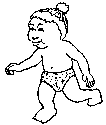 | **20. Kicks a ball (in any way- even only a little bit)**  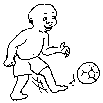 | **21. Runs well, stopping and starting without falling over**  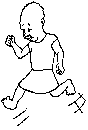 | **22. Does your child climb on to furniture to reach things that they want?** | **23. Throws a ball overhand (raising his arm and shoulder)** | 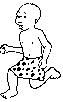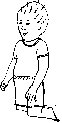**24. Kneels and can get up without using hands.** |
| **25. Climbs and jumps off platform with support** | **26. Jumps off platform on their own** | 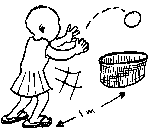**27. Throws a ball in to a basket at 1 metre** | 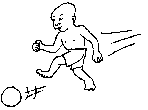**28. Runs, stops and kicks a ball at some distance** | 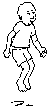**29. Jumps with feet together off the ground** | 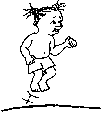**30. Jumps over line on the ground** |
| 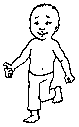**31. Stands on one foot for more than 1 second** | 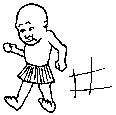**32.Walks on heels six steps** | 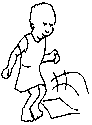**33. Jumps over a piece of paper or larger object.** | 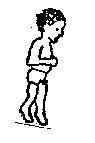**34. Walks on tip toes six steps.** | 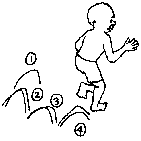**35. Hops on one foot four steps** | 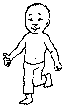**36. Stands on one foot for more than 5 s** |
| **37. Can throw a ball up in the air and catch it with 2 hands.** | **38. Heel/toe walk with one foot behind the other along the string/same line with good balance.** |  |  |  |  |

*Not included in questionnaire for this study ** skipped automatically because of program

**FINE MOTOR & PERFORMANCE**

| 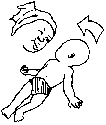**1. Follows mum/carer’s face to mid line. *** | 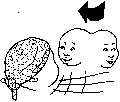**2. Fixes and follows completely**  **from right to left*** | 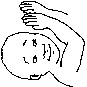**3.Puts hands together in front of eyes or mouth. *** | 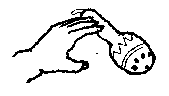 **4.Reaches for a large thing.*** | 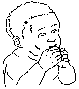**5. When holding objects tends to put them in the mouth*** | **6. Grasps hold of large things.***  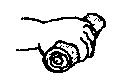 |
| --- | --- | --- | --- | --- | --- |
| 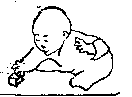**7. Can pick a larger object from the ground such as a wooden spoon.*** | **8. Looks at a small object**  **held in the hand such as a**  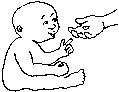 **bean or maize piece. *** | **9. Transfers objects from one hand to another***  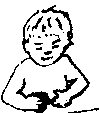 | **10. Picks up small things with four fingers in a raking motion****  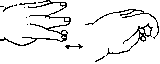 | **11. Strikes one object with another or claps toys or hands together****  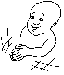 | **12. Finds object hidden under a sheet****  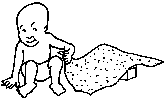 |
| **13. Can use a neat pincer grasp to pick up object between thumb and forefinger****  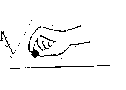 | **14. Puts blocks or stones in and**  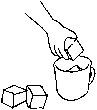**out of a plastic tea cup in imitation.** | **15. Copies pushing a little wooden or wire car along**  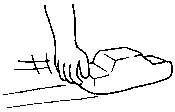 | **16. Puts blocks or 2 cm size stones in and out of a plastic jar in imitation.**  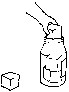 | **17. Dumps blocks out of jar purposefully**  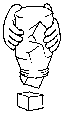 | **18. Scribbles on paper with chalk or on the ground with a stick in straight lines.** |
| **19.Scribbles on paper with chalk or on the ground with a stick in a circular motion** | **20.. Can build a tower of two**  **bricks** | **21. Puts pegs in a board in a longer time. (< 2 min)**  • • • •  • • • • | **22. Ask child to hold four bricks in their hands and to give you one of them (make sure they have four in their hand at the time)** | **23. Makes a tower of four bricks** | **24. Makes a**  **tower of at least six bricks** |
| **25. A Fill up two cups. One with very little water and one with a lot of water and ask the child to give you the cup with more water (do it three times)** | **26 Can do the peg board quicker – within 30 s (<30s)**  • • • •  • • • • | 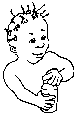**27. Unscrews and screws the cap on and off a peanut butter plastic jar** | **28. Threads six beads**  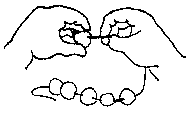 | **29. With two cups of different sizes (same colour) child is able to give you bigger cup. Say to the child, “Give me the bigger one?”** | **30. Imitates a vertical line drawn by the assessor** |
| 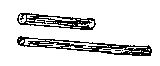**31. Picks the “longer” stick 3 times out of 3 tries** | 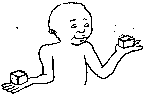**32. Picks the heaviest of**  **two objects 3 times out of 3 tries** | **33.Can make a bridge** | 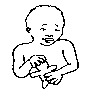**34. Makes a doll or a car out of clay** | **35. Copies a circle drawn on paper or on the sand** | **36. Copies a cross drawn on paper or on the sand** |
| 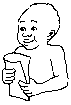**37. Can fold a piece of paper in half** | **38. Can draw a square** | **39. Can make a bridge with 6 blocks** | **40. Can make stairs with 6 blocks** | **41.Can copy a pattern with 4 bottle tops** | **42. Can copy a square pattern with 4 bottle tops** |

*Not included in questionnaire for this study ** skipped automatically because of program

**LANGUAGE / HEARING**

| **1. Startles to sounds*** | **2. Happy making sounds such as “uh”, “eh”, “a” which are separate from crying sounds.*** | 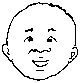**3.Laughs and chuckles*** | **4. Turns to voice – looks in the direction of a voice or a sound. ***  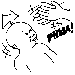 | **5. Makes single syllable sounds such as .. ma/da/pa/ba*** | **6. Responds and turns to his or her name being called.**  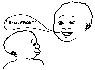 |
| --- | --- | --- | --- | --- | --- |
| **7. Says two syllable babble such as ....dada, lala, mimi, tata,** | 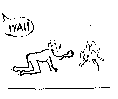**8. Understands when being told “no” or being cautioned** | **9. Shakes head or does something to indicate “No”**  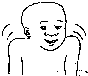 | 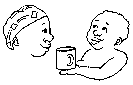**10. Follows one stage commands such as “give me the cup”** | **11. Unclear talk or jabber in sentences..(sounds like sentences but not always clear to the listener)** | **12. Says two words other than mama or dada e.g. cup/cow/spoon/** |
| **13. Says two words together such as “mama-cup” or “dada-water” or “dada goes”** | **14. Says at least six or more words, but words other than those used for mother and father** | **15. Follows two stage commands “go and get the cup over there and put it in the basket”** | **16. Can pull out/identify from the box up to 5 things e.g. cup/ball/spoon/bottle/cloth**  **Say to the child “give me the cup...ball...spoon...bottle..cloth.”More than 5 recognised** | **17. Speaks clearly in sentences that are understood well (three words together at a time).** | **18. Points to more than one body part Ask...” Where is your.... nose/eyes/mouth”...**  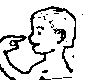**> 1 body part** |
| **19. Names at least five (5) things in the box. Ask the child.... “What is this?” and pull out of basket a cup/spoon, ball, bottle, cloth...**  **Named at least 5.** | **20. Knows his or her first name – When asked, “What is your name?” – can tell you their name”** | **21. Knows actions of objects. “Which one is used for sweeping?” “Which one is used for drinking?” .... Ask them to pick them up.** | **22. Can pull out/identify from the box up to 10 things e.g. cup/ball/spoon/bottle/cloth/soap/ brush/bicycle/plate**  **Say to the child “give me the cup...ball...spoon etc..etc..”**  **More than 10 RECOGNISED** | **23. Names at least ten (10) things in the box. Ask the child.... “What is this?” and pull out of basket a cup/ball/spoon/bottle/cloth/ battery/ brush/bicycle/plate**  **NAMED at least 10** | **24. Can the child talk or explain things that happened in the past e.g talk to you in the past tense about what happened yesterday** |
| **25. Can the child sing songs or repeat rhymes from memory** | **26. Is able to categorise things...“Tell me some things that you eat”...“Tell me some animals you know”Can answer at least one of these questions** | **27 Able to follow a three stage command e.g. “stand up, clap your hands and go over to the....”.?** | **28. Knows the use of objects e.g. “What do you do with a cup?” “What do you do with soap?”** | **29. Can copy 2 syllables that are repeated. Say to the child...When I say this…Copy me:**  **“Pa, Chi, Tu, Go”**  **2 out of 4 eg. Pa, Chi** | **30. Knows the answer of 2 out of 3 questions with an adjective e.g What do you do when you are hungry? What do you do when you are tired? What do you do when you are thirsty?** |
| **31. Understands the term “faster”. E.g. “which goes faster – a car or a bicycle?”** | **32. Can copy 4 syllables that are repeated…Say to the child...When I say this....copy me.. : “Pa, Chi, Tu, Go”4 out of 4______** | **33. Understands prepositions e.g. “Put the stone/block under the cup, put the stone/block on the cup/put it next to the cup/put it behind the cup”. Needs to do 3 of 4 of these.** | **34. Knows opposites e.g. “A man is big, a baby is .....”, “If the sun comes up in the day, the moon comes up in......” (must do 2 of 3 of these)** | **35 Knows quantities…can count some bricks or stones...Put bricks on the table and ask the child...” Can you count these?” Can count up to THREE bricks** | **36 Knows quantities...can count some bricks or stones...**  **Put bricks on the table and ask the child.... “Can you count these?” Can count up to FIVE bricks** |
| **37. Knows quantities – can count up to TEN (10) bricks or stones** | **38. Knows materials eg. What is a cup made of (plastic), a stick (wood), a wire bicycle (wire), a book (paper). 2 of 3 answered.** | **39. Child knows how old they are. Can answer the question: How old are you?** | **40. Child can tell you where he or she currently lives e.g. the village or place** |  |  |

*Not included in questionnaire for this study

**PERSONAL-SOCIAL**

| **1. Smiles but not at a particular person. ***  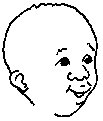 | **2. Smiles in response to a person.***  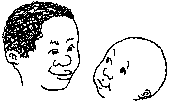 | 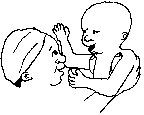**3. Frolics with mother or caregiver in response to being played with.*** | 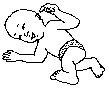**4. Frolics alone, happily playing, moving around moving body and kicking legs in a happy way.*** | **5.Recognises or settles (quiets) with caregivers/known family members. Stops crying or quiets when mum or another known carer takes the baby.*** | **6. Will take phala (porridge) from a spoon when fed by a caregiver***  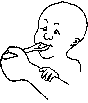 |
| --- | --- | --- | --- | --- | --- |
| **7.Helps hold a cup while mum gives a drink.***  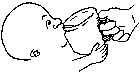 | **8. Stretches to be picked up.**  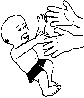 | **9. Can HOLD a spoon with phala (porridge) but NOT get to mouth well yet.** | 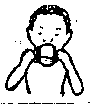 **10. Drinks from a cup by self without spilling.** | 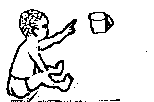**11. Is able to indicate by pointing that they want something.** | **12. Can eat by picking up morsels (of nsima/maize) made by mum.** |
| 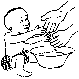 **13. Puts hands out to be washed by mum or carer – helps by putting hands out.** | **14. Can hold a spoon and take phala (porridge) by self but spills some/a little bit**  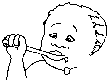 | **15.Indicates in some way the need for a poo or a pee. For example by crying, pulling at pants or saying something...** | **16. Wants to join in singing games. Ask “Does the child like to be included in singing games even if he/she can not yet do them?”** | 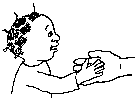**17. Able to greet either by extending hand or verbally.** | **18. Understands to share things with others and will do so if with friends or family and asked to share.** |
| **19. Does a poo or a pee by themselves without wetting their pants.** | **20. Eats phala (porridge) off a spoon without spilling.**  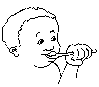 | **21. Does the child play by pretending objects are something else?** | **22. Will the child pretend to cook or make something (like porridge) with imaginary play things e.g tin and sand?** | **23. Can make own morsels of nsima/maize and put in mouth (often with relish)** | 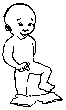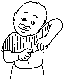**24. Is able to undress by themselves – can take off even one item of clothing e.g. shorts or skirt..need to be able to remove it completely.** |
| **25. Wants to go and visit a friend’s house… shows independence** | 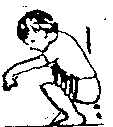**26.Can go to the toilet by themselves anywhere (not necessarily at the pit latrine)** | **27. Can eat food or relish with bits or bones in it such as fish with bones or tangerines with seeds** | **28. Is able to dress but not completely. Can put on at least one item e.g. T-shirt/skirt**  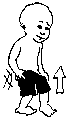 | **29. Washes hands well by self before/after eating**  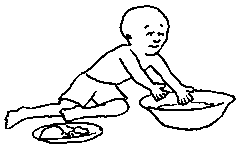 | **30. Knows how to keep quiet at important meetings or ceremonies.**  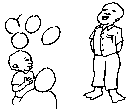 |
| **31.Does household chores in a useful way e.g. drawing water or hoeing. could be even a small amount of help but not just pretend.** | **32. Able to dress by self completely. May need help with buttons, shoe, laces or zips.**  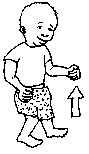 | **33. Understands the concept of discipline e.g. causes and consequences such as that saying bad words might lead to punishment...** | **34. Plays games with turn taking such as fulaye, mira, jingo.**  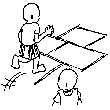 | **35. Knows how to be respectful to elders. Is polite and shows respect such as putting hands together or kneeling before elders** | **36. Can go to the toilet or pit latrine by self.**  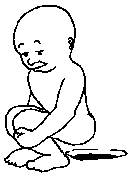 |

*Not included in questionnaire for this study

65.	WHO. Community-based management of servere acute malnutrition. In: World Health Organization WFP, United Nations System Standing Committee on Nutrition, United Nations Children's Fund, ed.2007.

74.	Wang DH, Wang Z, Chen R, Kothapalli KSD, Brenna JT. Very Long-Chain Branched-Chain Fatty Acids in Chia Seeds: Implications for Human Use. Journal of Agricultural and Food Chemistry 2020.

75.	Wang DH, Wang Z, Chen R, Brenna JT. Characterization and Semiquantitative Analysis of Novel Ultratrace C10-24Monounsaturated Fatty Acid in Bovine Milkfat by Solvent-Mediated Covalent Adduct Chemical Ionization (CACI) MS/MS. Journal of Agricultural and Food Chemistry 2020;68:7482-9.

76.	Taha AY. Linoleic acid–good or bad for the brain? npj Science of Food 2020;4.
